# Characterizing Long COVID in an International Cohort: 7 Months of Symptoms and Their Impact

**DOI:** 10.1101/2020.12.24.20248802

**Authors:** Hannah E. Davis, Gina S. Assaf, Lisa McCorkell, Hannah Wei, Ryan J. Low, Yochai Re’em, Signe Redfield, Jared P. Austin, Athena Akrami

## Abstract

Growing evidence shows that a significant number of patients with COVID-19 experience prolonged symptoms, known as Long COVID. Few systematic studies exist which investigate this population, and hence, relatively little is known about the range in symptom makeup and severity, expected clinical course, impact on daily functioning, and expected return to baseline health. In this study, we analysed responses from 3,762 participants with confirmed (diagnostic/antibody positive; 1,020) or suspected (diagnostic/antibody negative or untested; 2,742) COVID-19, from 56 countries, with illness duration of at least 28 days. 3608 (96%) reported symptoms beyond 90 days. Prevalence of 205 symptoms in 10 organ systems was estimated in this cohort, with 66 symptoms traced over seven months. Except for loss of smell and taste, the prevalence and trajectory of all symptoms were similar between groups with confirmed and suspected COVID-19. Respondents experienced an average of 14.5 symptoms in an average of 9.08 organ systems. Three clusters of symptoms were identified based on their prevalence over time. The most likely early symptoms were fatigue, dry cough, shortness of breath, headaches, muscle aches, chest tightness, and sore throat. The most frequent symptoms reported after month 6 were fatigue, post-exertional malaise, and cognitive dysfunction. Majority (>85%) experienced relapses, with exercise, physical or mental activity, and stress as the main triggers. 1,700 (45.2%) reported requiring a reduced work schedule compared to pre-illness and 839 (22.3%) were not working at the time of survey due to their health conditions.

**Significance Statement:** Results from our international online survey of 3,762 individuals with suspected or confirmed COVID-19 illness suggest that Long COVID is composed of heterogeneous post-acute infection sequelae that often affect multiple organ systems, with impact on functioning and quality of life ranging from mild to severe. This study represents the largest collection of symptoms identified in the Long COVID population to date, and is the first to quantify individual symptom trajectory over time, for 7 months. Three clusters of symptoms were quantified, each with different morphologies over time. The clusters of symptoms that persist longest include a combination of the neurological/cognitive and systemic symptoms. The reduced work capacity because of cognitive dysfunction, in addition to other debilitating symptoms, translated into the loss of hours, jobs, and ability to work relative to pre-illness levels.

**Structured Abstract:** *Objective:* To characterize the symptom profile and time course in patients with Long COVID, along with the impact on daily life, work, and return to baseline health.

*Design:* International web-based survey of suspected and confirmed COVID-19 cases with illness lasting over 28 days and onset prior to June 2020.

*Setting:* Survey distribution via online COVID-19 support groups and social media

*Participants:* 3,762 respondents from 56 countries completed the survey. 1166 (31.0%) were 40-49 years old, 937 (25.0%) were 50-59 years old, 905 (24.1%) were 30-39 years old, 277 (7.4%) were 18-29 years old, and 477 (12.7%) were above 60 years old. 2961 (78.9%) were women, 718 (19.1%) were men, and 63 (1.7%) were nonbinary. 317 (8.4%) reported being hospitalized. 1020 (27.1%) reported receiving a laboratory-confirmed diagnosis of COVID-19. 3608 (96%) reported symptoms beyond 90 days.

*Results:* Prevalence of 205 symptoms in 10 organ systems was estimated in this cohort, with 66 symptoms traced over seven months. Except for loss of smell and taste, the prevalence and trajectory of all other symptoms are similar between confirmed (diagnostic/antibody positive) and suspected groups (diagnostic/antibody negative or untested). Respondents experienced symptoms in an average of 9.08 (95% confidence interval 9.04 to 9.13) organ systems. The most frequent symptoms reported after month 6 were: **fatigue** (77.7%, 74.9% to 80.3%), **post-exertional malaise** (72.2%, 69.3% to 75.0%), and **cognitive dysfunction** (55.4%, 52.4% to 58.8%). These three symptoms were also the three most commonly reported overall. In those who recovered in less than 90 days, the average number of symptoms peaked at week 2 (11.4, 9.4 to 13.6), and in those who did not recover in 90 days, the average number of symptoms peaked at month 2 (17.2, 16.5 to 17.8). Respondents with symptoms over 6 months experienced an average of 13.8 (12.7 to 14.9) symptoms in month 7. 85.9% (84.8% to 87.0%) experienced relapses, with exercise, physical or mental activity, and stress as the main triggers. 86.7% (85.6% to 92.5%) of unrecovered respondents were experiencing fatigue at the time of survey, compared to 44.7% (38.5% to 50.5%) of recovered respondents. 45.2% (42.9% to 47.2%) reported requiring a reduced work schedule compared to pre-illness and 22.3% (20.5% to 24.3%) were not working at the time of survey due to their health conditions.

*Conclusions:* Patients with Long COVID report prolonged multisystem involvement and significant disability. Most had not returned to previous levels of work by 6 months. Many patients are not recovered by 7 months, and continue to experience significant symptom burden.

## Introduction

Public discourse on COVID-19 has largely centered around those with severe or fatal illness [1]. As prevention efforts have focused on minimizing mortality, the morbidity of COVID-19 illness has been underappreciated. Recent studies show that a growing number of patients with COVID-19 will experience prolonged symptoms, the profile and timeline of which remains uncertain [2–6]. Early in the course of the pandemic, patients identified this trend, referring to themselves as “Long-Haulers” and the prolonged illness as “Long COVID” [7]. Nonetheless, there exist few systematic studies investigating this population, and hence, relatively little is known about the range in symptom makeup and severity, expected clinical course, impact on daily functioning, and expected return to baseline health [8]. In this paper, we report results from an online survey investigating the symptoms of Long COVID in patients with illness onset between December 2019 and May 2020, allowing analysis of symptoms over an average 6 months’ duration.

While as of yet there is no agreed upon case definition of Long COVID [6, 9], we define the illness as a collection of symptoms that develop during or following a confirmed or suspected case of COVID-19, and which continue for more than 28 days [10]. The few studies that exist on Long COVID document fluctuating and unpredictable symptoms which can affect multiple organ systems at once and/or over time [3,5,11].

### Objectives of study

The aim of this study is to better describe the patient experience and recovery process in those with confirmed or suspected COVID-19 illness, with a specific emphasis on the Long COVID experience. The unique approach of this study utilizes patient-driven research in order to establish a foundation of evidence for medical investigation, improvement of care, and advocacy for the Long COVID population. The survey was created by a team of patients with COVID-19 who are members of the Body Politic online COVID-19 support group. The group conducted its first survey in April 2020 and issued a subsequent report in May 2020 [5]. In order to better investigate additional aspects of patient experience, a second survey was developed, emphasizing symptom course and severity over time with an in-depth look into neurological and neuropsychiatric symptoms, recovery, and return to baseline, including work impact. Other topics investigated in the survey will be included in future reports.

## Methods

### Study design

Data were collected using an online survey hosted on the Qualtrics platform. All respondents gave digital informed consent prior to participating. Survey responses contained no personally identifiable information, and email addresses collected for survey distribution were hashed into anonymized ID codes. The study was approved by the UCL Research Ethics Committee [16159.002] (London, UK), and Oregon Health and Science University Institutional Review Board (IRB) (Portland, OR, USA), with UCL serving as the primary site. The Weill Cornell Medical College IRB determined non-engagement.

The survey consisted of 257 questions and required a median time of 69.3 minutes to complete. 61.6% of respondents completed the survey after giving consent. Methods used to distribute the survey did not allow us to determine the number of people who viewed the invitation, so the response rate could not be calculated. To account for post-exertional malaise and brain fog, which are common Long COVID symptoms that limit sustained focus and attention, respondents were encouraged to take breaks while completing the survey. Progress was saved for up to 30 days to allow respondents to return to the survey at a later time.

The survey was created in English and translated into eight additional languages: Spanish, French, Portuguese, Italian, Dutch, Russian, Bahasa Indonesian, and Arabic. Links to the survey were disseminated via email, social media, and online patient support groups listed in Appendix A. Data included in the analysis were collected from September 6 to November 25, 2020.

### Patient and Public Involvement

The study was designed and conducted by a team of patients with Long COVID (all authors), who formed the Patient-Led Research Collaborative in May 2020. During the curation of survey questions, inputs from patients on various support groups, including Body Politic COVID-19 Slack support group, and Long COVID Support group on Facebook, were incorporated. In specific, we worked closely with other patients to compile the list of symptoms, design research questions on how the Long Covid condition may affect daily life of the patients, and optimise the questionnaire design in order to reduce survey fatigue.

### Participants - Inclusion criteria

The survey “Information Sheet” stated: “You are being invited to participate in this research study because you have had a COVID-19, or suspected COVID-19 infection (still suffering or suffered symptoms) for longer than 1 week and you are 18 years of age or older.” All respondents consented to these criteria. To characterize Long COVID symptoms over an extended period, analysis was limited to respondents with illness lasting longer than 28 days and symptom onset between December 2019 and May 2020.

A total of 7,285 responses were downloaded from the Qualtrics server on November 25, 2020. The following responses were removed from the dataset: incomplete (those not reaching the end of the survey, n=2,367), no illness onset date (n=2), onset date before December 2019 (n=26), 0 days of symptoms (n=1), duplicate users with the same IDs (same email addresses with same demographic information, n=150), symptoms for 28 days or less (n=401), and illness onset after May 2020 (n=576). This resulted in complete data from 3,762 respondents.

#### SARS-CoV-2 test results

In addition to positively tested subjects [n=1,020, either diagnostic (RT-PCR/antigen, Table 1) or antibody], we included participants with absent (n=1,819) or negative test results (n=923, diagnostic and antibody), since the prevalence of 203 symptoms (out of 205, see Section “*Symptoms by test results*”), as well as symptom trajectory (Figure 7) and the survival functions (Figure 1a), were not significantly different between diagnostic/antibody positive and negative or untested groups. This strongly suggests SARS-CoV-2 infection in the diagnostic/antibody negative (or untested) group, and we therefore included respondents who did not test positive with a SARS-CoV-2 diagnostic or antibody test in most of our analyses (see Supplemental Figures S7-S8 for separate analyses).

**Figure 1.**
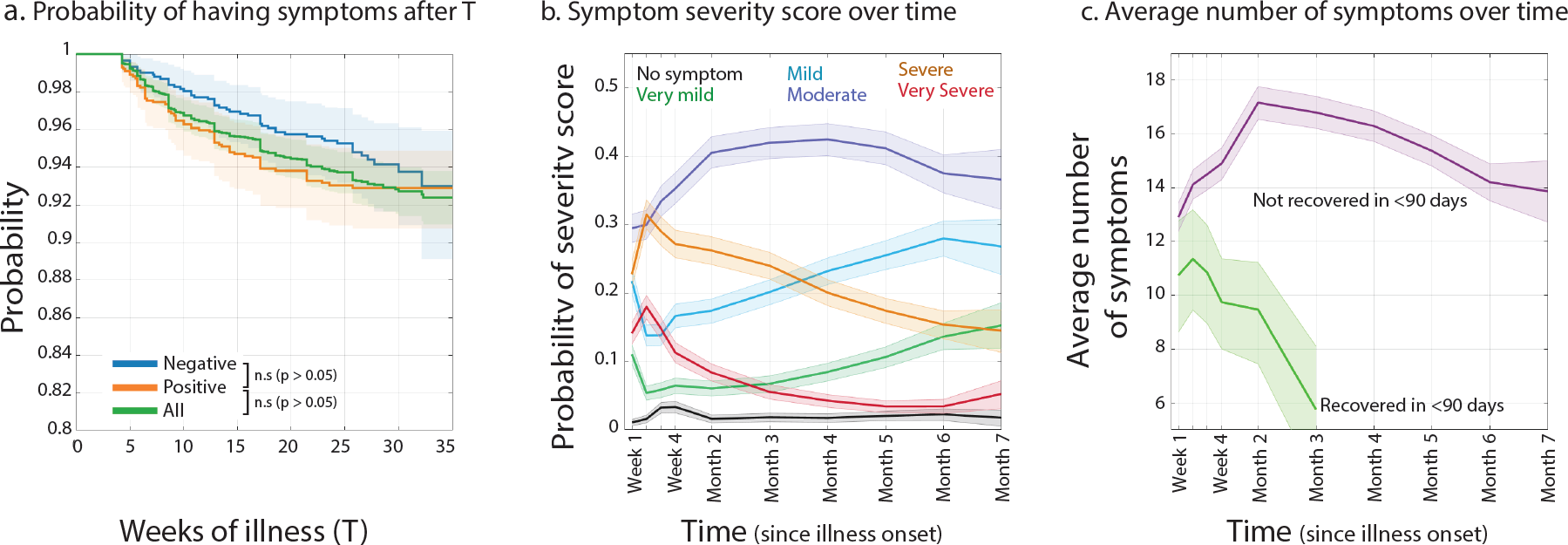
a) Survival function (Kaplan-Meier estimator), characterizing the distribution of disease duration for those who tested Negative (blue) on both diagnostic (RT-PCR/antigen) and antibody tests, those who tested Positive (orange) in either diagnostic or antibody test, and All (green) respondents. The Y axis indicates the probability that symptoms will persist longer than the time specified on the X axis. b) Probability of each symptom severity score over time. c) Average number of reported symptoms over time for those who recovered in less than 90 days (n=154), or otherwise experienced symptoms for more than 90 days (n=3505). a-c) In all plots, times are relative to initial symptom onset. Shaded regions represent 95% simultaneous confidence bands.

**Table 1.** Testing status.

| Type of SARS-CoV-2 Test | Number of Respondents Tested | % of Respondents Tested* | Number of Respondents with Positive Results | % of Respondents with Positive Results* |
| --- | --- | --- | --- | --- |
| Diagnostic (RT-PCR/antigen) | 2330** | 61.9% | 600 | 15.9% |
| Antibody (IgG, IgM or both) | 2166 | 57.6% | 683 | 18.2% |
| Diagnostic (RT-PCR/antigen) or Antibody | 3121 | 83.0% | 1020 | 27.1% |

\*Percentages are out of the total number of respondents (N=3,762).
\*\*Total of 2362 received diagnostic tests, out of which 32 were inconclusive or awaiting response.

### Statistics and data analysis

#### Variables

In this study we quantified symptom prevalence, probability time-course, severity, count, onset time, temporal clustering. We also measured fatigue using Fatigue Assessment Scale. Return to baseline and working status were also measured.

##### Prevalence estimation

205 symptoms were investigated by identifying their presence or absence. For 74 of these symptoms, respondents indicated at which points in their illness (weeks 1-4, months 2-7) they experienced the symptom, if at all. For each of the other 131 symptoms, participants indicated whether they had experienced the symptom at any point throughout the duration of their illness (Figures 2, 3). Prevalence estimates were calculated by dividing the number of those who identified experiencing a symptom, either at a given time point (Figure 4) or over the entire sickness period (Figure 2, 3), by the total number of participants to which the symptom applied. Eight symptoms were excluded from analysis, as their measurement required specialized equipment or tests that many participants may not have had access to. Excluded symptoms were high blood pressure, low blood pressure, thrombosis, seizures (confirmed or suspected), low oxygen levels, high blood sugar, and low blood sugar. The remaining 66 symptoms (out of 74) were included in analysis of the timeline of disease progression over 7 months (see below, Figure 4). Each symptom was further categorized into one of ten organ systems (Supplemental material, Appendix A), which were visualized as groups. The respondents for certain symptoms (non-primary language and reproductive/genitourinary symptoms) consisted of the subset of total respondents whom the symptom could apply to (i.e. those who spoke more than one language, cisgender female or non-binary and menstruating, cisgender female or non-binary and above or below 40 years of age, and male). Therefore, the symptom prevalence was calculated within the relevant subsample.

**Figure 2:**
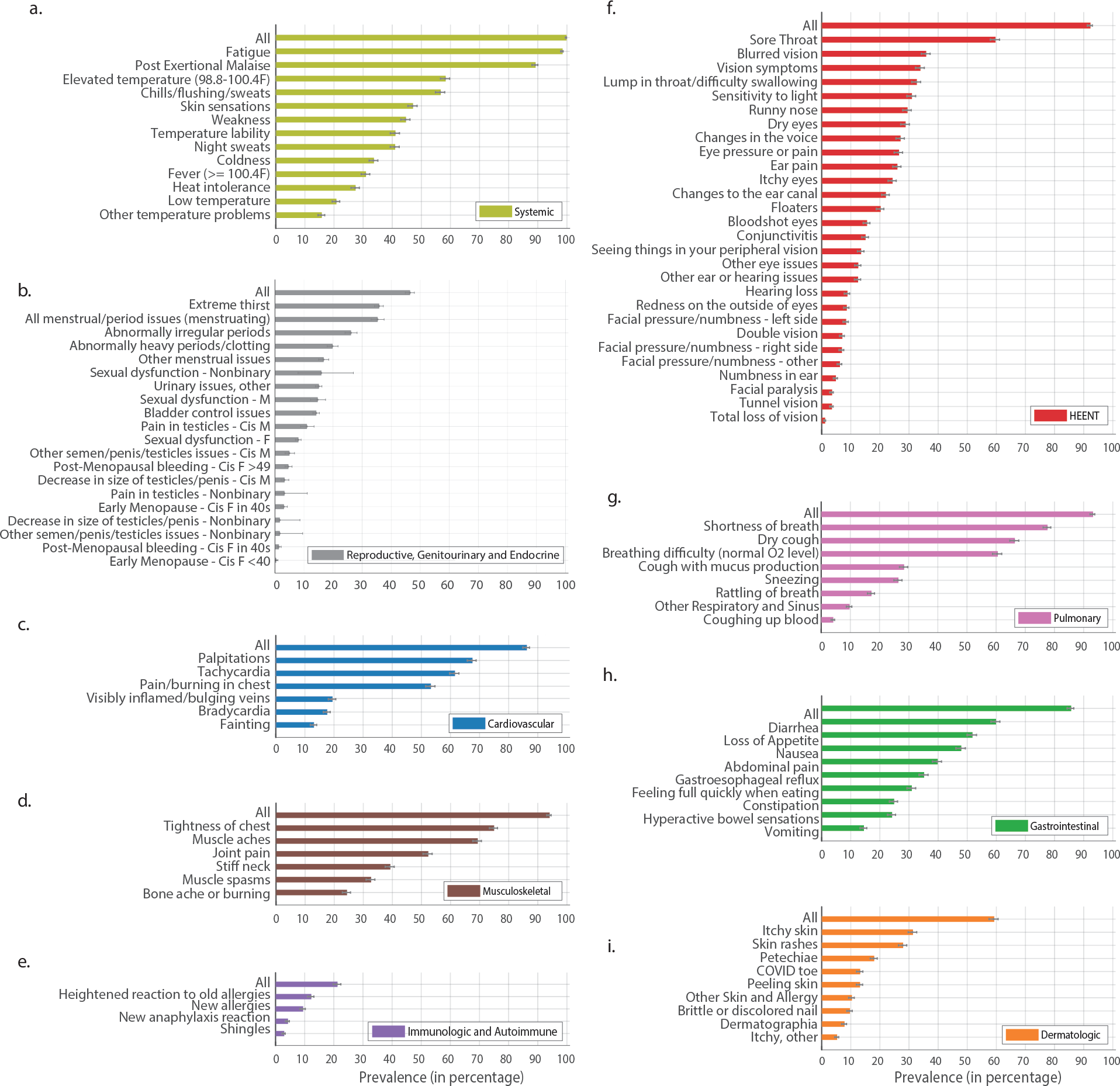
Symptom prevalence estimates (non-neuropsychiatric symptoms). Bars represent the percentage of respondents who experienced each symptom at any point in their illness. Symptoms are categorized by the affected organ systems. When all rows in a given panel use the same denominator, the first row, labeled “All,” indicates the percentage of respondents who experienced any symptoms in that category. Error bars are 95% confidence intervals.

##### Symptom time course estimation

The survey asked respondents to detail their experience of a subset of 66 symptoms over time. Respondents indicated whether each of these symptoms was present during a series of time intervals following the onset of their first symptoms: week 1 (days 1-7), week 2 (days 8-14), week 3 (days 15-21), week 4 (days 22-30), month 2 (days 31-60), month 3 (days 61-90), month 4 (days 91-120), month 5 (days 121-150, month 6 (days 151-180), and month 7 (days 181-210).

The time course of each symptom was defined as the probability of experiencing the symptom in each time interval, given that: 1) recovery had not occurred prior to the end of the interval, and 2) the symptom was applicable (menstruation-related symptoms are presented only for menstruating respondents). Probabilities were estimated for each interval as the fraction of respondents who experienced the symptom, among those who satisfied the two criteria above. The number of qualifying respondents in each time interval is given in Table 2.

**Table 2.** Number of qualifying respondents in each time interval.

| Symptoms (66) | Week 1 | Week 2 | Week 3 | Week 4 | Month 2 | Month 3 | Month 4 | Month 5 | Month 6 | Month 7 |
| --- | --- | --- | --- | --- | --- | --- | --- | --- | --- | --- |
| Menstruation Related (1) | 1792 | 1792 | 1792 | 1792 | 1757 | 1726 | 1704 | 1615 | 1190 | 462 |
| Other Symptoms (65) | 3762 | 3762 | 3762 | 3762 | 3681 | 3624 | 3563 | 3352 | 2454 | 966 |

Plotted time courses in Fig. 4 are linearly interpolated between the centers of each time interval.

##### Symptom severity and count

Overall symptom severity for each time interval (weeks 1-4, month 2-7) was measured using Likert scale (no symptom, very mild, mild, moderate, severe, very severe). The probability of each Likert option was calculated as the fraction of qualifying participants (as described above) who selected that option (Figure 1b). Total number of experienced symptoms (from the subset of 66) was measured for each qualifying respondent in each time interval. The mean value of symptom count was then calculated by averaging over all qualifying respondents.

##### Symptom onset analysis

The heatmap in Figure 5 shows the probability distribution of onset times for each symptom. Continuous, piecewise-constant distributions were fit using maximum likelihood, accounting for interval censoring (onset times for each respondent could only be measured up to the enclosing time intervals, described above). For each symptom, the estimated probability density at time *t* was given by the fraction of respondents who first experienced the symptom in the interval containing *t* (among those who experienced it at any point), divided by the duration of the interval. Mean onset time was calculated as the expected value of the estimated distribution.

##### Symptom time course clustering

Symptom time courses were clustered using spherical k-means, a variant of k-means based on cosine distances [12]. Each time course is a 10-dimensional vector, representing the conditional probability of experiencing the symptom in each of the 10 time bins (as above). The cosine distance is a monotonic function of the angle between vectors, and is insensitive to their magnitudes. Therefore, it is well suited to measuring differences between time course shapes (i.e. changes in relative amplitude over time), while remaining invariant to differences in overall symptom prevalence. We used a variant of Lloyd’s algorithm designed for spherical k-means, with initialization by the k-means++ algorithm, and 100 random restarts to avoid suboptimal local minima. The number of clusters (*k*=3) was chosen by hand, to provide a reasonable tradeoff between capturing structure in the data and obtaining a parsimonious explanation.

##### Symptom time course sorting

The heatmaps in Figure 6 and Figure S3 show normalized symptom time courses, sorted such that similarly-shaped time courses appear nearby in the ordering. The sort ordering was computed as follows. Similarity between time courses was measured using the cosine distance, as above. Classical multidimensional scaling (MDS) was then used to embed time courses into a one-dimensional Euclidean space, such that pairwise distances in the embedding space approximated the given cosine distances. Time courses were sorted according to their order in the embedding space.

##### Confidence intervals

All confidence intervals and confidence bands were estimated using a nonparametric bootstrap approach with 10,000 iterations. Individual confidence intervals and pointwise confidence bands used the bias-corrected, accelerated (BCa) bootstrap [13]. Simultaneous confidence bands used the percentile bootstrap, with the percentile adjusted to give the correct simultaneous coverage probabilities.

##### Stratification based on the diagnostic test time

Test time, defined as the number of days between first experiencing symptoms and receiving a diagnostic test (RT-PCR or antigen), was significantly shorter in respondents with positive results compared to those with negative results (see *Symptoms by test results,* and Supplemental Figure S6). To compare the symptom prevalence and time course (Supplemental Figure S7-S8), the two groups were stratified based on the test time by including respondents who were tested within certain time windows after illness onset. For prevalence estimates, time windows were ‘smaller than 10 days’, ‘between 10 and 20 days’, and ‘larger than 20 days’. For time course estimates, we limited the comparison to those who have been tested within 20 days after illness onset. Data stratification based on test time was done only for the diagnostic test.

##### Text analysis

The survey asked respondents to elaborate on their experience in free text for the following areas: body parts for sensorimotor symptoms, brain fog and memory issues, most debilitating symptoms, other diagnosis post illness, and work status. Deductive thematic analysis was used to tag and extract themes around impact on work [14]. For textual input on participants’ experience of symptoms, such as cognitive dysfunction, a range of quotes was selected to provide a deepened understanding of the diversity of experiences [15]. Identifying data were anonymized and longer sentences were truncated for brevity. For the sensorimotor textual input questions, which asked which body part was affected, natural language processing was used in Python. The text was converted to lowercase, stripped of punctuation and extra whitespace, and stopwords were removed (using the original stopwords list from the NLTK library as well as common non-symptom text inputs [16]). The text was tagged for the parts of speech using a word tokenizer, and only nouns were reserved. The nouns were run through a translation function to convert all non-English nouns to their English counterparts, then counted, and the top four body parts were added to the table. The answers to “most debilitating symptoms’’ followed a similar process, without the parts-of-speech tagging; another function was written to group similar descriptions (e.g. cognition, brain fog, and difficulty concentrating all went under “cognitive dysfunction”).

## Results

### Demographics

This study included 3,762 survey respondents based on the eligibility criteria described above. Detailed demographics are listed in Table 3. The majority of respondents were women (78.9%, significantly more than other genders, p < 0.001, chi-squared test), white (85.3%, p < 0.001, chi-squared test), and between the ages of 30 and 60 (33.7% between ages 40-49, 27.1% ages 50-59, 26.1% ages 30-39). A total of 56 countries were represented in the sample. Most of the respondents resided in the United States (41.2%, p < 0.01, Tukey’s HSD multiple comparisons test). 91.9% of respondents completed the survey in English.

**Table 3.** Demographics of survey respondents.

| Factor | Number of Respondents (N=3,762) | % of Respondents |
| --- | --- | --- |
| **Gender** |  |  |
| Women* | 2969 | 78.9% |
| Men* | 718 | 19.1% |
| Nonbinary | 63 | 1.7% |
| Other | 6 | 0.2% |
| Preferred not to answer | 6 | 0.2% |
| **Age Group** |  |  |
| 18-29 | 277 | 7.4% |
| 30-39 | 905 | 24.1% |
| 40-49 | 1166 | 31.0% |
| 50-59 | 937 | 25.0% |
| 60-69 | 380 | 10.1% |
| 70-79 | 85 | 2.3% |
| 80+ | 12 | 0.3% |
| **Ancestry\*\*** |  |  |
| White | 3418 | 85.3% |
| Hispanic, Latino, Spanish Origin | 150 | 3.7% |
| Asian, South Asian, SE Asian | 134 | 3.3% |
| Black | 80 | 2.0% |
| Middle Eastern, North African | 66 | 1.7% |
| Indigenous Peoples | 50 | 1.6% |
| Pacific Islander | 3 | 0.1% |
| Other | 98 | 2.5% |
| Prefer not to answer | 9 | 0.2% |
| **Environment** |  |  |
| Urban | 1543 | 41.0% |
| Suburban | 1586 | 42.2% |
| Rural | 633 | 16.8% |
| **Country of Residence** |  |  |
| USA | 1567 | 41.2% |
| UK and Northern Ireland | 1316 | 35.0% |
| France | 163 | 4.3% |
| Canada | 155 | 4.1% |
| Spain | 99 | 2.6% |
| Netherlands | 61 | 1.6% |
| Ireland | 58 | 1.5% |
| Sweden | 55 | 1.5% |
| Other | 288 | 7.7% |
| **Healthcare Worker** |  |  |
| Yes | 668 | 17.8% |
| No | 3094 | 82.2% |

| Hospitalization |  |  |
| --- | --- | --- |
| Non-Hospitalized | 2133 | 56.7% |
| Visited ER or Urgent Care | 1312 | 34.9% |
| Hospitalized | 317 | 8.4% |

\*Respondents included 2961 (78.7%) cisgender women and 8 (0.2%) transgender women, 714 (19.0%) cisgender men and 4 (0.1%) transgender men.
\*\*Respondents were invited to select multiple ancestries. Percentages in this section are thus based on the total number of ancestries reported. 182 (4.8%) respondents reported two ancestries, while 30 (0.8%) reported three or more ancestries.

More than half of respondents (56.7%, p < 0.001, chi-squared test) did not seek hospital-based care. 34.9% visited an ER or urgent care clinic but were not admitted to a hospital. 8.43% of respondents were hospitalized. 17.8% of respondents were healthcare workers (see Supplemental Material, Appendix A, for pre-existing conditions).

### Symptoms and severity over time

#### Symptom duration

Respondents were considered recovered if they identified themselves as no longer experiencing symptoms at the time of survey completion. 257 respondents (6.8%) recovered after day 28 of illness, and 3,505 (93.2%) were still experiencing symptoms at the time of survey completion.

To investigate disease duration, the survey asked respondents to indicate the number of days their symptoms lasted. For non-recovered respondents, this number provided only a lower bound on the eventual duration of symptoms. To account for this censoring in the data, we characterized the distribution of durations using the Kaplan-Meier estimator [17]. The resulting survival function (Figure 1a, Supplemental Figure S1a for Male vs Female comparison) measures the probability that symptoms will persist beyond any specified amount of time. In this Long COVID cohort, the probability of symptoms lasting beyond 35 weeks was 91.8% (95% confidence interval 89.5% to 93.5%), with no statistically significant difference between positively (diagnostic/antibody) and negatively tested groups (p = 0.18, chi-squared test), or men and women (p=0.49, chi-square test, Supplemental Figure S5). Of the 3,762 respondents, 2,454 experienced symptoms for at least 180 days (6 months). Among the remaining 1,308 respondents, 233 recovered and the rest (n=1,075) took the survey before reaching 6 months of illness.

The trajectory of Long COVID can be described by assessing symptom severity and average number of symptoms over time. The probability of each of the severity score Likert options is illustrated as a function of time (Figure 1b, Methods) to demonstrate the evolution in symptom severity throughout the course of illness. The probability of “severe” and “very severe” symptoms peaked during acute infection (<28 days), while the probability of “moderate” and “mild” rose gradually thereafter. The majority of respondents reported “moderate” symptoms throughout the course.

In those who recovered in less than 90 days, the average number of symptoms peaked at week 2 (mean number of symptoms: 11.35, 95% confidence interval 13.58 to 9.44), and in those who did not recover in 90 days, the average number of symptoms peaked at month 2 (mean number of symptoms: 17.16, 17.78 to 16.54), with less of a decline over time (Figure 1c, see Supplemental Figure S1 b-c for more comparisons between recovered and unrecovered participants).

Respondents with symptoms for over 6 months experienced an average of 13.79 symptoms (95% confidence interval 12.68 to 14.88) in month 7.

#### Symptoms experienced at any point

Participants were asked to indicate the presence or absence of 205 symptoms. For 74 of these symptoms, participants indicated at which points in their illness (weeks 1-4, months 2-7) they experienced the symptom, if at all. Eight symptoms were excluded from the main analyses (Methods, Supplemental Figure S5). Symptoms were divided into 10 categories representing the organ system in which they are present (see Appendix A, Table 4). Prevalence estimates for 131 symptoms were estimated as the percentage of respondents who experienced each symptom at any point (Figure 2 for non-neuropsychiatric and Figure 3 for neuropsychiatric symptoms, see Methods). In some cases, such as reproductive symptoms, the denominator used to calculate the percentage varied depending on factors such as respondent age or sex (see Methods).

**Figure 3:**
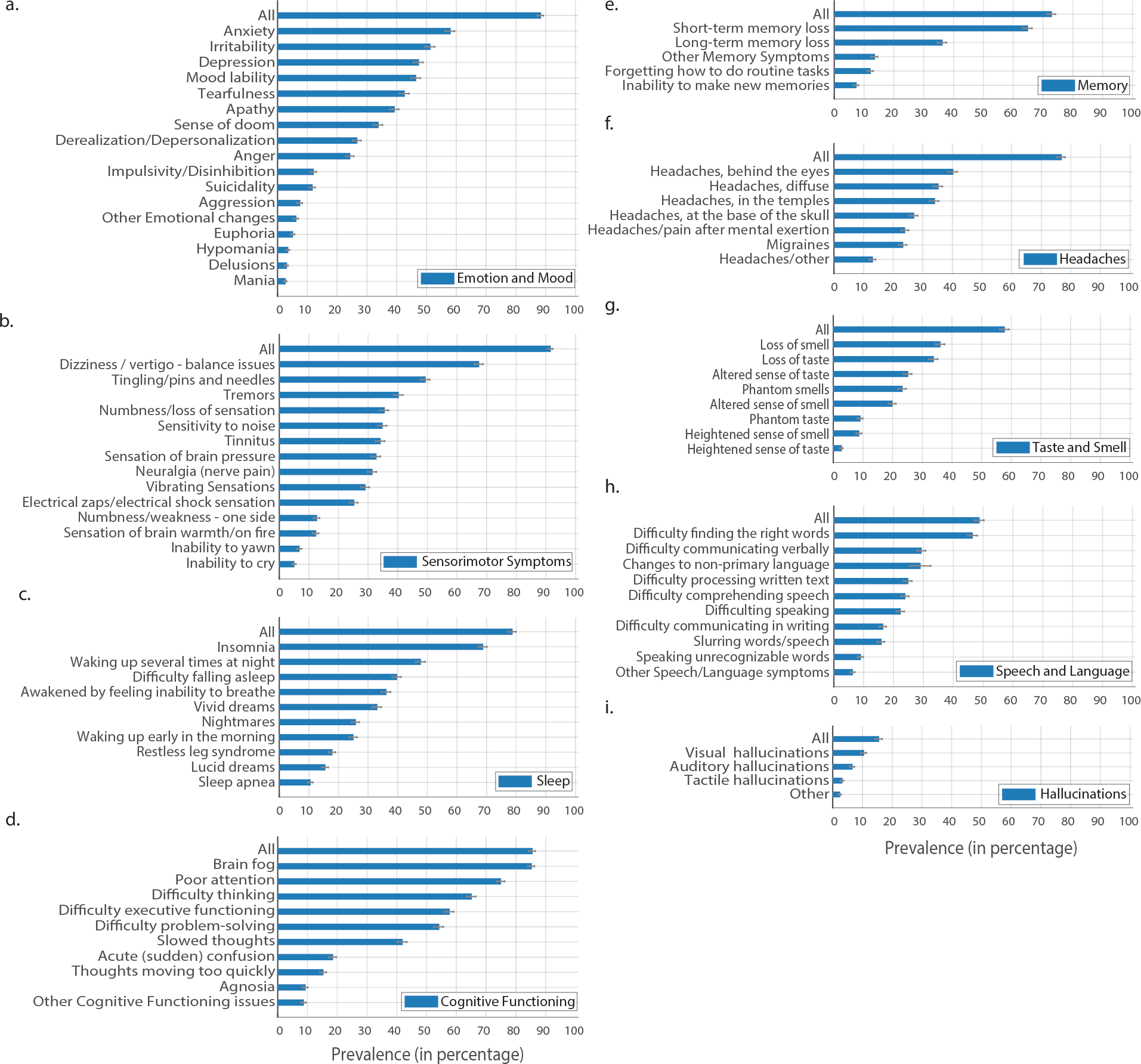
Symptom prevalence estimates for neuropsychiatric symptoms. Similar to Figure 2, for neuropsychiatric symptoms, divided into nine sub-categories. Each bar represents the percentage of respondents who experienced that symptom. Error bars are bootstrap 95% confidence intervals.

**Table 4.** Overall System Prevalence Data.

| Symptom Category | Total # | Mean<br>Prevalence | Lower CI | Upper CI |
| --- | --- | --- | --- | --- |
| Systemic | 3750 | 99.70 | 99.49 | 99.84 |
| Reproductive / Genitourinary / Endocrine | 2341 | 62.25 | 60.68 | 63.74 |
| Cardiovascular | 3236 | 86.04 | 84.90 | 87.16 |
| Musculoskeletal | 3530 | 93.85 | 93.03 | 94.60 |
| Immunologic / Autoimmune | 791 | 21.05 | 19.77 | 22.43 |
| HEENT | 3761 | 100 | 100 | 100 |
| Pulmonary / Respiratory | 3499 | 93.03 | 92.21 | 93.8 |
| Gastrointestinal | 3216 | 85.50 | 84.37 | 86.6 |
| Dermatologic | 2221 | 59.06 | 57.52 | 60.63 |
| Neuropsychiatric - Cognitive Dysfunction | 3212 | 85.43 | 84.29 | 86.55 |
| Neuropsychiatric - Speech and Language | 1828 | 48.62 | 47.00 | 50.21 |
| Neuropsychiatric - Memory | 2739 | 72.81 | 71.40 | 74.20 |
| Neuropsychiatric - Headaches | 2887 | 76.74 | 75.36 | 78.04 |
| Neuropsychiatric - Smell and Taste | 2166 | 57.60 | 56.06 | 59.21 |
| Neuropsychiatric - Sleep | 2955 | 78.58 | 77.25 | 79.88 |
| Neuropsychiatric - Emotion and Mood | 3320 | 88.25 | 87.19 | 89.26 |
| Neuropsychiatric - Hallucinations | 580 | 15.42 | 14.30 | 16.64 |
| Neuropsychiatric - Sensorimotor | 3440 | 91.44 | 90.48 | 92.29 |

#### Symptoms over time

To characterize the progression of the 66 symptoms over seven months, we estimated symptom time courses—the probability of experiencing each symptom at each time point, given that recovery has not yet occurred (Figure 4, see Supplemental Table S22 for the raw data; Supplemental Figure S9 for male vs. female comparison). We also estimated the distribution of onset times for each symptom (Figure 5).

**Figure 4:**
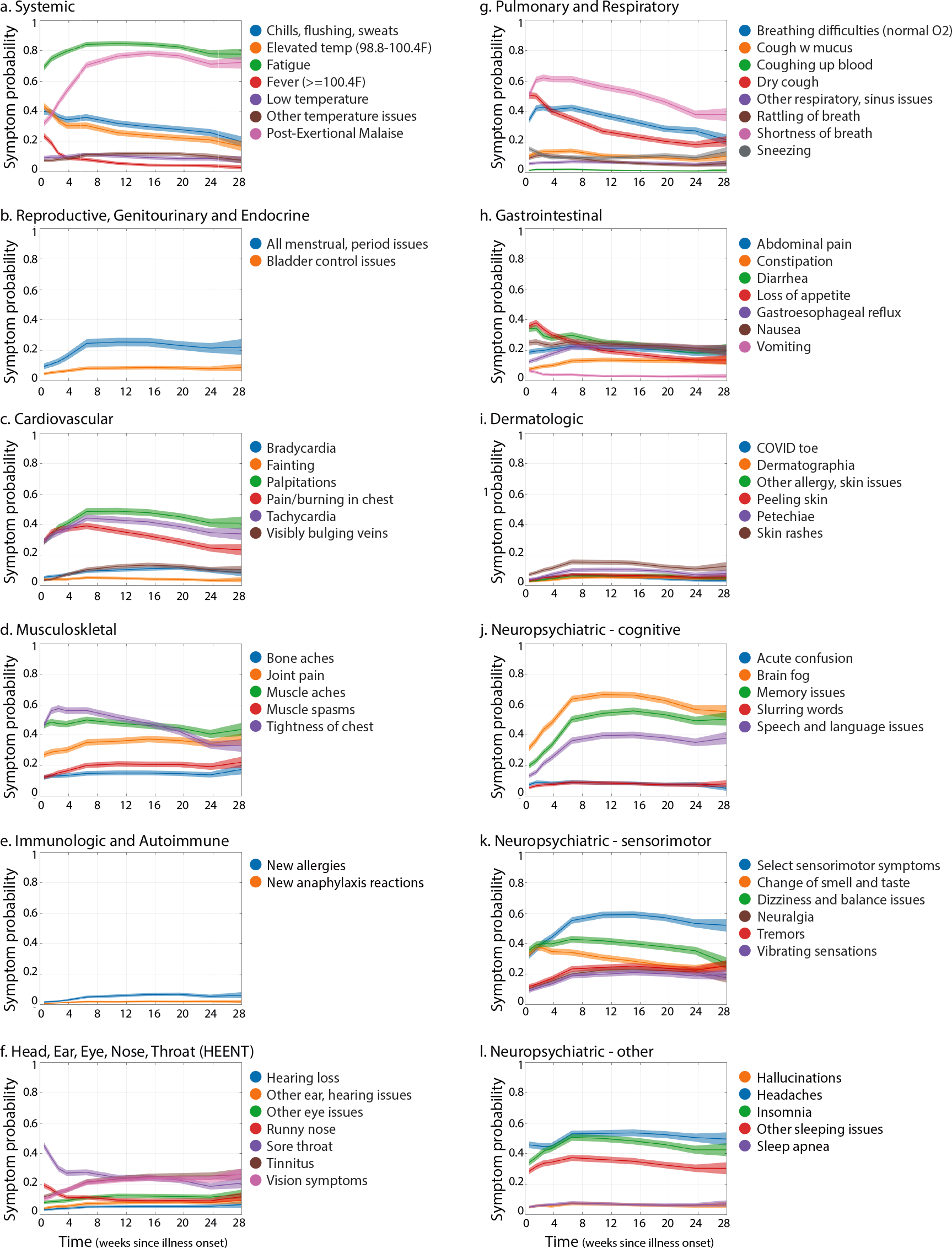
Symptom time courses. Plotted time courses represent the estimated probability of experiencing each symptom at each time point, given that recovery has not yet occurred (see Methods). Times are relative to initial illness onset. Symptoms are grouped according to the affected organ systems. Shaded regions show 95% simultaneous confidence bands, estimated separately for each symptom.

**Figure 5.**
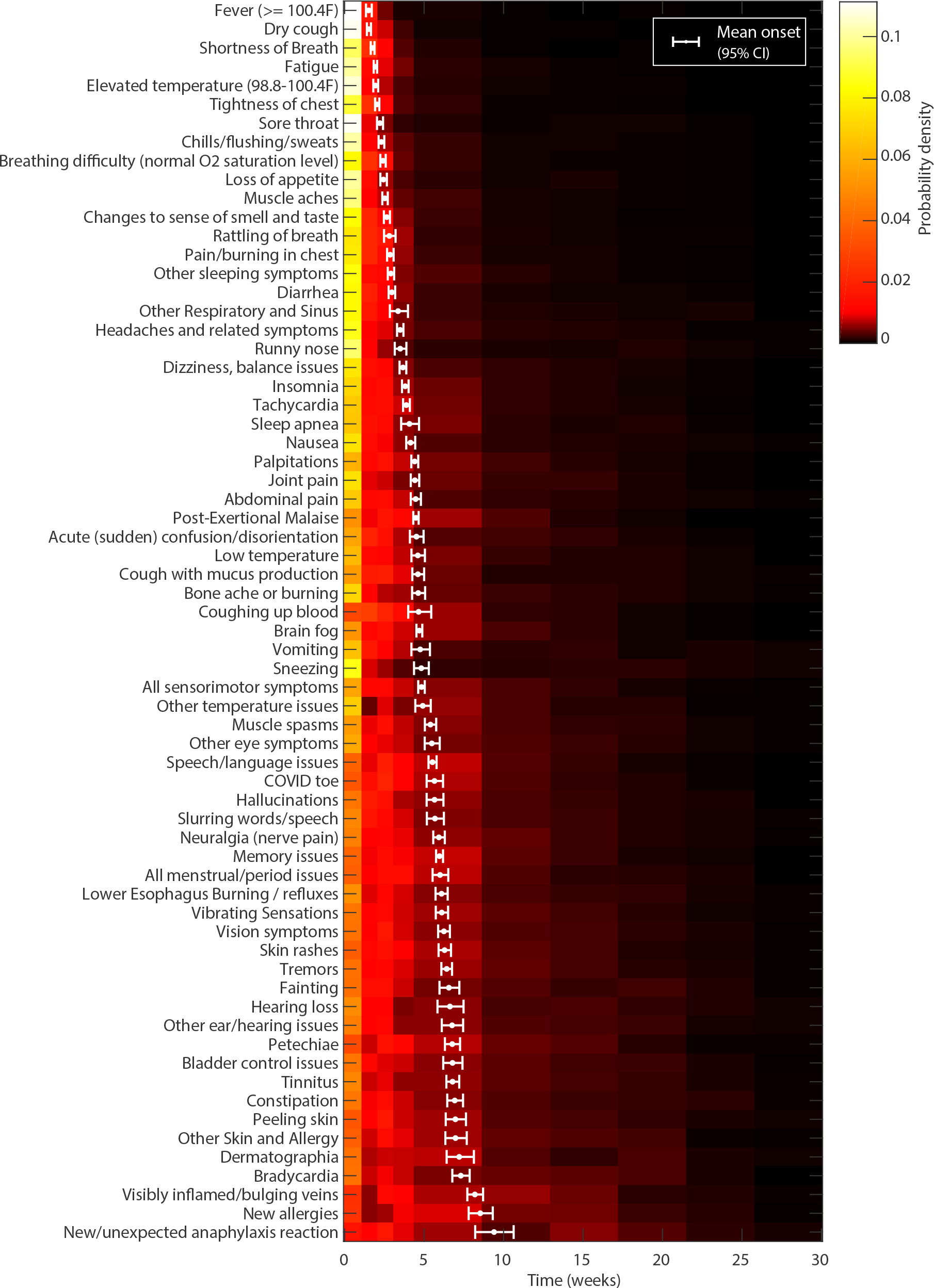
Symptom onset times. Heatmap shows the estimated probability distribution of the onset time for each symptom. White points and error bars show the mean onset time and 95% pointwise confidence intervals. Symptoms are sorted by mean onset time.

To summarize general patterns in the progression of symptoms over time, we used a clustering algorithm (see Methods) to group the 66 symptoms into three clusters, according to the shapes of their time courses (i.e. changes in relative amplitude over time, ignoring their overall prevalence). Symptoms that clustered together generally had similarly-shaped time courses (Figure 6). Cluster 1 consists of symptoms that are most likely to appear early in the illness (reaching a high point in the first two or three weeks), followed by a decreasing trend in probability over time. Cluster 2 consists of symptoms with a slow decrease, slow increase, or unchanging probability over time. On average, symptoms in this cluster exhibit a slightly increased probability of presenting in the second month of illness. Cluster 3 consists of symptoms that are most likely to ramp up sharply in the first two months. Their probability may hit a plateau (like constipation), decrease slightly (like post-exertional malaise and fatigue), or increase slightly in the later months (like tinnitus, hearing loss, muscle spasms, and tremors). All clusters contained symptoms from multiple organ systems, and cluster 3 contained symptoms from all organ systems (with the exception of pulmonary/respiratory symptoms). A general progression from early to late symptoms can also be seen in the heatmap of normalized time courses (Figure 6, S2), which have been sorted by similarity in shape (see Methods).

**Figure 6.**
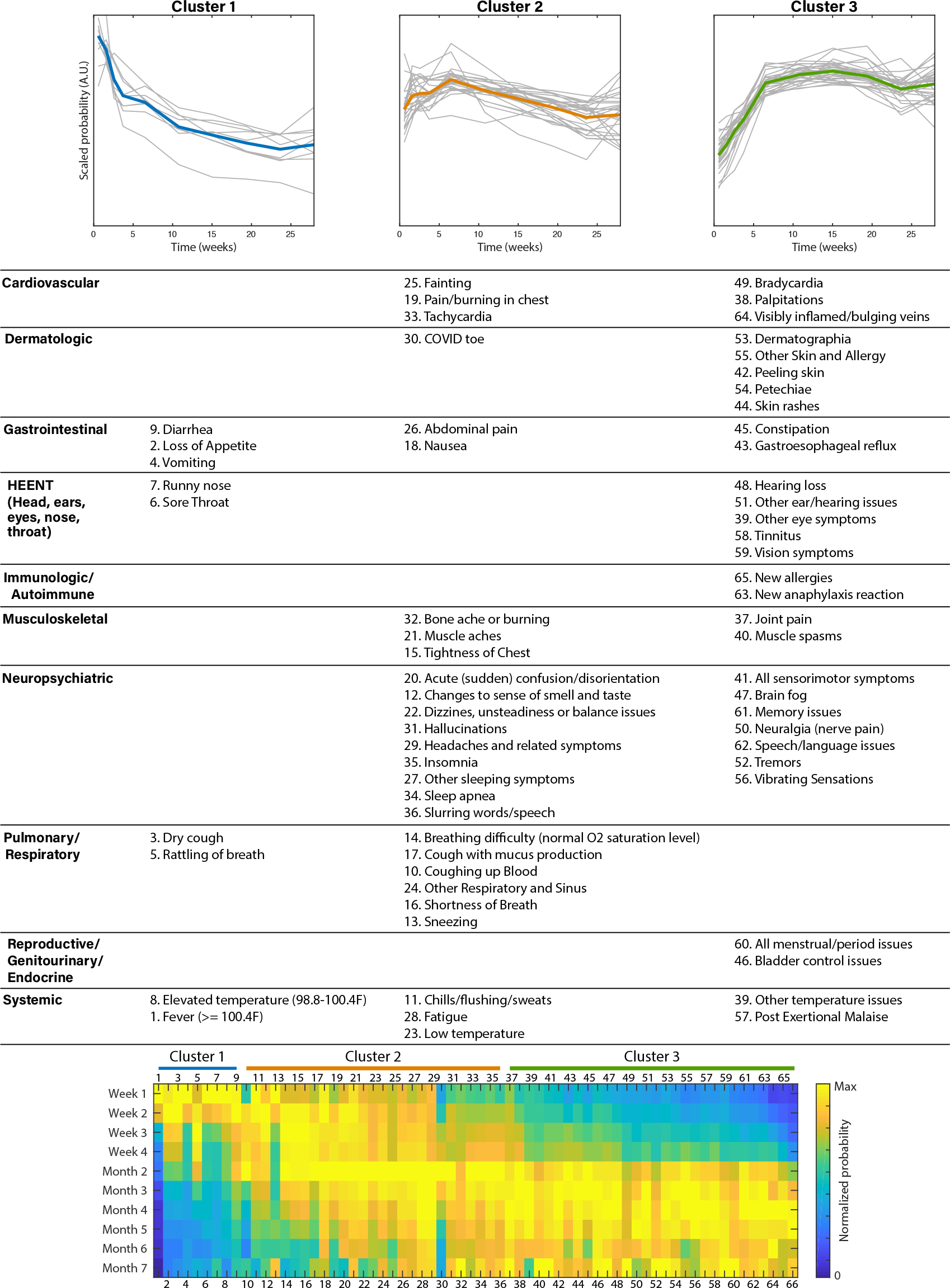
Symptom clusters, based on temporal similarities. Plots (top row) show time courses for the symptoms in each cluster (in grey) and their mean (Cluster 1 in blue, Cluster 2 in orange, Cluster 3 in green). Time courses have been scaled separately for each symptom (by root mean squared amplitude) to visually compare their shapes. The table lists symptoms in each cluster, grouped by the affected organ systems. The heatmap (bottom row) shows time courses for all symptoms, sorted such that similarly shaped time courses are adjacent (see Methods). Columns have been scaled by their maximum amplitudes for visual comparison. Symptoms are numbered according to their table entries.

Symptom prevalence plots, together with the onset times and clusters (Figures 2-6) show that experienced symptoms affect multiple organ systems. The mean number of organ systems affected in each respondent was 9.08 (95% confidence interval 9.04 to 9.13; see Symptom Details). Symptoms in the same organ-based category did not necessarily cluster together, and could appear across clusters. This indicates that symptoms affecting the same organ system can have differently shaped time courses and, conversely, symptoms affecting different organ systems can have similarly shaped time courses. Systemic and neurological/cognitive symptoms were the most likely to persist from disease onset to month 7 (see Symptom Details).

#### Symptoms by test result

Among respondents who received a diagnostic test (RT-PCR or antigen) for SARS-CoV-2 at any point during their illness, 1,730 tested negative and 600 tested positive. The primary difference between these two groups was the time elapsed between symptom onset and testing, with a median of 6 days for those who tested positive and 43 days for those who tested negative (p < 0.001, Mann-Whitney U test) (Supplemental Figure S6). Symptoms were remarkably similar between the two groups. We compared symptom prevalence among positively and negatively tested respondents, stratified by test time. Out of 205 symptoms, 203 showed no significant difference at the 5% level (Fisher test, Bonferroni corrected). The loss of smell and taste were the only exceptions (loss of smell: 22.2% (negative) vs 60.8% (positive), p < 0.0001; 21.5% loss of taste: 21.5% (negative) vs. 54.9% (positive), p < 0.0001; Fisher test, Bonferroni corrected). In addition, 683 participants tested positive for SARS-CoV-2 antibodies (either IgG, IgM, or both). Similarly, the loss of smell and taste were the only significantly different symptoms when comparing prevalence among respondents who tested negative (diagnostic *and* antibody, 21.6% loss of smell, 25.3% loss of taste) versus positive (diagnostic *or* antibody, 60.0% loss of smell, 52.5% loss of taste), stratified by test time (p < 0.0001, Fisher test, Bonferroni corrected).

Furthermore, respondents experienced similar variation in symptoms over time, despite differences in testing status. For 65 out of 66 symptoms, time courses overlapped substantially between participants with confirmed COVID-19 (n=1,020, positive RT-PCR, antigen, or antibody test at any point) and participants with no positive test result (n=2,742, Figure 7). As above, change in smell/taste was the lone exception. Similar overlap was observed when separately comparing positively tested participants to negatively tested and untested participants (Supplemental Figures S7 and S8).

**Figure 7.**
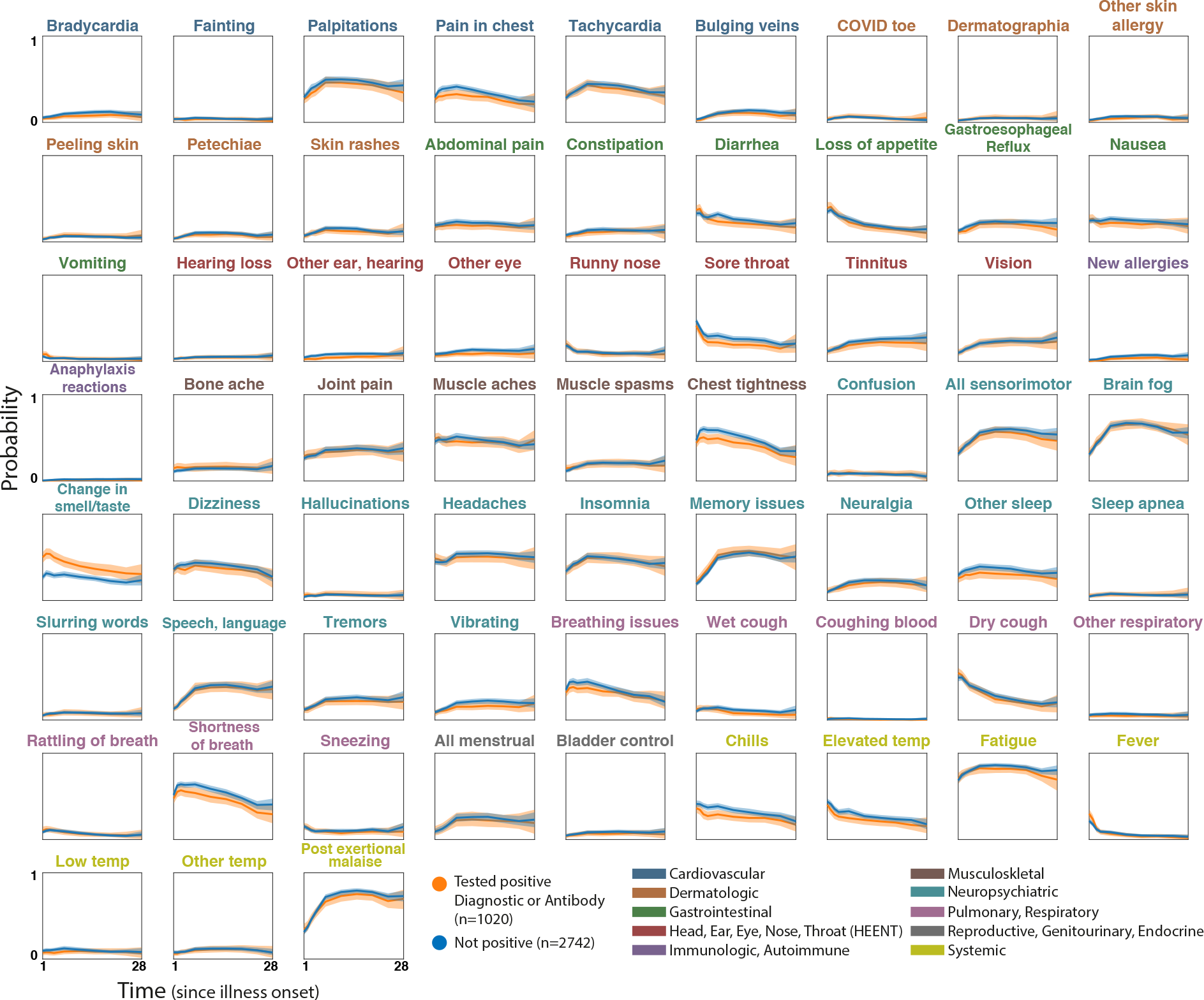
Symptom time courses for participants with COVID-19 confirmed via testing vs. rest of the population. Plots show symptom time courses (similar to Fig. 4) for respondents who were confirmed COVID-positive via diagnostic or antibody testing (orange) vs those without a positive confirmation (untested, or tested negative, blue). Shaded regions show simultaneous 95% confidence bands. Symptom names are colored according to the affected organ systems.

### Symptom Details

#### Neuropsychiatric

##### Brain fog/Cognitive dysfunction and memory impairment

85.1% (95% confidence interval 83.9% to 86.2%) of respondents (3203) reported experiencing brain fog and cognitive dysfunction (Figure 3d, Supplemental Table S13). The most common symptoms were poor attention or concentration 74.8% (73.4% to 76.2%), difficulty thinking 64.9% (63.4% to 66.4%), difficulty with executive functioning (planning, organizing, figuring out the sequence of actions, abstracting) 57.6% (56.0% to 59.1%), difficulty problem-solving or decision-making 54.1% (52.4% to 55.6%), and slowed thoughts (49.1%, 40.2% to 43.4%). For 31.2% (29.7% to 32.7%) of respondents, onset of brain fog/cognitive dysfunction occurred in their first week of symptoms. Reports of cognitive dysfunction increased over the first three months, peaking at 66.7% (65.1% to 68.2%), then decreased slightly in the following months. 55.5% (52.5% to 58.8%) of month 7 respondents experienced cognitive dysfunction during month 7 (Figure 4j).

72.8% (71.4% to 74.2%) of all respondents (2739) experienced memory impairments (Figure 3e, Supplemental Table S14). Of those, 64.8% (63.3% to 66.4%) experienced short-term memory loss, 36.12% (34.6% to 37.6%) experienced long-term memory loss, 12.0% (11.0% to 13.1%) forgot how to do routine tasks, and 7.3% (6.5% to 8.2%) were unable to make new long-term memories. The likelihood of experiencing memory symptoms increased the first few months, with 55.9% (54.3% to 57.5%) reporting memory symptoms in month 4. 50.5% (47.3% to 53.6%) of respondents with symptoms for over 6 months experienced memory symptoms in month 7 (also Figure 4j).

Of those who experienced memory and/or cognitive dysfunction symptoms and had a brain MRI, 87% of the brain MRIs (n=345, of 397 who were tested) came back without abnormalities.

##### Impact of cognitive dysfunction/memory on daily abilities and impact by age

88.0% of the total respondents (3310) experienced either cognitive dysfunction or memory loss (Figure 8). The greatest area of impact reported was on work, with 86.2% (95% confidence interval 84.4 to 88.0%) of working respondents feeling mildly to severely unable to work - 29.1% (26.7% to 31.6%) severely. This is reflected in the working status of respondents, discussed in the Impact on Work section below. Other areas of impact included making serious decisions 85.3% (80.7% to 89.8%), communicating thoughts and needs 74.8% (72.5% to 77.1%), having conversations with others 68.3% (65.8% to 70.8%), maintaining medication schedules 62.5% (59.8% to 65.1%), following simple instructions 54.4% (51.6% to 57.2%), and driving 53.2% (50.5% to 56.0%). See Figure 8d for the full list.

**Figure 8.**
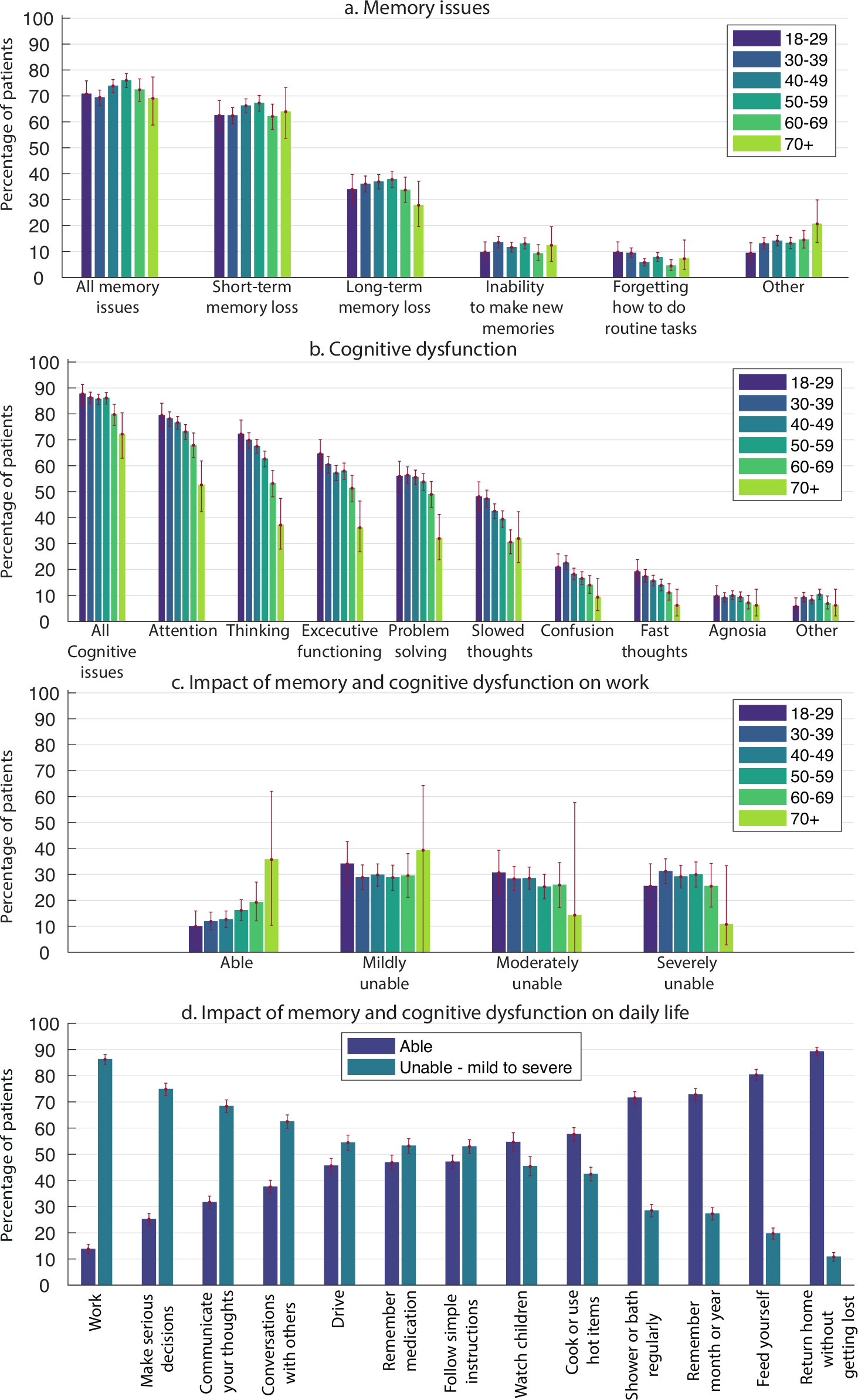
Memory and cognitive dysfunction. a) Percentage of respondents in six age groups who experienced different types of memory impairments. b) Same as (a) for cognitive dysfunction. c) Impact of memory and cognitive dysfunction on work (for those who work), for different age groups. Participants were asked to rate the impact by choosing one of the four options “Able, Mildly unable, Moderately unable, and Severely unable”. d) Overall impact of memory and cognitive dysfunction on daily life. Participants to whom the question was not applicable were excluded. Error bars show 95% confidence intervals.

Memory symptoms, cognitive dysfunction, and the impact of these on daily life were experienced at the same frequency across all age groups (Figure 8a-c). Selected quotes from respondents who described specific instances of memory loss or brain fog can be found in Appendix B.

##### Speech and language

Speech and language issues occurred in 48.6% (95% confidence interval 47.0% to 50.2%, Supplemental Table S15) of respondents (Figure 3h). The most common speech/language symptom was word retrieval, with 46.3% (44.8% to 47.9%) of respondents having difficulty finding words while speaking or writing. 29.2% (27.8% to 30.7%) of respondents had difficulty communicating verbally, 24.8% (23.3% to 26.1%) had difficulty reading/processing written text and 23.8% (22.5% to 25.2%) had difficulty processing/understanding others. 28.9% (27.1% to 23.6%) of those who spoke two or more languages had changes to their non-primary language. Speech and language symptoms occurred in 13.0% (12.0% to 14.1%) of respondents in the first week, increasing to 40.1% (38.5% to 41.7%) experiencing these issues in month 4. 38.0% (34.5% to 41.0%) of respondents with symptoms for over 6 months reported speech and language symptoms in month 7 (Figure 4k).

##### Sensorimotor symptoms

Sensorimotor symptoms encompass a collection of symptoms: tremors, “vibrating sensation”, numbness, coldness in a body part, tingling/pins and needles, “electric zap,” facial paralysis, facial pressure/numbness, and weakness (Figure 3b). These were experienced by 80.5% (95% confidence interval 79.3% to 81.8%) of respondents, occurring in 32.0% (30.5% to 33.4%) of week 1 respondents and increasing to 59% (57.5% to 60.7%) of month 4 respondents (Figure 4k). Tingling, prickling, and/or pins and needles was the most common at 49% (47.7% to 50.8%) of respondents. See Supplemental Table S3, and Table S16 for the most commonly affected anatomical locations.

##### Sleep

78.6% (95% confidence interval 84.0% to 79.9%) of respondents experienced difficulty with sleep (Figure 3c, Supplemental Table S17). Table 5 lists each type of sleep symptom, as well as the percentage of respondents with that symptom who also listed it as pre-existing (before COVID-19 infection).

**Table 5.** Prevalence of sleeping issues before and during illness.

| Sleep Symptom | Experienced During Illness* | Pre-existing Issue** |
| --- | --- | --- |
| Insomnia | 60% (67.1 to 70.1%) | 21% |
| Night Sweats | 41% (39.2 to 42.4%) | 16% |
| Awakened Feeling Unable to Breathe | 36% (34.5 to 37.6%) | N/A |
| Restless Legs | 18% (16.6 to 19%) | 14% |
| Sleep Apnea | 10% (9.5 to 12.8%) | 34% |
| Vivid Dreams | 33% (31.5 to 34.5%) | 23% |
| Nightmares | 26% (24.3 to 27.1%) | 20% |
| Lucid dreams | 15% (14.2 to 16.6%) | 34% |

\*Of all respondents
\*\*Of those who experienced the symptom

##### Headaches

Headaches were reported by 77.0% of participants (95% confidence interval 75.4% to 78.0%, Supplemental Table S18), with the most common manifestations being ocular 40.9% (38.6 to 41.7%), diffuse 35.0% (33.6% to 36.7%), and temporal 34.0% (32.4% to 35.5%) (Figure 3f). 24.0% (22.5% to 25.2%) of respondents reported headaches after thinking/mental exertion and 23.0% (21.9% to 24.6%) experienced migraines. Of those experiencing migraines, 56.4% did not list migraines as a pre-existing condition. 46% of all respondents reported headaches during week 1, 54% of respondents experiencing symptoms in month 4 reported headaches in month 4, and 50% of respondents experiencing symptoms in month 7 reported headaches in month 7 (Figure 4l).

##### Emotion and mood

Changes to emotion and mood were reported by 88.3% (95% confidence interval 87.2% to 89.3%Supplemental Table S19) of participants (Figure 3a). Anxiety was the most common psychological symptom reported at 57.9% (56.4% to 59.5%), followed by irritability at 51.0% (49.5% to 52.7%). Depression was reported by 47.3% (45.7% to 48.9%) with 39.2% (37.6% to 40.7%) experiencing apathy. Mood lability, assessed by “mood swings” and “difficulty controlling emotions,” was reported by 46.3% (37.6% to 40.7%). Suicidality was reported by 11.6% (10.6% to 12.6%), and mania and hypomania were reported at 2.6% (2.1% to 3.1%) and 3.4% (2.8% to 4.0%), respectively. Of those who reported anxiety, 61.4% (59.4% to 63.4%) had no anxiety disorder prior to COVID. Of those who reported depression, 53.12% (50.8% to 55.4%) had no depressive disorder prior to COVID.

##### Taste and smell

Changes to taste and smell (Figure 3g) were reported by 57.6% (95% confidence interval 56.0% to 59.2%, Supplemental Table S20), with no significant difference seen in loss of smell (35.9%, 34.4% to 37.5%) vs. loss of taste (33.7%, 32.2% to 35.2%, p > 0.1, chi-squared test). Altered sense of taste was experienced by 25.1% (23.7% to 26.4%) of respondents, phantom smells (i.e. olfactory hallucinations or phantosmia) by 23.2% (21.9% to 24.6%) of respondents, and altered sense of smell by19.8% (18.5% to 21.1%) of respondents. Phantom smells were accompanied by a write-in question asking for a description of the smells, in which the most common words were “smoke,” “burning,” “cigarette,” and “meat.”

Changes to smell and taste were more likely to occur earlier in the illness course, with 33.2% occuring in week 1. 25.2% (22.5% to 28%) of respondents with symptoms for over 6 months experienced changes to taste and smell in month 7 (Figure 4k).

##### Hallucinations

The most common hallucination reported was olfactory hallucinations 23.2% (21.9% to 24.6%, Supplemental Table S21), mentioned above (Figure 3i). Visual hallucinations were reported by 10.4% (9.5% to 11.4%) of respondents, auditory hallucinations by 6.5% (5.7% to 7.3%), and tactile hallucinations by 3.1% (2.6% to 3.7%).

#### Systemic

Fatigue (98.3%, 95% confidence interval 97.9% to 98.7%) and post-exertional malaise (PEM) 89.0% (88.0% to 90.0%) were the most common symptoms reported by respondents (Figure 2a, Supplemental Table S4), as reported previously [18]. Each increased in likelihood over the first two months of illness before plateauing (Figure 4a). Weakness was experienced by 44.5% (42.9% to 46.1%) of respondents.

Elevated temperature below 100.4F (58.2%, 56.5% to 59.8%) was almost twice as common as fever above 100.4F (30.8%, 29.3% to 32.3%). 3.0% (2.5% to 3.7%) experienced a continuous fever (>100.4F) for 3 or more months, and 15.0% (13.8% to 16.1%) experienced an elevated temperature, continuously, for 3 or more months.

Skin sensations of burning, itching, or tingling without a rash were reported by 47.8% (45.3% to 48.5%) of respondents.

#### Reproductive/Genitourinary/Endocrine

Total of 2979 respondents reported that the question “If applicable, do you have periods/a menstrual cycle” applied to them by responding either *Yes*, *No - Post-Menopausal*, or *No - Other*. Of the 1792 respondents who reported having periods/a menstrual cycle, 36.1% (95% confidence interval 33.8% to 38.3%) reported experiencing menstrual/period issues. For this group, these issues included abnormally irregular periods (26.1%, 24.0% to 28.2%, Figure 2b, Supplemental Table S5) and abnormally heavy periods/clotting (19.7%, 18.0% to 21.6%). Of the 1123 cis women respondents over 49, 4.5% (3.46% to 5.85%) experienced post-menopausal bleeding/spotting. Of the 938 cis women respondents in their 40s, 3.0% (2.0% to 4.3%) experienced early menopause.

Sexual dysfunction occurred across genders, experienced by 14.6% (95% confidence interval 12.1% to 17.4%) of men (cis or trans) respondents, 7.9% (7.0% to 9.0%) of women (cis or trans) respondents, and 15.87% (7.94% to 26.9%) out of 63 nonbinary respondents.

Pain in testicles was reported by 10.9% (8.6% to 13.2%) of the 714 cis men participants.

Extreme thirst was reported by 35.8% (34.3% to 37.3%) of respondents. Bladder control issues were experienced by 14.1% (13.1% to 15.3%) of respondents. Bladder control issues were not particularly variable over time (Figure 4b), while menstrual issues ramped up in prevalence over the first two months.

#### Cardiovascular

86% (95% confidence interval 84.9% to 87.2%) of respondents reported experiencing cardiovascular symptoms (Figure 2c, Supplemental Table S6). The most commonly reported symptoms were heart palpitations (67.4%, 65.9% to 68.8%), tachycardia (61.4%, 59.8% to 62.9%), and pain/burning in the chest (53.1%, 51.5% to 54.7%). Fainting was experienced by 12.9% (11.9% to 14%) of respondents.

**Table 6.** Test results for latent disease.

| Virus | Positive* | Positive (past) | Negative | Total Tested |
| --- | --- | --- | --- | --- |
| Epstein-Barr | 40 | 309 | 231 | 580 |
| Lyme Disease | 7 | 34 | 366 | 407 |
| CMV | 4 | 85 | 204 | 293 |

\* Includes both current and recent cases

Cardiovascular symptoms were more common over the first 2 months than in later months (Figure 4c). Even so, 40.1% (37.9% to 44.1%) of respondents with symptoms for over 6 months experienced heart palpitations, 33.7% (30.8% to 36.8%) experienced tachycardia, and 23.7% (20.7% to 26.0%) experienced pain/burning in the chest in month 7.

##### Postural Orthostatic Tachycardia Syndrome (POTS)

To assess the possibility of POTS, participants were asked whether they had the ability to measure their heart rate, if their heart rate changed based upon posture, and if standing resulted in an increase of over 30 BPM [19]. Of the 2,308 patients who reported tachycardia, 72.8% (n=1680) reported being able to measure their heart rate. Of those, 30.65% (n=515) reported an increase in heart rate of at least 30 BPM on standing.

#### Musculoskeletal

Musculoskeletal symptoms were common in this cohort, seen in 93.9% (95% confidence 93.0% to 94.6%) (Figure 2d, Supplemental Table S7). Chest tightness was most common (74.8%, 73.4% to 76.1%), followed by muscle aches (69.1%, 67.6% to 70.6%) and joint pain (52.2%, 50.5% to 53.8%). In month 7, chest tightness affected 32.9% (29.9% to 36.0%) of month 7 respondents and muscle aches affected 43.7% (40.6% to 46.9%) of month 7 respondents (Figure 4d).

#### Immunologic and Autoimmune

Immunologic and autoimmune symptoms were reported by 21.0% (95% confidence interval 19.8% to 22.4%) of respondents (Figure 2e, Supplemental Table S8). Heightened reaction to old allergies was most common, at 12.1% (11.0% to 13.1%), followed by new allergies at 9.3% (8.4% to 10.2%). New or unexpected anaphylaxis reactions were notable at 4.1% (3.5% to 4.7%). Change in prevalence over time was not notable (Figure 4e).

20.3% of respondents (n=765) reported experiencing changes in sensitivity to medications.

##### Reactivation and test results for latent disease

Since being infected with SARS-CoV-2, 2.8% (2.3% to 3.3%) of respondents reported experiencing shingles (varicella zoster reactivation), 6.9% reported current/recent EBV infection, 1.7% reported current/recent Lyme infection, and 1.4% reported current/recent CMV infection. Detailed results are shown in Table 6.

#### HEENT (Head, ears, eyes, nose, throat)

28 symptoms were defined as symptoms of the head, ears, eyes, nose, and throat (HEENT) (Figure 2f, Supplemental Table S9). All respondents experienced at least one HEENT symptom.

Sore throat was the most prevalent symptom (59.5%, 95% confidence interval 57.9% to 61.1%) which was reported almost twice as often as the next most prevalent symptom, blurred vision (35.7%, 34.2% to 37.3%). Within this category, symptoms involving vision were as common as other organs. Notably, 1.0% (0.7% to 1.4%) of participants reported total loss of vision (no data on the extension and duration of vision loss were collected).

Ear and hearing issues (including hearing loss), other eye issues, and tinnitus became more common over the duration studied (Figure 4f). Tinnitus, for example, increased from 11.5% (10.5% to 12.5%) of all respondents reporting it in week 1 to 26.2% (23.5% to 29.1%) of respondents with symptoms for over 6 months reporting it in month 7.

#### Pulmonary and Respiratory

93.0% (95% confidence interval 92.2% to 93.8%) of respondents reported pulmonary and respiratory symptoms (Figure 2g, Supplemental Table S10). Shortness of breath at 77.4% (76.1% to 78.8%) was more common than dry cough at 66.2% (64.7% to 67.7%) or breathing difficulty with normal oxygen levels at 60.4% (58.8% to 61.9%). Rattling of breath was reported by 17.0% (15.8% to 18.3%) of respondents.

Dry cough was reported by half of respondents in week 1 (50.6%, 49.0% to 52.5%) and week 2 (50.0%, 48.4% to 51.6%), and decreased to 20.1% (17.8% to 22.8%) of respondents with symptoms for over 6 months in month 7 (Figure 4g). Shortness of breath and breathing difficulties with normal oxygen increased from week 1 to week 2 and had relatively slow decline after month 2. Shortness of breath remained prevalent in 37.9% of respondents (34.8% to 41.0%) with symptoms in month 7 (Figure 10a).

#### Gastrointestinal

Gastrointestinal symptoms (Figure 2h, Supplemental Table S11) were reported at 85.5% (95% confidence interval 84.4% to 86.6%) overall. Diarrhea was the most commonly reported gastrointestinal symptom, experienced by 59.7% (58.1% to 61.3%) of respondents, followed by loss of appetite (51.6%, 50.0% to 53.2%) and nausea (47.8%, 46.2% to 49.4%). Of respondents experiencing symptoms after month 6, 20.5% (18.1% to 23.2%) reported diarrhea and 13.7% (11.6% to 16.0%) reported loss of appetite in month 7 as shown in Figure 4h.

#### Dermatologic

As shown in Figure 2i (Supplemental Table S12), dermatological symptoms were present in 59.1% (95% confidence interval 57.5% to 60.6%) of respondents. Itchy skin (31.2%, 29.7% to 32.6%) and skin rashes (27.8%, 26.3% to 29.2%) were most common. 17.8% (16.6% to 19.1%) of respondents reported petechiae, while COVID toe was reported by 13.0% (12.0% to 14.1%) of respondents. COVID toe, petechiae, and skin rashes were most likely to be reported in months 2 through 4 and decreased thereafter (Figure 4i).

#### Post-exertional malaise

The survey asked participants whether they have experienced “worsening or relapse of symptoms after physical or mental activity during COVID-19 recovery” [11]. Borrowing from Myalgic Encephalomyelitis/Chronic Fatigue Syndrome (ME/CFS) terminology [20], this is referred to as post-exertional malaise (PEM). 89.1% of participants (95% confidence interval 88.0% to 90.0%) reported experiencing either physical or mental PEM.

Of the respondents who experience PEM triggered by physical exertion, 49.6% (48.0% to 51.3%) experience it the following day, 42.5% (40.8% to 44.2%) experience it the same day, and 28.7% (28.3% to 31.3%) experience PEM immediately after (Figure 9). Of the respondents who experience PEM triggered by mental exertion, 42.2% (40.5% to 43.8%) experience it the same day, and 31.4% (29.9% to 33.0%) experience it immediately after. For some respondents the time PEM started varied. A high number of the respondents with PEM (68.3%, 66.4% to 69.6%) indicated that the PEM lasted for a few days. For physical exertion, the mean severity rating was 7.71, and for mental exertion, the mean severity rating was 5.47.

**Figure 9.**
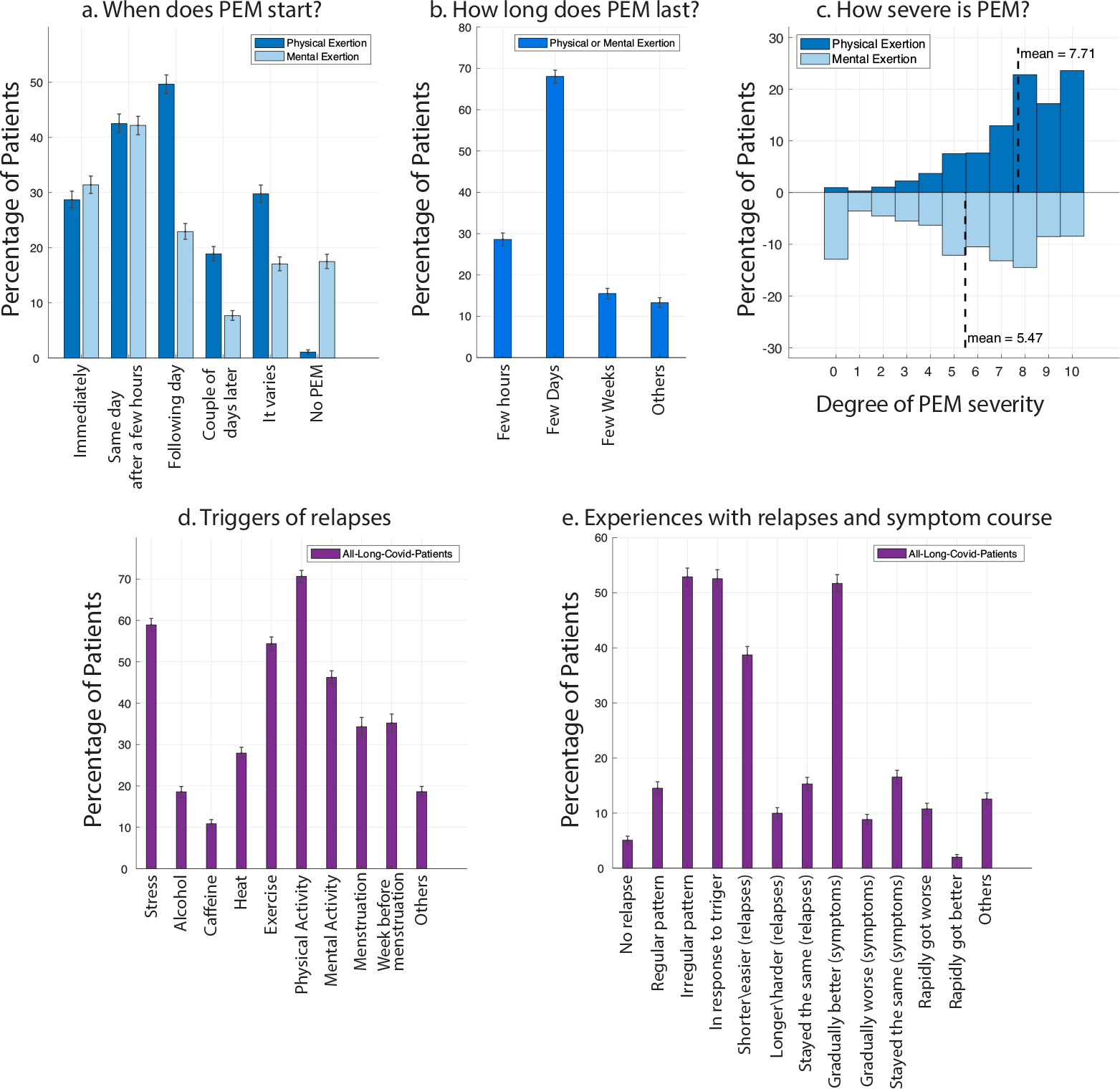
Worsening or relapse of symptoms after physical or mental activity (post-exertional malaise). Worsening of symptoms or relapse after physical or mental exertion. When does it start (a), how long does it last (b), and how severe is it? (c) (all patients who experienced PEM, n=3350). d-e: What are the triggers for relapses/worsening of symptoms and the experience of symptoms over time and relapses?

#### Three most debilitating symptoms

Participants were asked to list the top three to five most debilitating symptoms they have had over the course of their illness. **The top three most debilitating symptoms listed by patients were:** 1) fatigue (n>2652), 2) breathing issues (n>2242), and 3) cognitive dysfunction (n>1274).

## Recovery, return to baseline

### Relapses: triggers & experience

Patients with Long COVID can experience relapsing-remitting symptoms [5]. Minimum of 85.9% (84.8% to 87.0%) of respondents reported experiencing relapses. Respondents characterized their relapses as occurring in an irregular pattern (52.8%, 95% confidence interval 51.2% to 54.4%) and in response to a specific trigger (52.4%, 50.8% to 54.0%). The most common triggers of relapses, or of general worsening of symptoms, that respondents reported were physical activity (70.7%, 69.2% to 72.1%), stress (58.9%, 57.3% to 60.5%), exercise (54.39%, 52.8% to 56.0%), and mental activity (46.2%, 44.7% to 47.8%). More than a third of menstruating participants experienced relapses during (34.3%, 32.0% to 36.5%) or before menstruation (35.2%, 33.0% to 37.3%). Heat and alcohol were other triggers of relapse.

Triggers that were written in by respondents included food with sugar and high histamines (reported by 70 respondents); lack of sleep or rest (64 respondents); cold air (39 respondents); overworking or schoolwork (28 respondents); smoke, pollution, and chemical odors (24 respondents);

Approximately half (51.7%, 50.1% to 53.3%) of respondents indicated that their symptoms have slowly improved over time, while 8.9% (7.9% to 9.8%) indicated that their symptoms have gradually worsened and 10.8% (9.9% to 11.8%) have had symptoms rapidly worsen over time.

### Remaining symptoms after 6 months

Only 164 out of 3762 participants (4.4%) experienced a temporary break in symptoms (Figure S4). The remaining participants reported symptoms continuously, until symptom resolution or up to taking the survey. A total of 2454 (65.2%) respondents were experiencing symptoms for at least 6 months. For this population, the top remaining symptoms after 6 months were primarily a combination of systemic and neurological symptoms (Figure 10). Over 50% experienced the following symptoms: fatigue (80.0%, 95% confidence interval 78.5% to 81.6%), post-exertional malaise (73.3%, 71.5% to 75.1%), cognitive dysfunction (58.4%, 56.5% to 60.2%), sensorimotor symptoms (55.7%, 53.7% to 57.6%), headaches (53.6%, 51.5% to 55.5%), and memory issues (51.0%, 49.1% to 53.0%). In addition, between 30%-50% of respondents were experiencing the following symptoms after 6 months of symptoms: insomnia, heart palpitations, muscle aches, shortness of breath, dizziness and balance issues, speech and language issues, joint pain, tachycardia, and other sleep issues.

**Figure 10.**
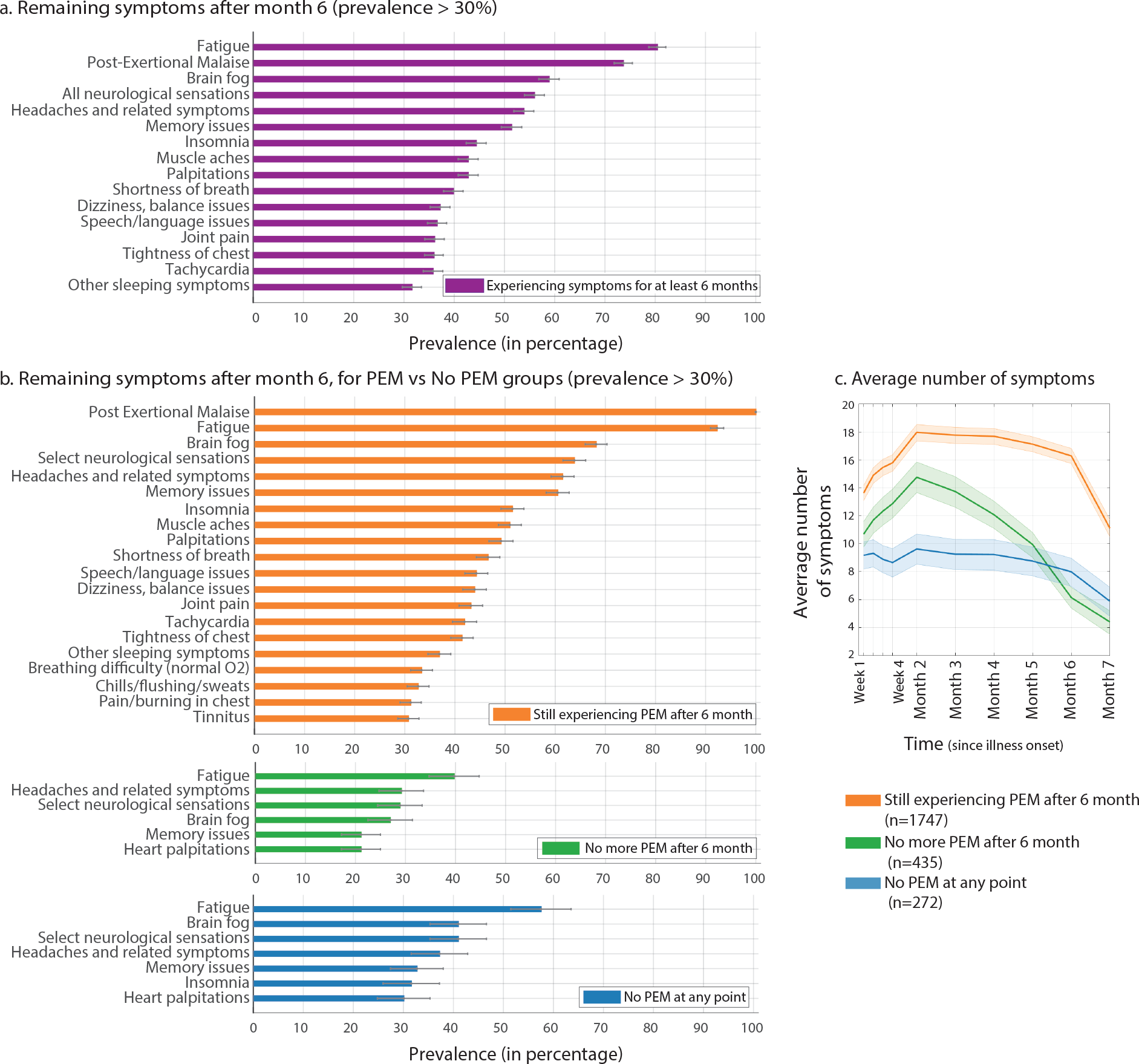
Remaining symptoms after 6 months. a) Symptoms remaining after 6 months. b) Symptoms remaining after 6 months for respondents still experiencing PEM after 6 months (orange), respondents not experiencing PEM after 6 months (green), and respondents who never experienced PEM (blue). c) Average number of symptoms over time for each group in (b).

Respondents were also asked if they had been diagnosed with any number of conditions post-acute COVID-19 infection. Nearly half of respondents (43.4%) responded with at least one common diagnosis and/or elaborated on their diagnosis in free text (see Table S2 Appendix A).

### Fatigue assessment

Participants answered the Fatigue Assessment Scale (FAS) questionnaire [21, 22], which includes ten questions that assess both physical and mental fatigue. FAS scores were calculated based upon participants’ subjective report during the “past one week.” Figure 11.a shows the distribution of FAS scores for the recovered (blue) and unrecovered (orange) participants. The scores were then summarized into three categories (Figure 11.b): no fatigue (scores of 10-21), fatigue (22-34), and extreme fatigue (≥35).

**Figure 11.**
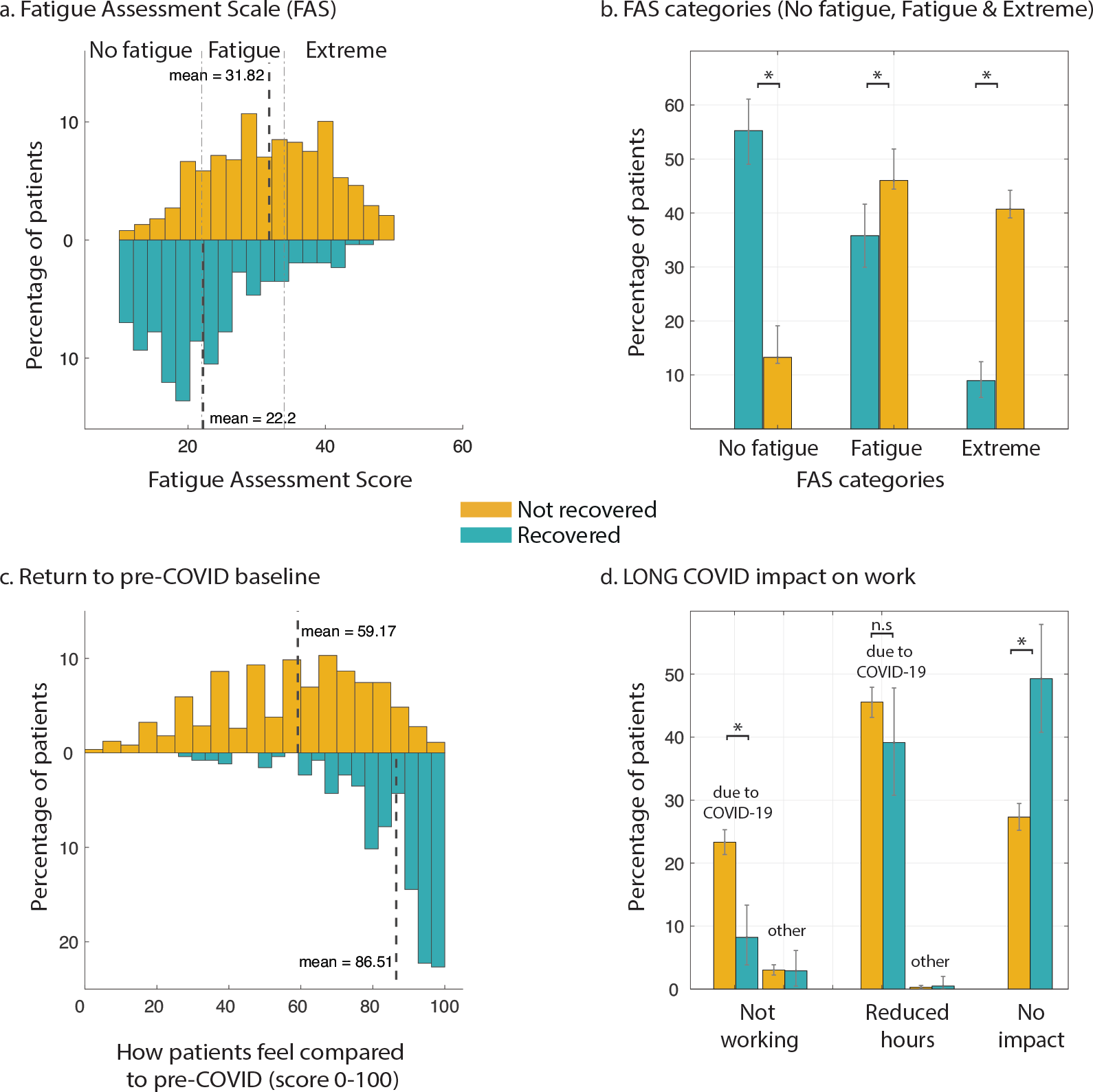
Return to baseline and work impact. a) Distribution of Fatigue Assessment Scale scores for recovered (n=257, blue) and unrecovered (n=3505, yellow) population. The vertical dashed lines indicate the range for “No fatigue” (10-21), “Fatigue” (22-34), and “Extreme” (>=35). Mean values for each distribution are also marked. b) Percentage of participants in each of the three categories. c) Distribution of scores in response to “return to pre-COVID” health baseline, where 0 indicates worst (most different from baseline) and 100 indicates best (most similar to baseline). d) Working status due to COVID-19. Error bars show 95% simultaneous confidence interval.

We contrasted the FAS scored of unrecovered and recovered participants. Of the total of 257 who recovered, respondents experienced symptoms for a mean of 91 (IQR 49-121) and a maximum of 250 days. Those not recovered (n=3505) had experienced 144 days of symptoms on average at the time of the survey (IQR 176-213). On average, unrecovered participants had higher FAS scores than unrecovered participants (31.8 vs 22.2, P < 0.001, Mann-Whitney U-test). 55.3% (95% confidence interval 49.4% to 61.5%) of recovered participants were classified as having no fatigue. This is significantly more than the 13.2% (12.2% to 14.4%, P < 0.001, Mann-Whitney U-test) of unrecovered participants who experienced no fatigue at the time of survey. 40.7% (39.9% to 42.3%) of unrecovered participants were classified as experiencing extreme levels of fatigue, significantly higher than the 8.9% (5.8% to 12.8%) of recovered participants in this category (P < 0.001, Mann-Whitney U-test). 35.8% (30.0% to 41.6%) of recovered and 46.0% (44.4% to 47.7%) of unrecovered participants were classified as experiencing non-extreme fatigue (P < 0.001, Mann-Whitney U-test). FAS scores for the respondents who tested positive for COVID-19 and those who tested negative (with either a diagnostic or antibody test) are similar (P = 0.92, Mann-Whitney U-test, data not shown).

### Return to baseline

Respondents were asked, *“How would you rate how you feel today, on a scale of 0-100% (with 100% being your pre-COVID baseline)?”* (Figure 11c). Unrecovered participants reported a mean score of 59.2, while recovered participants reported a mean score of 86.5 (p<0.001, Mann-Whitney U-test). “Pacing” was the treatment with the highest percentage of respondents considering it ‘significantly helpful’ (23.1% out of 1788 who tried it). 18.8% found it ‘slightly helpful’.

### Impact on work

Of unrecovered respondents who worked before becoming ill, only 27.3% (95% confidence interval 25.3% to 29.4%) were working as many hours as they were prior to becoming ill at the time of survey, compared to 49.3% (40.8% to 57.9%) of recovered respondents (see Figure 11d). Nearly half 45.6% (43.2% to 48.0%) of unrecovered respondents were working reduced hours at the time of the survey, and 23.3% (21.3% to 25.4%) were not working at the time of the survey as a direct result of their illness. This included being on sick leave, disability leave, being fired, quitting, and being unable to find a job that would accommodate them. The remaining respondents retired, were volunteers, or did not provide enough information to determine their working status. Overall, 45.2% (42.9% to 47.2%) of respondents reported requiring a reduced work schedule compared to pre-illness. 22.3% (20.5% to 24.3%) were not working at the time of survey due to their health conditions.

Respondents had the option to elaborate on their situation in free text, and these responses captured the precariousness of working with Long COVID (see selected quotes in Appendix B). Several themes emerged through thematic analysis: At least 45% of working respondents were working remotely at the time of the survey, and it was noted how critical this was to respondents’ continued ability to work. Teleworking enabled respondents to take breaks when necessary and saved them the physical exertion of commuting to work. Respondents mentioned asking for other accommodations at work like flextime or moving to a role with lower physical or mental strain. Even with telecommuting, phased returns, and other accommodations, respondents commented on how difficult it was for them to work full or part-time, but described their financial need to do so.

It is important to note that the survey captured only a moment in time. Respondents described taking months of leave before going back to work either full-time or at reduced hours. Further, there were respondents who indicated that they tried to go back to work for several weeks but then relapsed or were unable to complete their work satisfactorily.

## Discussion

### Principal findings

Results from this international online survey of 3,762 individuals with suspected or confirmed COVID-19 illness suggest that Long COVID is composed of heterogeneous post-acute infection sequelae that often affect multiple organ systems, with impact on functioning and quality of life ranging from mild to severe. A unique patient-led approach allowed for a thorough and systematic identification of possible symptoms based upon reports in online support networks. To our knowledge, this represents the largest collection of symptoms identified in the Long COVID population to date. While several others have investigated Long COVID symptoms [6, 10], our approach also allowed the first representation of individual symptom trajectory over time. The cohort was composed predominantly of individuals with continued symptoms at 6 months. Of the symptoms for which time course data were collected, the most likely early symptoms were fatigue, dry cough, shortness of breath, headaches, muscle aches, chest tightness, and sore throat. Importantly, while presence of fever has widely been used for screening purposes [23–25], we found only 30% of participants reported a fever, consistent with previous reports [24], while the majority did experience some combination of mild elevation in temperature (98.6 - 100.4 F), diaphoresis, temperature lability, and chills.

In this cohort, the most likely symptoms to persist after month 6 were fatigue, post-exertional malaise, cognitive dysfunction (“brain fog”), neurologic sensations (neuralgias, weakness, coldness, electric shock sensations, facial paralysis/pressure/numbness), headaches, memory issues, insomnia, muscle aches, palpitations, shortness of breath, dizziness/balance issues, and speech and language issues. Some symptoms, like bone aches, tinnitus, and other ear symptoms, increased in likelihood during and after month 6. Prolonged symptoms were most likely to be reported as “moderate” (36.6%, 95% confidence interval 32.2% to 40.9%). Notably, the probability of having “severe” or “very severe” symptoms after month 6 was more than 21% (severe: 14.5%, 11.3% to 18.5%; very severe: 5.2%, 3.1% to 7.1%). Respondents indicated that fatigue, breathing issues, and cognitive dysfunction were the most debilitating of symptoms. Those not recovered within three months experienced an average of 13 symptoms during week 1, increasing to 17 symptoms during month 2. They continue to experience an average of 14 symptoms after 6 months.

We propose clusters of symptoms in three groups, each with different morphologies over time. Importantly, the clusters of symptoms that persist longest include a combination of the neurological/cognitive and systemic symptoms. This indicates the need for a multidisciplinary approach to workup and care of the Long COVID population.

Dysautonomia, in part manifesting as Postural Orthostatic Tachycardia Syndrome (POTS), and Myalgic Encephalomyelitis/Chronic Fatigue Syndrome (ME/CFS) appear as highly possible diagnoses for this population [26]. By the time respondents took the survey, 155 had received a diagnosis of POTS, and 118 had received a diagnosis of ME/CFS. Based on the reported symptoms, higher percentages of respondents might meet the criteria for these diagnoses than have been clinically diagnosed. 33.9% of respondents who reported tachycardia measured an increase of at least 30 BPM when standing, suggesting a possible POTS diagnosis [27]. Given these findings, we suggest that all patients who present with any signs or symptoms of POTS, including tachycardia, dizziness, brain fog, or fatigue, be screened for POTS [19].

To investigate the possible overlap with ME/CFS in this population, we asked participants to identify whether they experienced worsening of symptoms after physical or mental exertion. Post-exertional malaise (PEM) is one of the three required symptoms for ME/CFS diagnosis, along with unrefreshing sleep and a reduction in ability to engage in pre-illness levels of activity [28]. Similar to cognitive dysfunction, we found PEM to be highly represented in this cohort (89.1% at any time during the course of illness, 72.2% at month 7). Intriguingly, among those still experiencing symptoms at month 6 with no PEM (n = 707, 28.8%), fatigue was still the most common symptom. This suggests that while a subset of the Long COVID population may meet ME/CFS diagnostic criteria, there remains a subpopulation with significant fatigue who do not meet the criteria, and therefore the mechanisms of fatigue and the degree to which ME/CFS adequately explains it require further investigation.

Participants also experienced symptoms that are not commonly mentioned in public discussion of Long COVID [29, 30], and may benefit from further attention. These include but are not limited to: anaphylaxis and new allergies, seizures, suicidality, changes in sensitivity to medication, vision loss, hearing loss, and facial paralysis. Several of these symptoms, as well as the more commonly reported Long COVID symptoms, overlap with symptoms of Mast Cell Activation Syndrome (MCAS), possibly warranting further exploration into the role of mast cells in Long COVID [31].

This work also highlights the wide range of neurologic symptoms experienced by patients with Long COVID. While respiratory and some cardiovascular symptoms have been widely reported, neurological symptoms remain unclear [32]. Prior studies have identified evidence of cognitive dysfunction induced by COVID-19 illness, with few studies in the non-hospitalized population [26, 33]. Memory and cognitive dysfunction, experienced by over 85% of respondents, were the most pervasive and persisting neurologic symptoms in this cohort, equally common across all ages, and with substantial impact on work. Headaches, insomnia, vertigo, neuralgia, neuropsychiatric changes, tremors, sensitivity to noise and light, hallucinations (olfactory and other), tinnitus, and other sensorimotor symptoms were also all common among respondents, and may point to larger neurological issues involving both the central and peripheral nervous system. This area is particularly important to study, as others have found neurological symptoms may be more common in nonhospitalized patients, and that those with neurological symptoms may have impaired seroconversion [33].

The reduced work capacity because of cognitive dysfunction, in addition to other debilitating symptoms, translated into the loss of hours, jobs, and ability to work relative to pre-illness levels. 68.9% of unrecovered respondents reported reduced work hours or not working at all as a direct result of their COVID-19 illness, and on average the unrecovered group felt they were less than 60% returned to their pre-illness baseline. More than half of recovered respondents, however, still reported being unable to work their pre-COVID hours, as well as being on average only 86.5% back to their pre-illness baseline. Also, only 55.3% of recovered respondents had Fatigue Assessment Scores ranked as “no fatigue”. This could suggest that some respondents who reported that they were no longer experiencing symptoms felt that any lingering effects were part of their new baseline, despite not fully recovering health- or work-wise.

The investigated cohort had a relatively high socioeconomic status which may have skewed results - respondents may have been more likely than the average Long COVID patient to have jobs with sufficient sick leave, have enough savings to sustain them through a period of no or low income, and/or have jobs that were able to offer accommodations. The write-in responses revealed that there were respondents who were working full-time or at reduced hours at the time of the survey, but had taken several weeks, if not months, off of work. For those who returned to their job, respondents reported experiencing relapses triggered by the mental exertion and stress of work, often needing to go back on leave. This emphasizes the importance of all patients having adequate time off to recover, being able to qualify for disability benefits if long-term assistance is needed, and receiving accommodations at work including telecommuting, flexible hours, and phased returns. Lower wage earners may find it especially challenging to access these accommodations and benefits, particularly in locations without robust legal protections for disabled workers, yet they are in need of these protections the most to ensure financial stability [34].

Overall, these findings suggest that the morbidity of COVID-19 illness has been greatly underappreciated. Patients experience multisystem symptoms for over 7 months, resulting in significant impact to patients’ lives and livelihoods.

### SARS-CoV-2 testing

Our analysis confirms prior studies’ results which show that, with the exception of change to smell and taste, symptoms are not significantly different between those who test positive for SARS-CoV-2 and those who test negative (or have not been tested) but who otherwise show strongly suggestive symptoms [5, 35]. The reason for this is not known, though the sensitivity of diagnostic tests may be different depending on the primer/probe sets [36, 37]. Further, the likelihood of false negatives increases after day 3 of symptom onset, when the false negative rate is 20%, reaching 66% by day 21 [38]. This reinforces the need for early testing in patients with suspicion of SARS-CoV-2 infection given that up to 54% of patients could have an initial RT-PCR false-negative result [39]. The importance of early testing was reflected in this cohort as well: the median number of days between first experiencing symptoms and being tested was 6 days for those who tested positive and 43 days for those who tested negative. Access to adequate diagnostic tests in the early stages of the pandemic was notably limited, which likely contributed to respondents in this cohort being unable to be tested and/or being tested later in their illness [40]. The site of sample collection can also play an important role in testing accuracy. Compared to nasopharyngeal swab sampling, sputum testing resulted in significantly higher rates of SARS-CoV-2 RNA detection than oropharyngeal swab testing [41]. Similarly, viral particles may be detected from stool specimens, while respiratory tract specimens are negative by RT-PCR [42]. Regarding antibody testing, it has been reported that antibody levels decrease with time [43, 44]. In a recent study, 96% of participants had reduced antibodies and 28% no longer tested positive for antibodies at the 2 month follow up [43]. There are evidences suggesting that patients with neurological symptoms but minimal respiratory symptoms may fail to seroconvert [33]. These findings indicate that absent or negative SARS-CoV-2 diagnostic and antibody tests should not be used as an indicator to rule out Long COVID in patients who otherwise have suggestive symptoms. Therefore, we included suspected COVID-19 cases in our analysis, as have several other studies [35,45–47]. Further investigations are needed to understand why some Long COVID patients test positive and others do not despite having similar symptom courses.

### Limitations

While the majority of participants did not report receiving a positive SARS-CoV-2 diagnostic or antibody test result, our analysis confirms that this is not a limitation of our study; rather, it is a limitation of the availability and accuracy of SARS-CoV-2 tests.

However, there are several limitations to this study. First, the retrospective nature of the study exposes the possibility of recall bias. Second, as the survey was distributed in online support groups, there exists a sampling bias toward Long COVID patients who joined support groups and were active participants of the groups at the time the survey was published. Additionally, despite eight translations and inclusive outreach efforts, the demographics were strongly skewed towards English speaking (91.9%), white (85.3%), and higher socioeconomic status (see Figure S1). In future studies, more outreach and partnerships with diverse support groups, low-income communities, and communities of color can be established to counter sampling bias. Moreover, the study required respondents to have stable internet and email addresses, which may have excluded participants who lacked access and/or had low digital literacy. Lastly, the effort to complete the survey may have deterred some respondents who experienced cognitive dysfunction, or were no longer ill and did not have enough incentives to participate.

Due to these limitations, we suggest that the results laid forth be considered only in this context, and caution that extrapolation to the entire Long COVID population requires caution.

### Implications

Research by the United Kingdom’s Office of National Statistics estimates that 21% of people who were infected with SARS-CoV-2 still experience symptoms at five weeks, a number which includes asymptomatic patients and will likely be higher for symptomatic patients [48]. Given the millions of cases of COVID-19 worldwide, the prevalence of Long COVID is likely to be substantial, and will only increase as the virus continues to spread. This research demonstrates how expansive and debilitating this prolonged illness can be, with profound impacts to people’s livelihoods and ability to care for themselves and their loved ones. This research demonstrates the importance of slowing the spread of COVID-19 through validated public health measures and vaccinations; a robust safety net including sick leave, disability benefits, and workplace protections and flexibilities for patients, as well as adequate family leave for caregivers; and continued research into Long COVID, ME/CFS, POTS, MCAS, and similar illnesses to determine etiologies and find treatment options. Since Long COVID often impacts multiple organ systems and causes extensive disruption to daily functioning, it is important for medical professionals to consider a multidisciplinary approach to treat and care for patients.

### Future research

This paper is a description of Long COVID from a patient’s perspective - the symptoms; disease course; and impact on daily life, work, and return to baseline health. Valuable future research would emphasize the pathophysiology of Long COVID, answering questions such as: What are the biological underpinnings of Long COVID and what are potential treatments? How is Long COVID impacting the day-to-day life of people, taking into account different socio-economic backgrounds? What can be done to support those with Long COVID, both medically and through policy, until treatments are identified?

Our future work will focus on investigating several emerging topics in Long COVID: mental health outcomes, diagnostic and antibody testing, symptom clustering, patient perspectives on medical attention, and socioeconomic impact from the illness. We look forward to partnering with other research teams for diagnostic data, and driving further collaborations between patient communities, scientists, and clinicians via patient-centered research. We welcome inquiries for analysis by Long COVID patients, particularly patients and community groups of color.

## Data Availability

Based on the UCL Ethics Approval, the raw data cannot be shared publicly. However, anonymised data will become available to all interested parties, after the publication.

## Acknowledgments

We would like to thank the Body Politic COVID-19 Slack Support Group and Admin Team for connecting us to each other and the Long COVID community. We would like to thank them for their incredible help and support in distributing the survey, and connecting us with volunteers. We would like to thank all respondents for their time and energy in contributing to the study and helping to refine the survey. We would like to thank Rachel Robles for her assistance in data cleaning, and Monique Jackson for illustrations used for outreach on social media. We would like to thank Jared Mercier for his help with IT support. We would like to thank the following translators: Oksana Zinchenko (Russian); Emeline Chavernac (French); Maarten Steenhagen and Red Team C19 NL Community (Dutch); Luisa Pereira, Lucía Landa, Maria Teresa Cabañero, Daniel Hernandez Diaz, Brenda Valderrama, and Lorena Ramírez-Nícoles (Spanish); Liliana Vagnoni (Italian); Victor Pedrosa, Monica Malta, and Noris Kern (Portugues); Juno Simorangkir (Indonesian); and Rawan Alsubaie, Sarah Mitkees, Mohamed Abdelhack, Dalia Aroury, Luna Aroury, and Ihsan Kaadan (Arabic). We would also like to acknowledge Dr. Alka Gupta of WCMC for her guidance and involvement in the ethics approval process. We would like to thank the Myalgic Encephalomyelitis community for their advice; And finally, we thank the large Long COVID community and allies for helping to distribute our survey and for providing constant support, including but not limited to Long COVID Support Group, Long Haul Covid Fighters support group, #ApresJ20 (France), Long Covid SOS/Long Covid International, Apuakoronaan (Finland), Pós-Covid-19 (Brazil), Covid Survivor Indonesia, Young Covid Survivors, Black Covid-19 Survivors, COVID Persistente Espana, COVID-19 Vi som är drabbade (Sweden), Long Covid Italia, COVID-19 Persistent Madrid, BIPOC Women Covid Long Hauler Support Group, Long COVID ACTS (Spain), Utah COVID-19 Long Haulers, C19 Longhauler Advocacy Project, JD Davids, Janet Sternberg, Mirror Memoirs, PODER, Bed-Study Strong, Susan Davis, and members of the Reynolds Program in Social Entrepreneurship.

## Author contributions

AA, GSA, HED, LM, YR, and HW conceived the project and designed the survey. HED cleaned the data. AA, GSA, HED, RJL, and LM analyzed the quantitative data. AA and RJL performed the statistical analyses. HED, LM, and HW analyzed the qualitative data. AA and RJL created the figures. HW created the tables. JPA and YR provided medical input. AA, GSA, HED, RJL, LM, SR, YR, and HW wrote the manuscript, with extensive comments from JPA. The corresponding author attests that all listed authors meet authorship criteria and that no others meeting the criteria have been omitted. AA, GSA, HED, RJL, LM, YR, and HW contributed equally to this work. AA is the guarantor.

## Funding

All authors contributed voluntarily to this work. The cost of survey hosting (on Qualtrics) was covered from AA research grant (Wellcome Trust/Gatsby Charity via Sainsbury Wellcome Centre, UCL).

## Competing research statement

All authors have completed the ICMJE uniform disclosure form and declare: no support from any organisation for the submitted work; no financial relationships with any organisations that might have an interest in the submitted work in the previous three years, no other relationships or activities that could appear to have influenced the submitted work.

## Data sharing statement

The data collected for this study, including anonymized individual patient data and a data dictionary defining each field in the data set will be made publicly available. Interested parties can contact the corresponding author (AA).

## Dissemination declaration

We plan to disseminate the results to study participants and or patient organisations.

## Transparency declaration

The corresponding author (AA) affirms that the manuscript is an honest, accurate, and transparent account of the study being reported; that no important aspects of the study have been omitted; and that any discrepancies from the study as planned have been explained.

## Supplemental Material

## Appendix A: Participant profile, symptom information

### Socioeconomic status

3084 (82.0%) participants reported their income at the time of the survey. A majority of participants in the USA, UK, and Canada belong to the middle and upper-middle income brackets, with 51.0% of participants in the USA earning more than $85,000/year and 22.5% earning more than $150,000/year. Meanwhile, 25.0% of survey participants from elsewhere reported less than €20,000/year in income and 51.1% reported earning less than €40,000/year.

### Pre-existing conditions

Most patients (83%) reported at least one pre-existing condition. The most commonly reported pre-existing conditions were seasonal allergies (36.3%), environmental allergies (24.1%), migraines (18.7%), and asthma (17.1%). Other conditions of note include acid reflux (12.2%), irritable bowel syndrome (12.9%), vitamin D deficiency (11.8%), obesity (10.7%), hypertension (9.1%), hyperlipidemia (7.4%), and myalgic encephalomyelitis / chronic fatigue syndrome (2.5%). In the United States, the prevalence of asthma is 7.7%. While this cohort is not representative of the U.S. population, the prevalence of asthma (17.07%) should be noted.

### Post-acute diagnoses

1146 respondents (30.5%) sought a diagnosis after the onset of illness. 802 respondents received one of the following diagnoses listed in the table below. Additionally, 197 respondents received a diagnosis of post-viral fatigue, post-viral syndrome, post-viral inflammation, post-COVID fatigue syndrome, or post-COVID syndrome, pointing to different working diagnoses for Long COVID [3, 7].

In text input, some respondents documented difficulties in receiving a diagnosis, with reasons ranging from a lack of access to specialists, waiting for tests to be scheduled and performed, to healthcare providers dismissing symptoms as anxiety.

### Survey distribution

The majority of respondents came from the Body Politic COVID-19 Slack support group, Long COVID Support group on Facebook, and Long Haul Covid Fighters support group on Facebook. Additionally, the survey was shared with international advocacy groups, including #ApresJ20 (France), Long Covid SOS/Long Covid International, Apuakoronaan (Finland), COVID Persistente Espana, COVID-19 Persistent Madrid, Long COVID ACTS (Spain), and Long Covid Italia. Additional support groups included Pós-Covid-19 (Brazil), Covid Survivor Indonesia, Young Covid Survivors, Black Covid-19 Survivors, COVID-19 Vi som är drabbade (Sweden) BIPOC Women Covid Long Hauler Support Group, Survivor Corps, and others. It was also shared on other social media platforms, including Instagram, Twitter, and Reddit, and with nonprofits and mutual aid organizations. Additionally, it was shared with the Body Politic and Patient-Led Research team email mailing groups reaching over 15,774 contacts.

### Symptom categories

Symptoms are grouped in 10 categories, as below, given which organ/system they manifest in:

- Systemic: fatigue, temperature, weakness, flushing and sweating related symptoms, and post-exertional malaise
- Neuropsychiatric - because of the number and prevalence of neuropsychiatric symptoms assessed, they are broken into the following nine sub-categories and discussed separately:
  - Cognitive Functioning
  - Memory
  - Speech and Language
  - Sensorimotor Symptoms
  - Sleep
  - Headaches
  - Emotion and Mood
  - Taste and Smell
  - Hallucinations
- Cardiovascular: heart rate, palpitations, blood pressure (excluded from in-depth analysis, see Supplemental Figure S5), visibly bulging veins, clots, and pain/burning in the chest
- Dermatologic: itchiness, rashes, and obvious changes in skin and nails
- Gastrointestinal: GI upset, hyperactive bowel sensations, and appetite-related symptoms
- Pulmonary and Respiratory - encompasses breathing, coughing, and sneezing, and oxygen saturation related symptoms (excluded from in-depth analysis, see Supplemental Figure S5)
- Head, Ear, Eye, Nose, Throat (HEENT): both physical and sensory symptoms related to the eyes, ears, nose, mouth, throat, and face including facial paralysis and numbness. Headaches are captured in the Neuropsychiatric category.
- Reproductive, Genitourinary, and Endocrine: symptoms related to menstruation and lack thereof, symptoms related to male reproductive function, symptoms related to sexual function, symptoms related to thirst and urinary function, and low and high blood sugar (excluded from in-depth analysis, see Supplemental Figure S5)
- Immunologic and Autoimmune: new and heightened immune responses
- Musculoskeletal: chest tightness and aches and pain throughout the musculoskeletal system

### Sensorimotor symptoms - impacted body parts

For each of these symptoms, we asked participants to write in the part(s) of the body that was affected, and performed natural language processing to identify the top four locations affected for each symptom.

## Appendix B: Quotes from Participants

#### Cognitive Dysfunction and Memory Loss

*“mother has started to help me take the medications I’m on because I **can’t remember** if I’ve taken them immediately after having the bottle in my hand”*

*“was trying to fill out a mortgage application form and couldn’t remember our rent. I put £3750 a month. My partner said, no it’s £1375. So I put £13750. My partner said no, so I tried several more times - I **was just guessing numbers**”*

*“sitting on the toilet to pee and **had to stop for a second to think** if I was really there and not about to pee myself or the bed”*

*“**don’t remember what I did** in March or April up until the last week of April. I had almost nothing on my schedule. I don’t know what I did”*

*“put food on the gas stove and walked away for over an hour, **only noticing when they were smoking/burning**”*

*“**forget how to do normal routines** like running a meeting at work”*

*“**felt lost driving** and had to stop and find my position in a GPS to be able to drive back home. It’s a route I have done hundreds of times”*

*“have trouble **comprehending new ideas**”*

*“**can’t hold multiple trains of thought** […] If I tell myself I have to water my plants, I must do it before another thought comes into my mind because otherwise I will forget”*

*“**can’t follow plots** in movies or tv shows, **have to write everything down**, have to remember to look at notes”*

*“had to terminate many phone calls because I **could no longer comprehend the speakers nor communicate clearly** with them”*

*“used to do the New York Times crossword puzzle every single day and I **can’t even manage the mini ones** now”*

*“**can’t focus on reading complex texts**, and it makes me feel very tired to do that”*

*“Found that I had become dyslexic - and knew it was happening at the time, **could not remember how to spell words** - also found I was missing words from sentences and sometimes writing things that did not make sense”*

#### Impact on Work

*“I **worked at some point for a few weeks, in June, but had to stop** (couldn’t handle a conversation on the phone without brain fog / feeling dizzy / heavy breath trouble because of talking) after a few minutes”*

*“Haven’t been able to work for […] months due to brain fog. **Was supposed to go back last week on reduced hours. I resigned instead.** I have worked there as Director of […] for just over […] years.”*

*“Still on medical leave. Unpaid and **denied short term disability**.”*

*“I **went from [being] a workaholic to no workaholic at all**. This is the extreme opposite of who I am. […] I do not know the person I have become.”*

*“I **went back to work too soon and wish I hadn’t. Finally had to take a 5 week break** in July/ August with the support of my employer. This helped a lot. I have now been back at work for 5 weeks and my symptoms have got worse to a degree.”*

*“I had to take two weeks off, had to work from home for four, but had to return for two weeks with fever as **my employer would not give me more time** […].”*

*“I **asked to reduce hours** or work more from home to which **it was denied**.”*

*“I’ve been working from home. Haven’t officially reduced my hours, but **my boss [has] been flexible and encouraged me to rest when needed**.”*

*“While **I’ve been able to keep my job while working from home**, I must admit that if it were not so, I would most definitely NOT be able to work at all. **I can barely leave my bedroom on most days**.”*

*“I **have needed more-flexible hours** (working remotely) post-COVID. That way, **I can rest as needed throughout the day**. If I had to return to in-person work at this point, it would be severely reduced hours if at all.”*

## Appendix C: Supporting Figures

**Figure S1.**
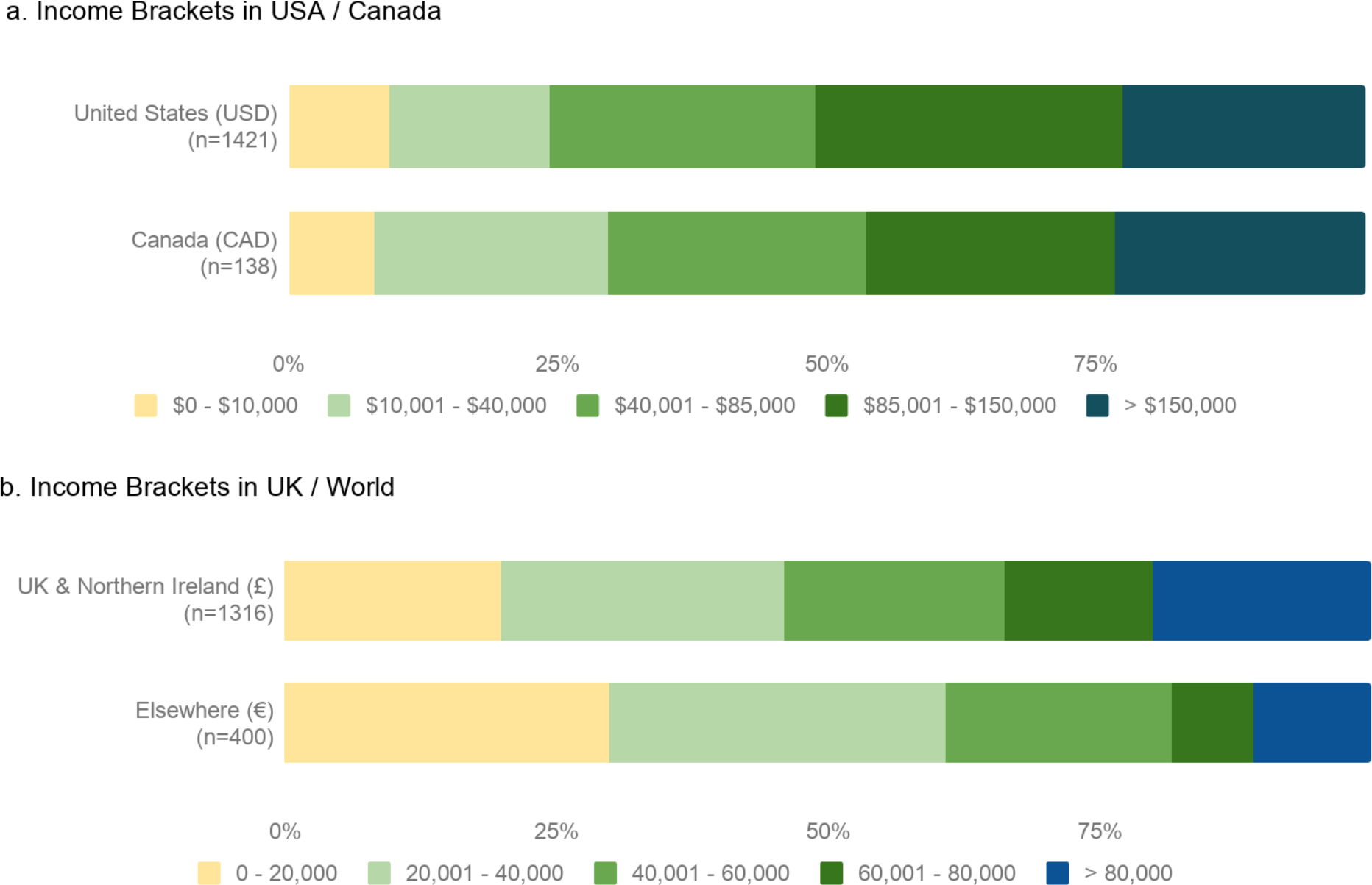
Income brackets by country. Stacked bar chart compares income brackets of participants from top countries and worldwide. Note that income brackets differ between a and b.

**Figure S2.**
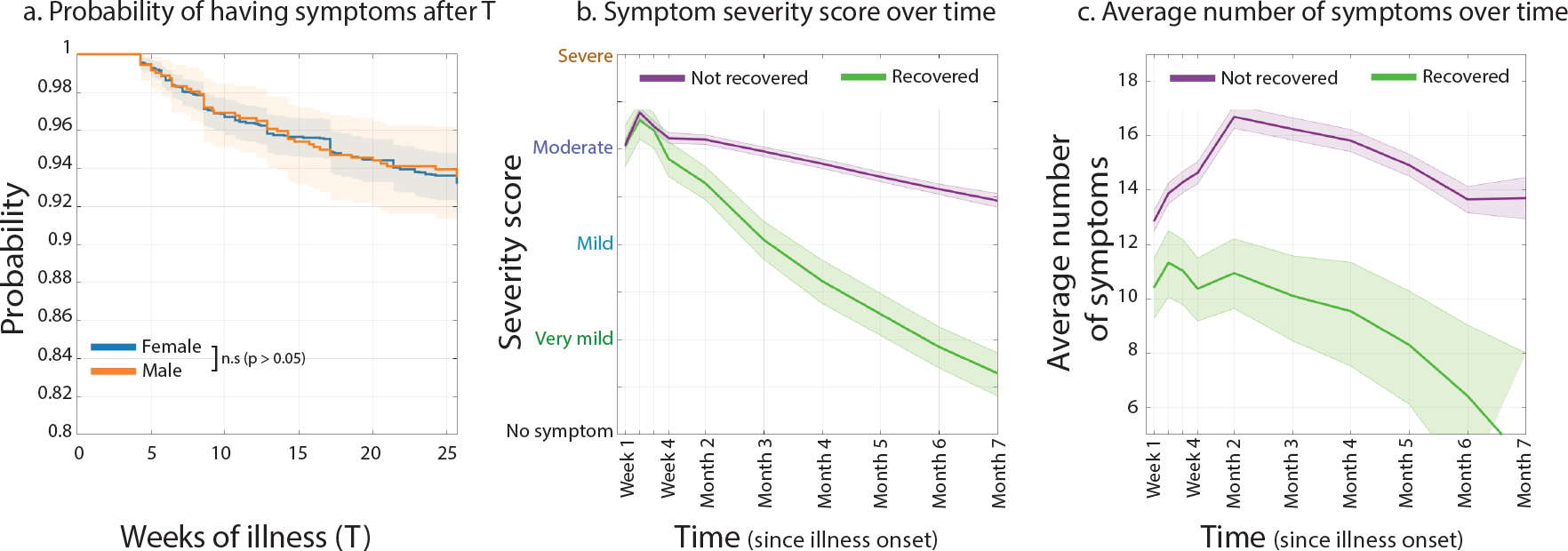
Figure 1 extension. **a.** Survival function for Male vs Female. (Kaplan-Meier estimator), characterizing the distribution of disease duration for Female (blue) and Male (orange) respondents. b. Average symptom severity over time, for “recovered” (green,) and “Not recovered” participants. c. Average number of reported symptoms over time, for “recovered” (green) and “Not recovered” participants

**Figure S3.**
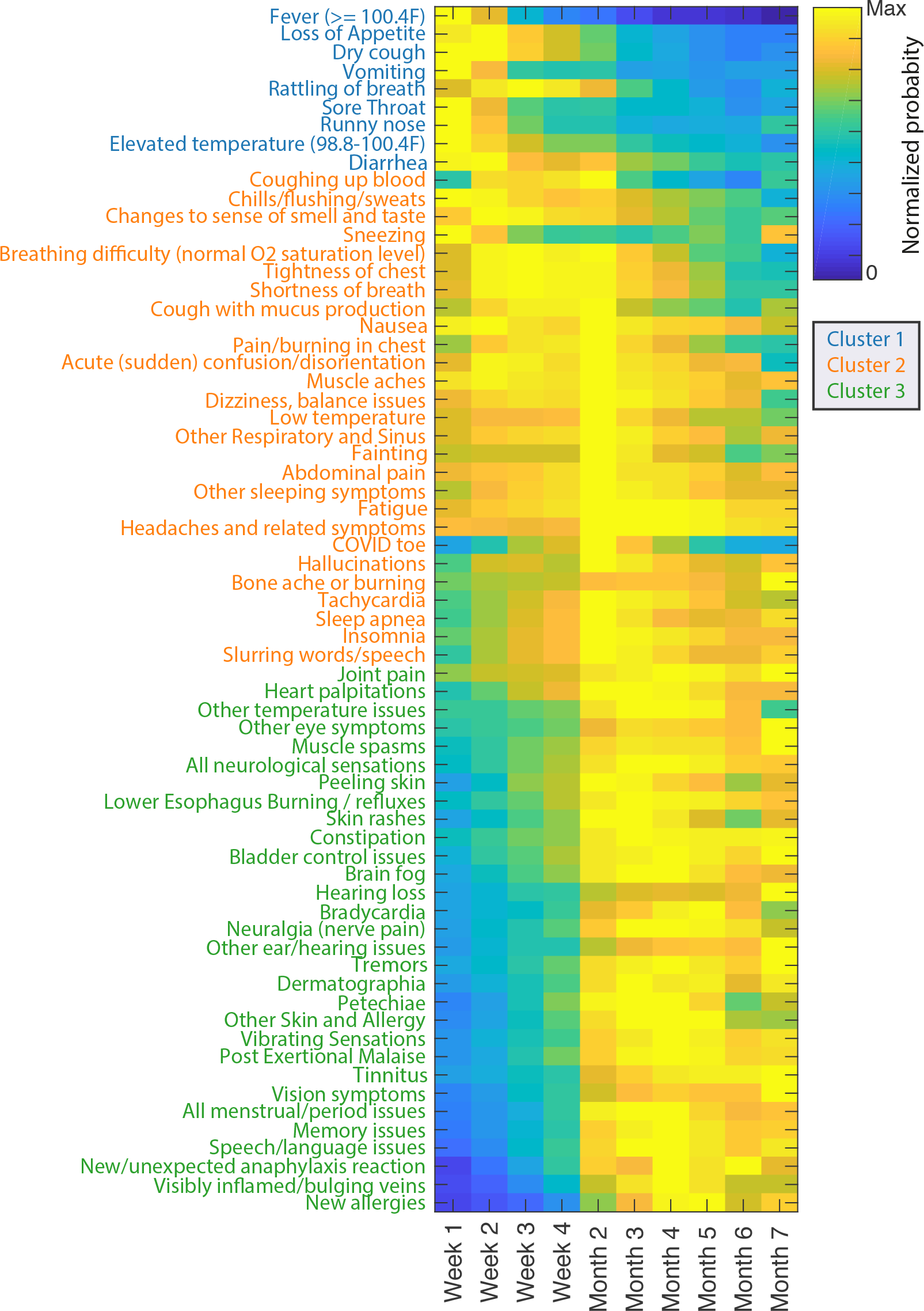
Normalized probability of symptoms over time. Heatmap shows the normalized probability of each symptom from week 1 to month 7. Rows are sorted using multidimensional scaling, to capture similarity in time course shapes such that similar shapes are adjacent.

**Figure S4.**
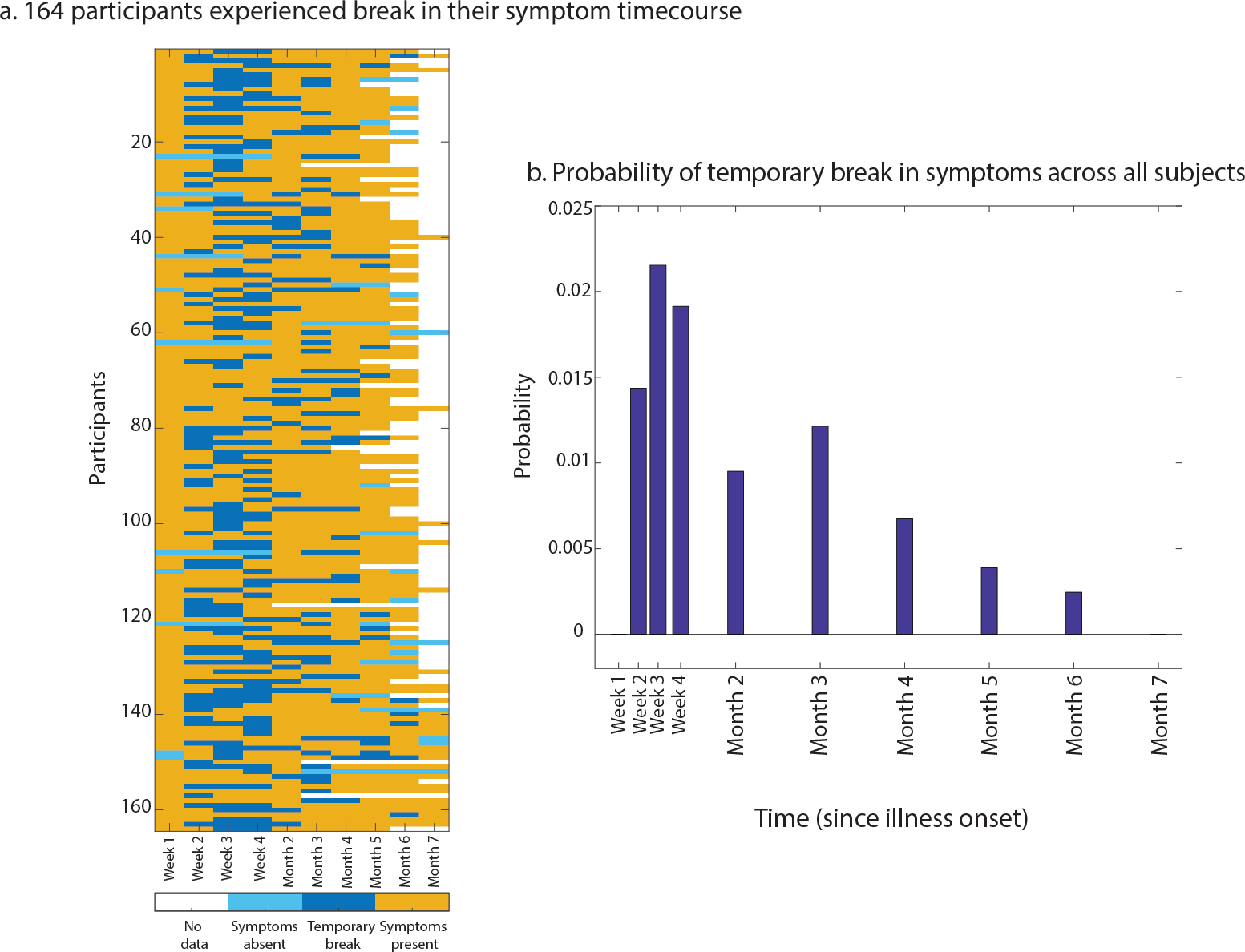
Symptom break in 4.4% of respondents. 164 out of 3762 subjects (4.4%) had a temporary break in symptoms, limited to the resolution that we’ve had (the first 4 weeks, and one data point for each month, until month seven). Yellow shows “symptom present”, dark blue indicates “temporary break” which is defined as a window between two “symptom present”. Light blue shows “symptom absent” i.e. a time point with no symptom, that has “symptom present” on only one side of it (either preceding or following). Light blue can happen either at the beginning, indicating symptoms that had not started yet, or symptoms not asked in the survey. They can also happen at the end, indicating recovery, or end of symptom reported. The right plot shows the probability of temporary breaks (dark blues), over all patients.

**Figure S5.**
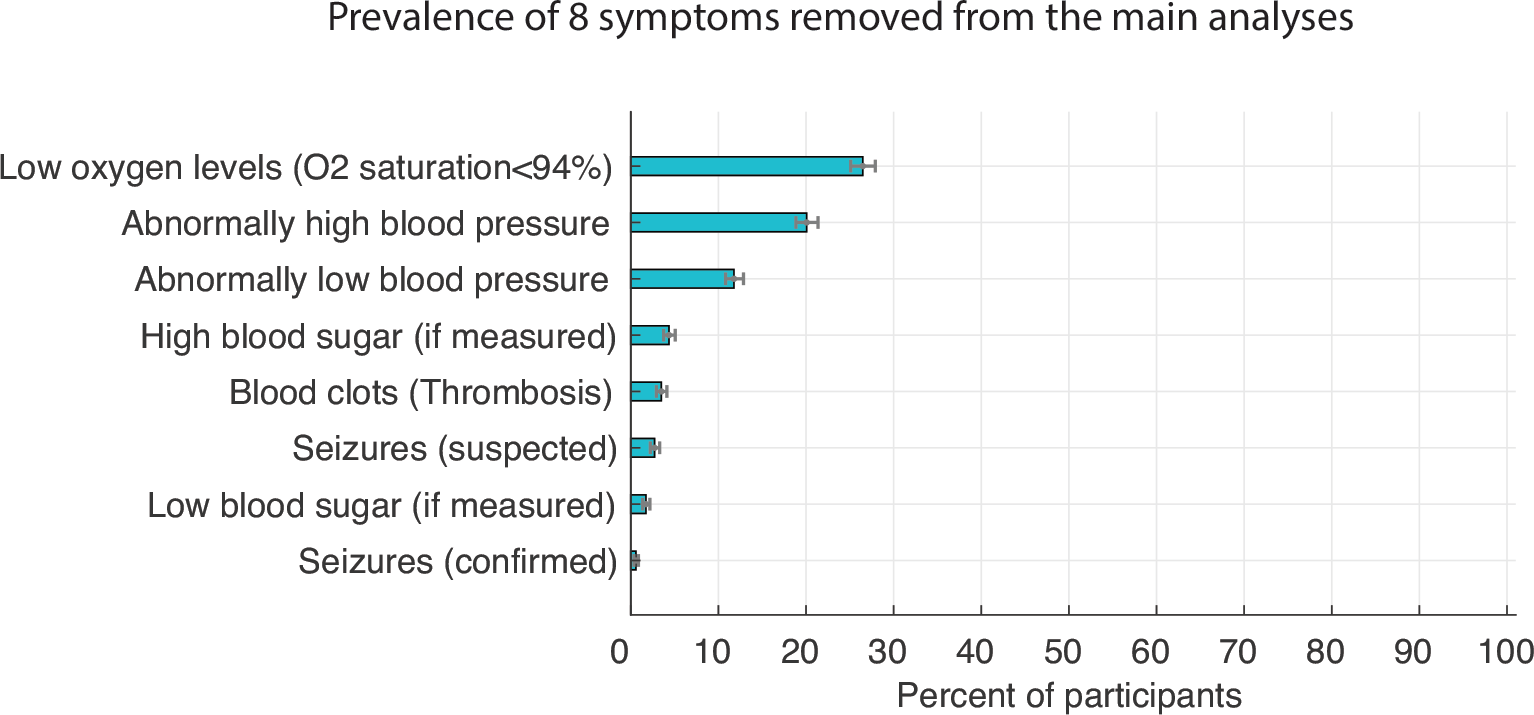
Prevalence of symptoms removed from the main analyses. Eight symptoms were excluded, as their measurement required specialized equipment or tests that many participants may not have had access to. Excluded symptoms included high blood pressure, low blood pressure, thrombosis, seizures (confirmed or suspected), low oxygen levels, high blood sugar, and low blood sugar.

**Figure S6.**
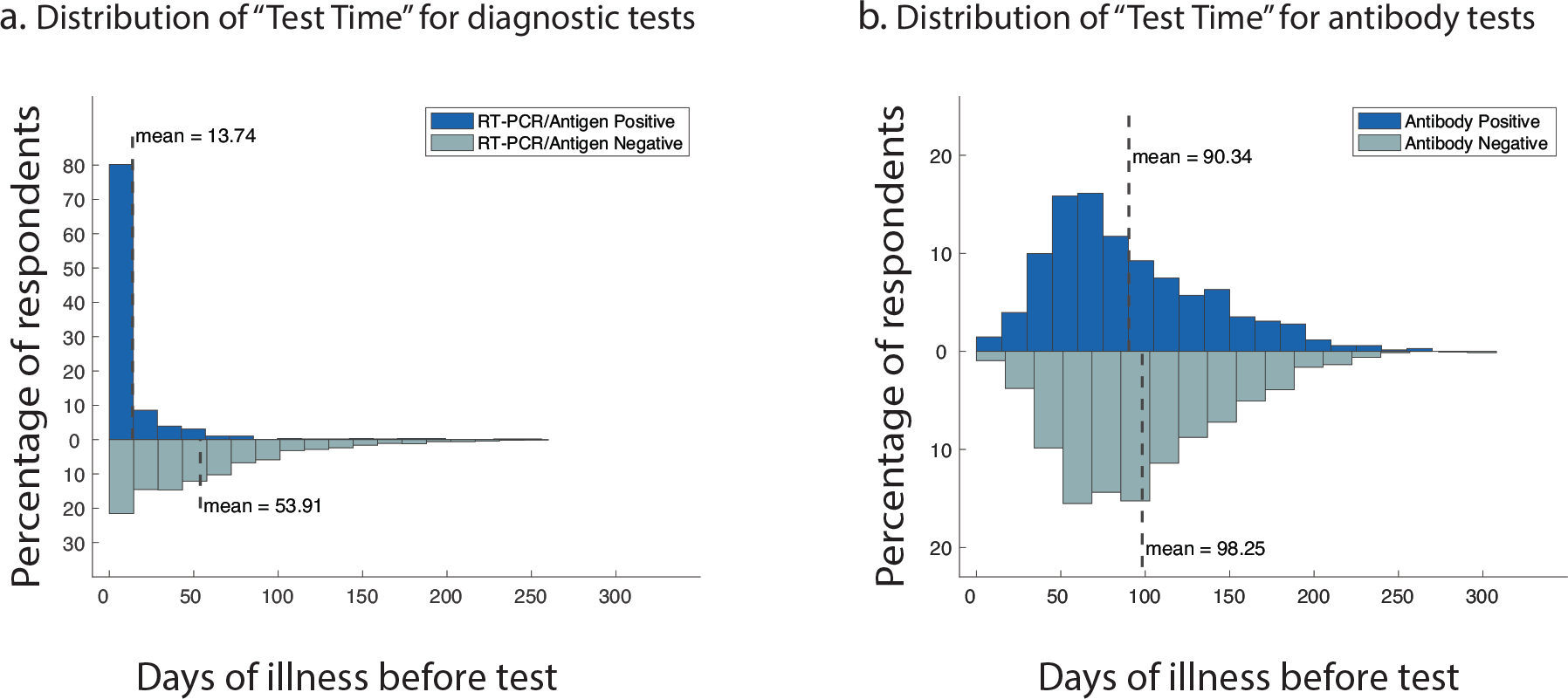
Test time. (a) Number of days between first experiencing symptoms and receiving a diagnostic test (RT-PCR or antigen) for Positively (blue) vs Negatively tested (grey) respondents. p < 0.001, Mann-Whitney U test (b) similar to a, for antibody testing.

**Figure S7.**
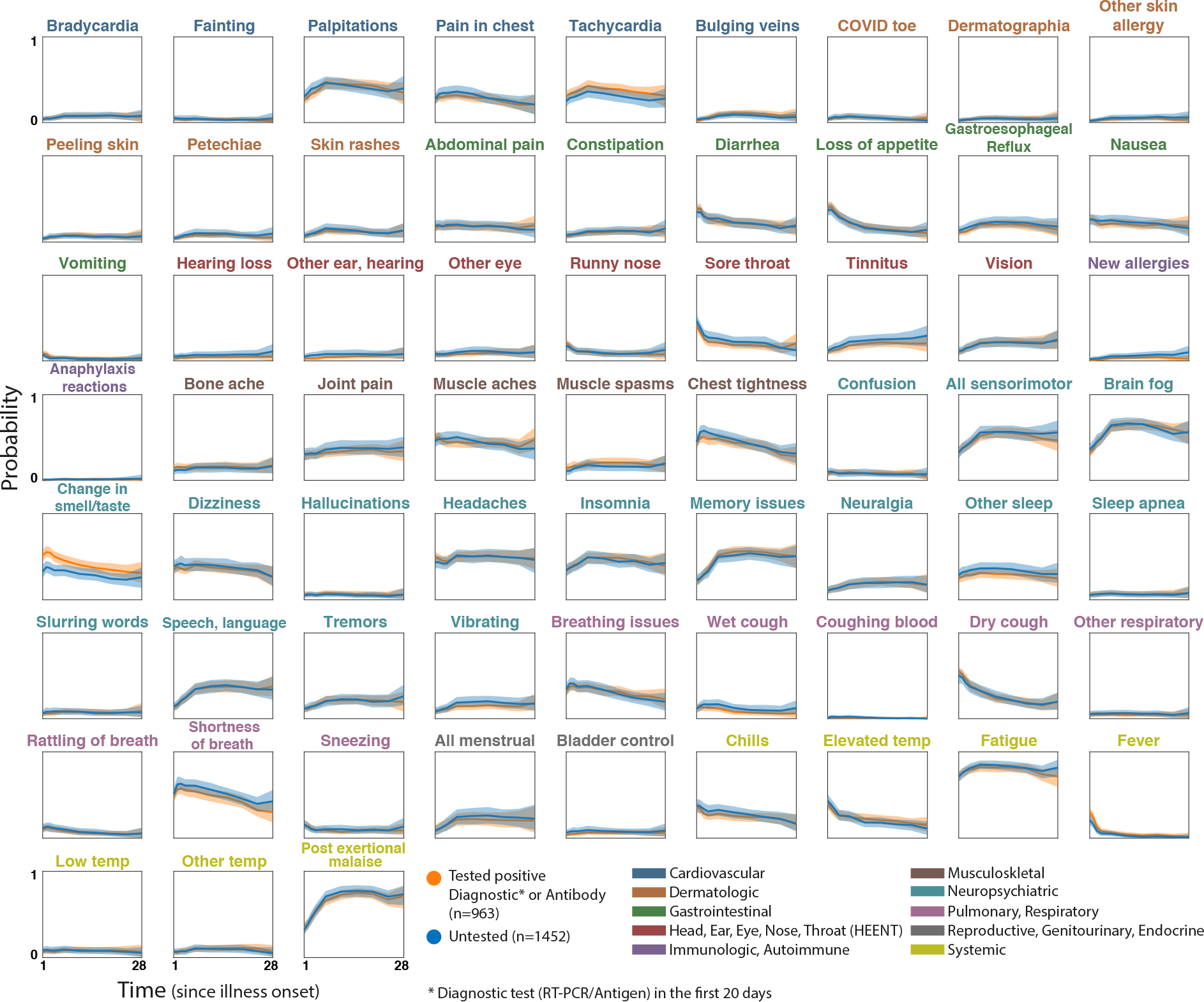
Symptom time courses for respondents with positive results vs. untested respondents. Plots show symptom time courses (similar to Fig. 7) for respondents who were confirmed COVID-positive via diagnostic or antibody testing (orange) vs those without any diagnostic or antibody testing (blue). Shaded regions show simultaneous 95% confidence bands (over time and symptoms). Symptom names are colored according to the affected organ systems.

**Figure S8.**
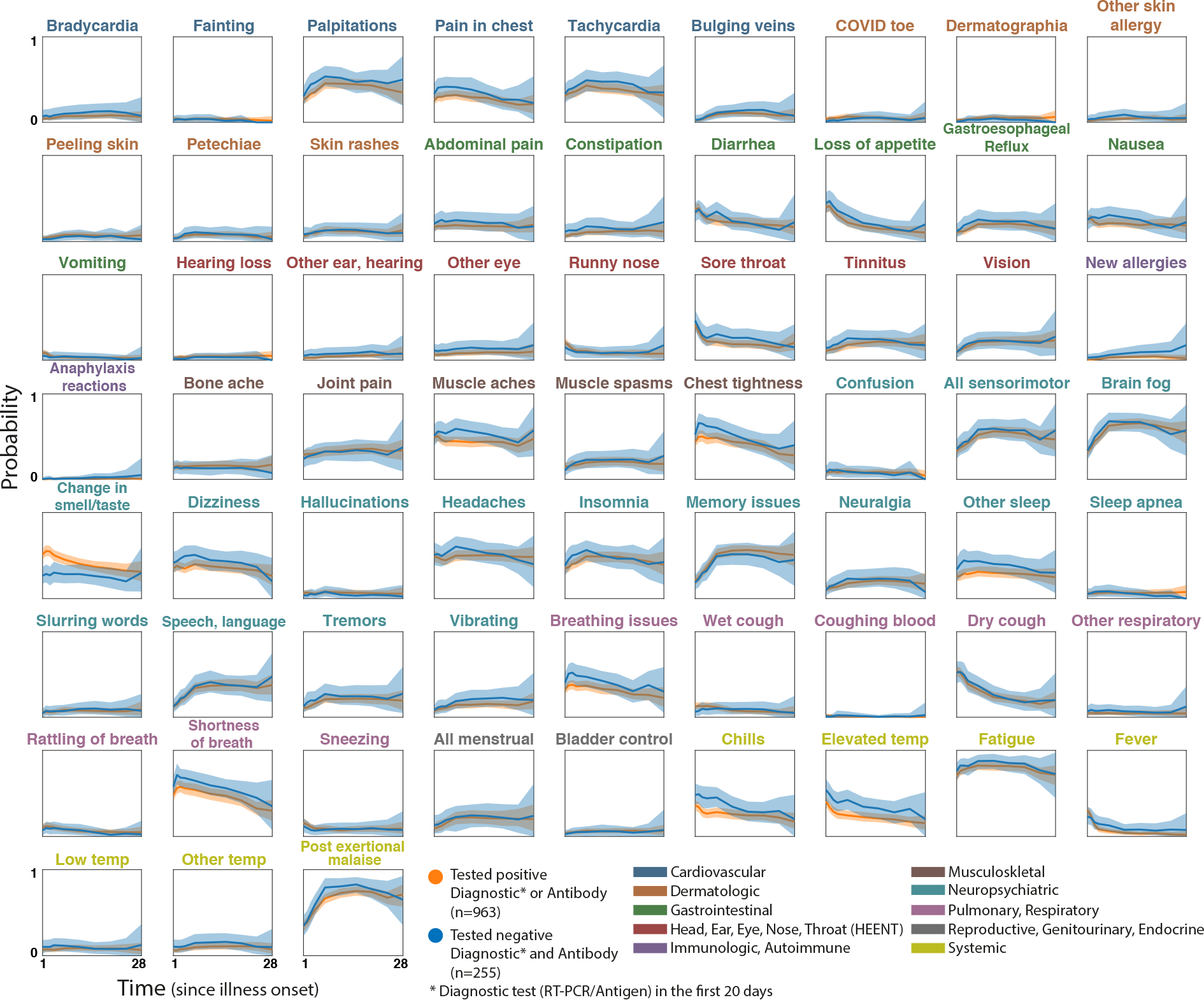
Symptom time courses for respondents with positive vs. negative test results. Plots show symptom time courses (similar to Fig. 7) for respondents who were confirmed COVID-positive via diagnostic or antibody testing (orange) vs those with negative diagnostic and antibody test results (blue). Shaded regions show simultaneous 95% confidence bands (over time and symptoms). Symptom names are colored according to the affected organ systems.

**Figure S9.**
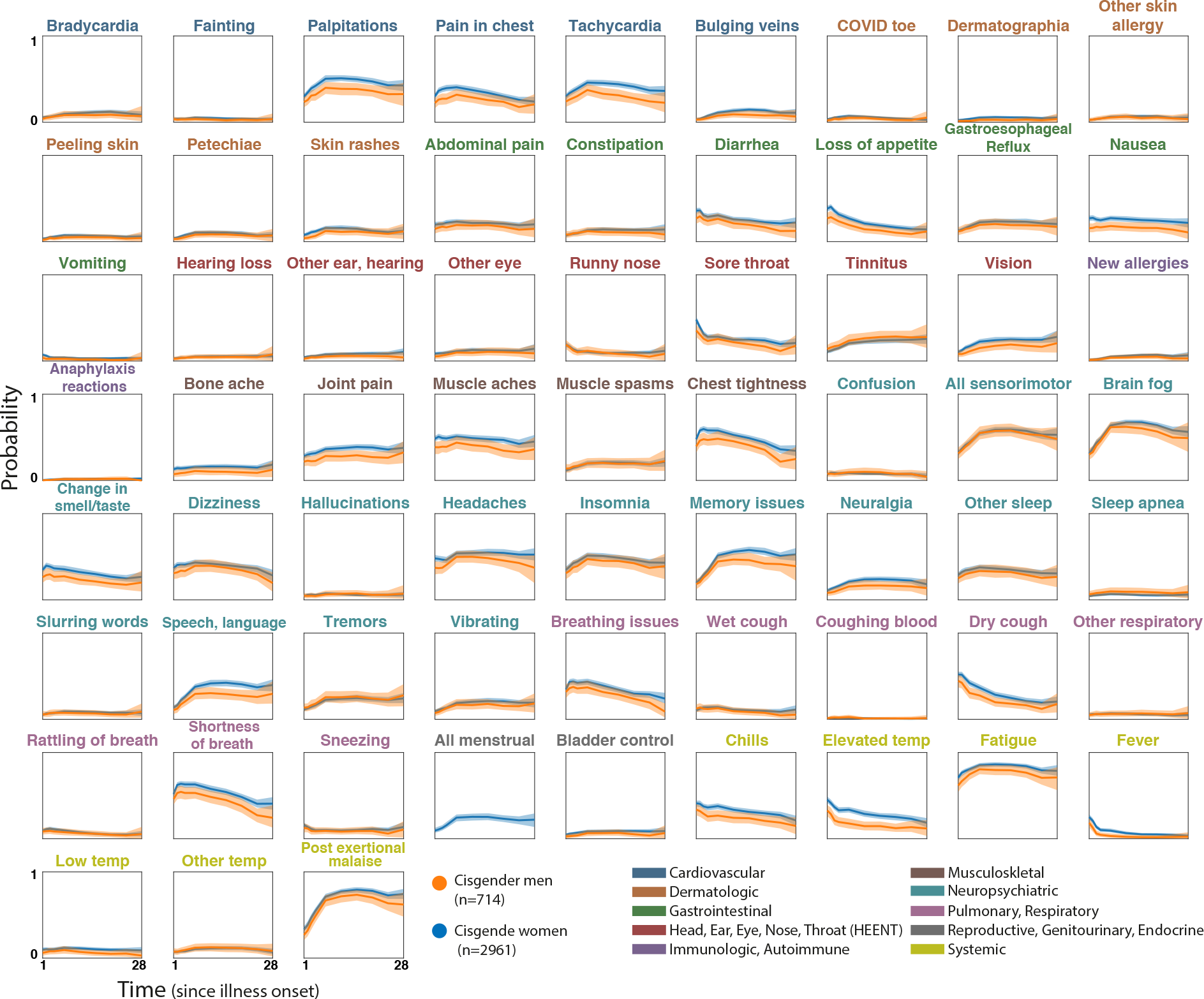
Symptom time courses for cisgender female vs. cisgender male respondents. Plots show symptom time courses (similar to Fig. 7) for cisgender women (blue) and cisgende men (orange). Shaded regions show simultaneous 95% confidence bands (over time and symptoms). Symptom names are colored according to the affected organ systems.

## Appendix D: Raw Data Tables

**Table S1.**
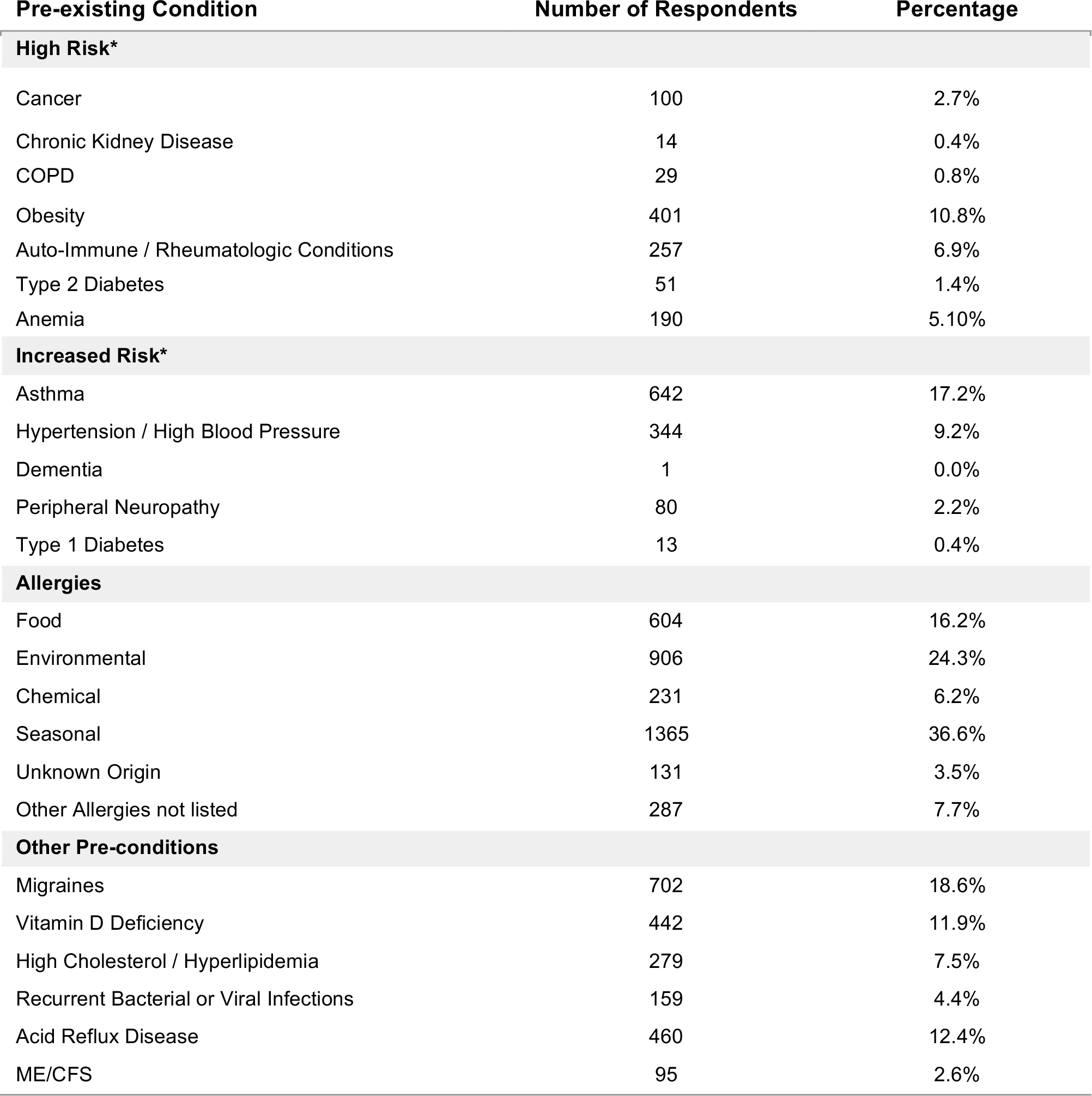
Pre-existing conditions reported by respondents.

**Table S2.**
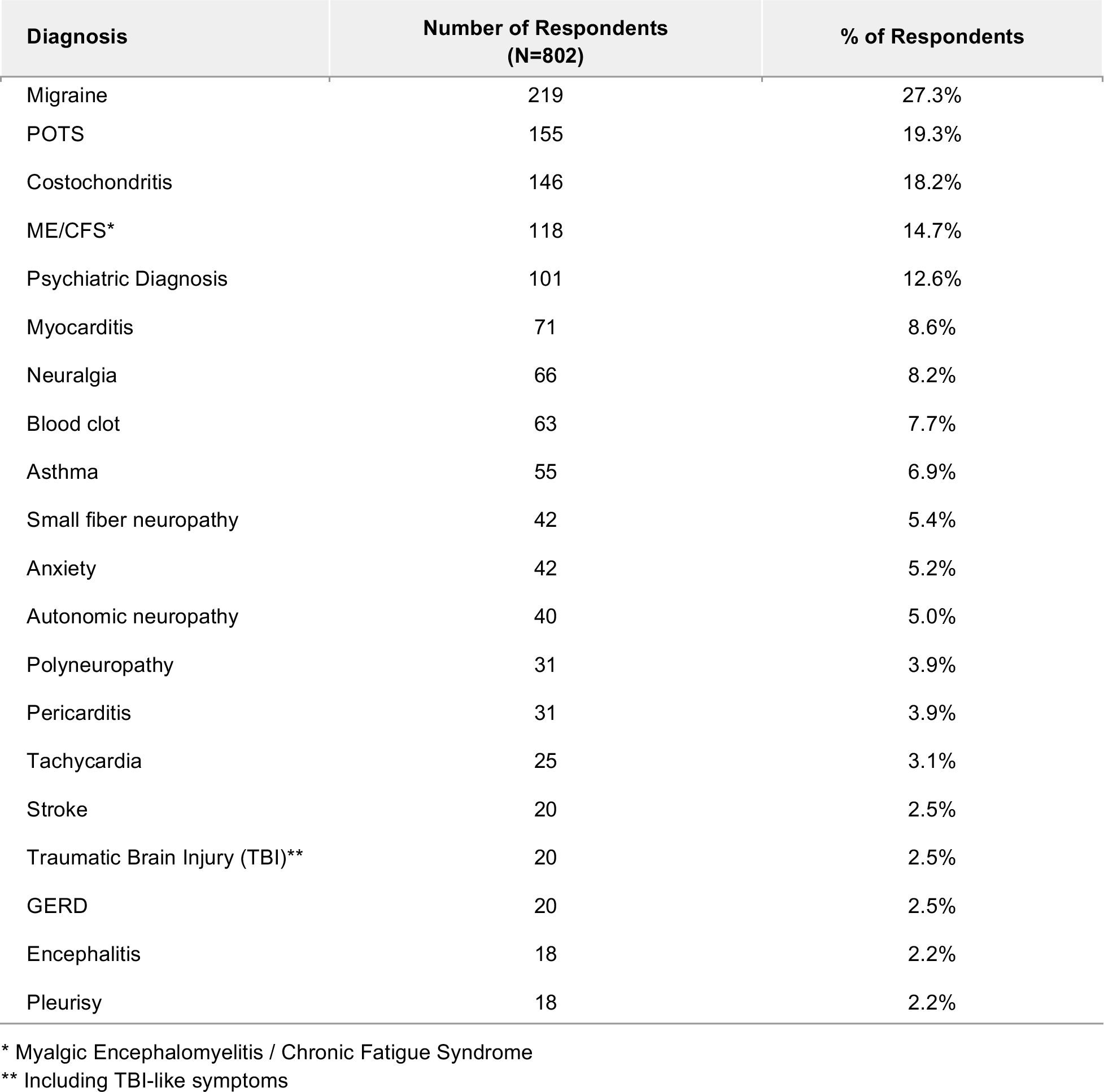
Diagnosis reported by respondents after the onset of illness.

| Diagnosis | Number of Respondents<br>(N=802) | % of Respondents |
| --- | --- | --- |
| Migraine | 219 | 27.3% |
| POTS | 155 | 19.3% |
| Costochondritis | 146 | 18.2% |
| ME/CFS* | 118 | 14.7% |
| Psychiatric Diagnosis | 101 | 12.6% |
| Myocarditis | 71 | 8.6% |
| Neuralgia | 66 | 8.2% |
| Blood clot | 63 | 7.7% |
| Asthma | 55 | 6.9% |
| Small fiber neuropathy | 42 | 5.4% |
| Anxiety | 42 | 5.2% |
| Autonomic neuropathy | 40 | 5.0% |
| Polyneuropathy | 31 | 3.9% |
| Pericarditis | 31 | 3.9% |
| Tachycardia | 25 | 3.1% |
| Stroke | 20 | 2.5% |
| Traumatic Brain Injury (TBI)** | 20 | 2.5% |
| GERD | 20 | 2.5% |
| Encephalitis | 18 | 2.2% |
| Pleurisy | 18 | 2.2% |

\* Myalgic Encephalomyelitis / Chronic Fatigue Syndrome
\*\* Including TBI-like symptoms

**Table S3.** Top 4 affected body parts of sensorimotor symptoms.

| Symptom | Top location | 2nd location | 3rd location | 4th location |
| --- | --- | --- | --- | --- |
| Numbness/Loss of Sensation | hand (n>413) | foot/feet (n>336) | arm (n>336) | leg (n>201) |
| Coldness | foot/feet (n>356) | hand (n>237) | body (n>157) | arm (n>62) |
| Tingling/Prickling/Pins & Needles | hand (n>482) | foot/feet (n>432) | arm (n>355) | leg (n>252) |
| Electric Zap/Shock Sensation | leg (n>131) | arm (n>120) | foot/feet (n>93) | head (n>85) |
| Facial Paralysis | side (n>51) | cheek (n>26) | mouth (n>14) | eye/jaw* (n>13) |
| Facial Pressure/Numbness | sinus (n>32) | face (n>31) | side (n>29) | lips/head** (n>24) |
| Weakness | leg (n>377) | arm (n>319) | body (n>218) | hand (n>101) |

\* Eye and jaw were reported equally for facial paralysis
\*\* Lips and head were reported equally for facial pressure and numbness

**Table S4.**
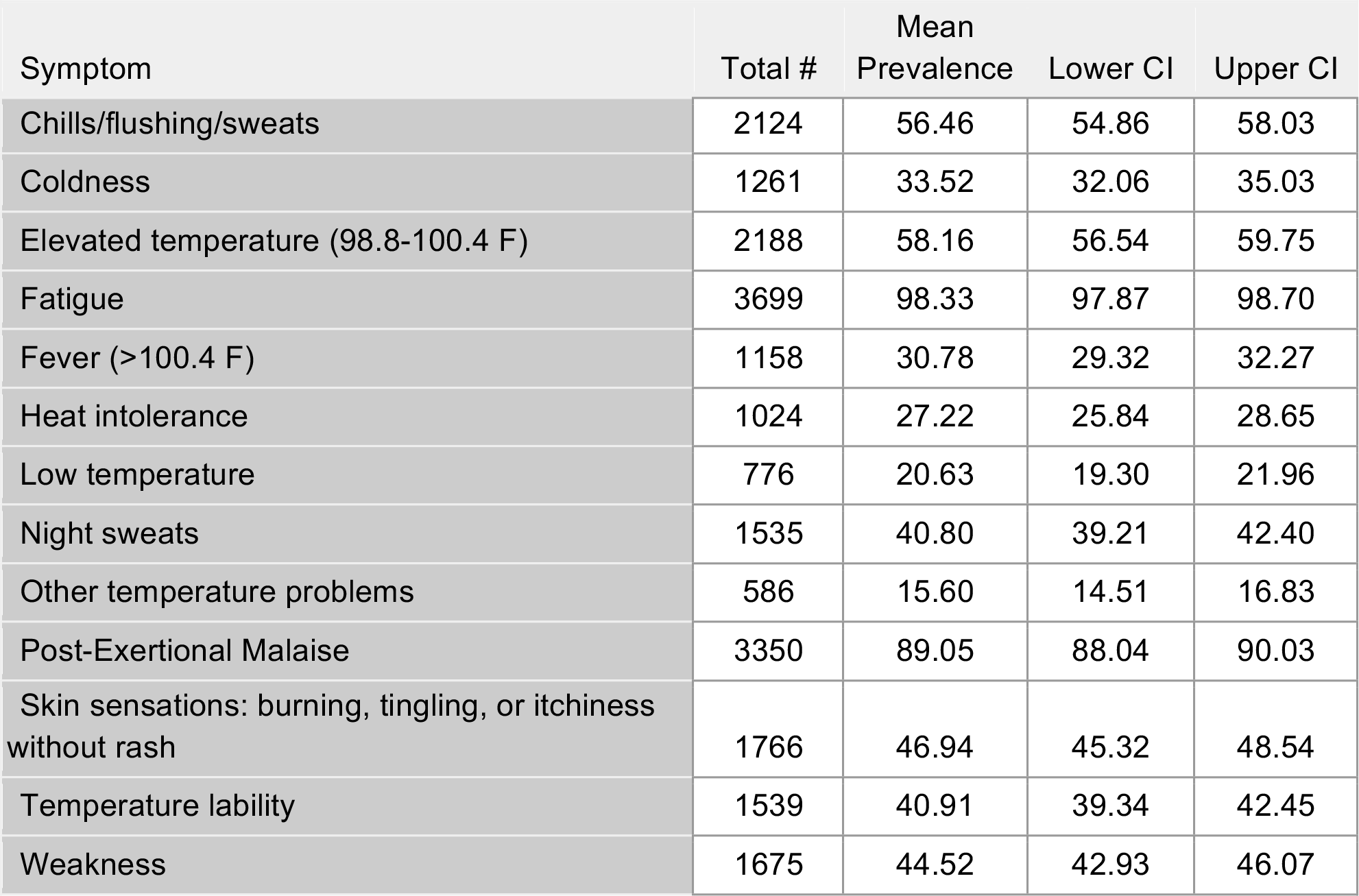
Systemic Symptom Prevalence Data.

**Table S5.**
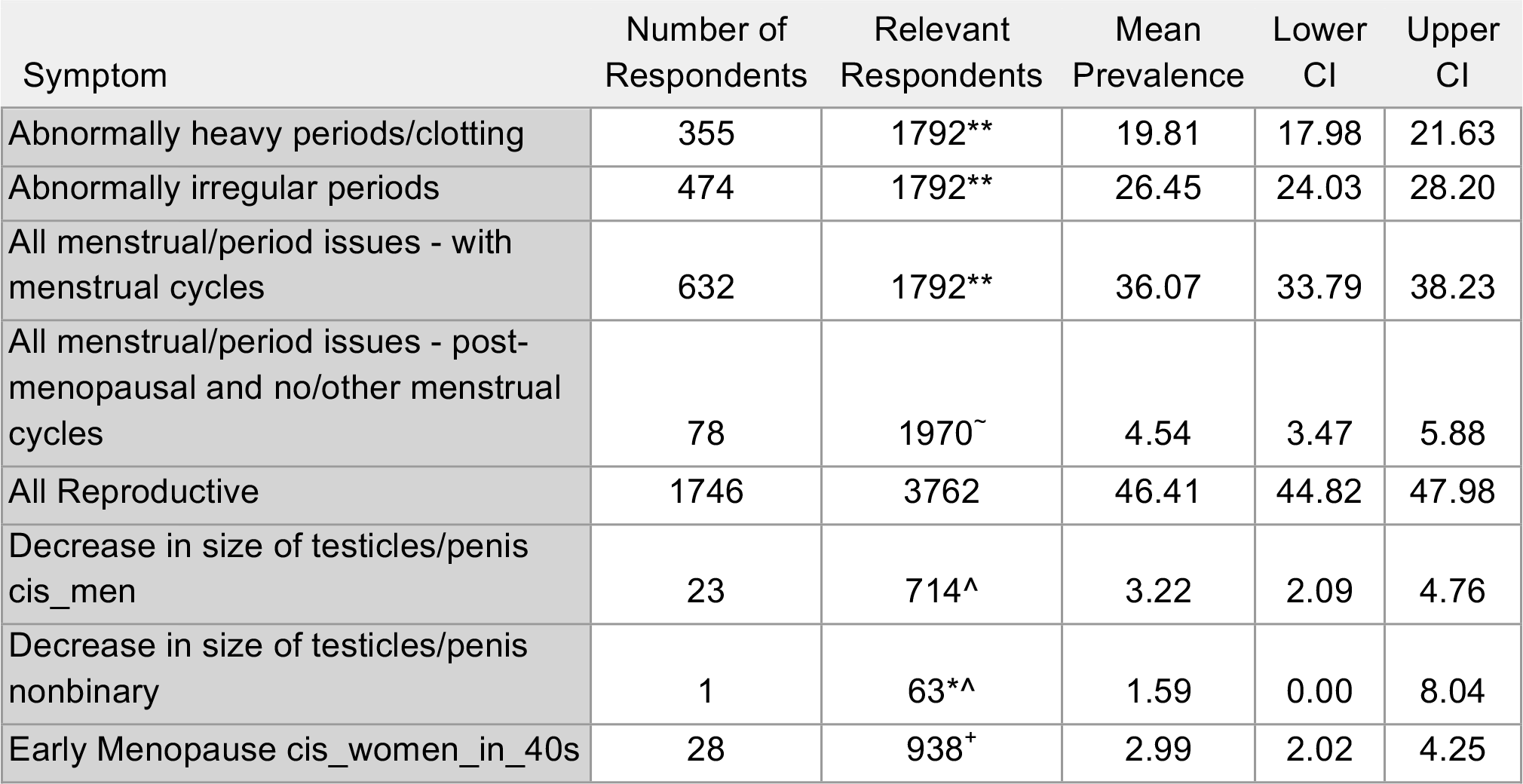
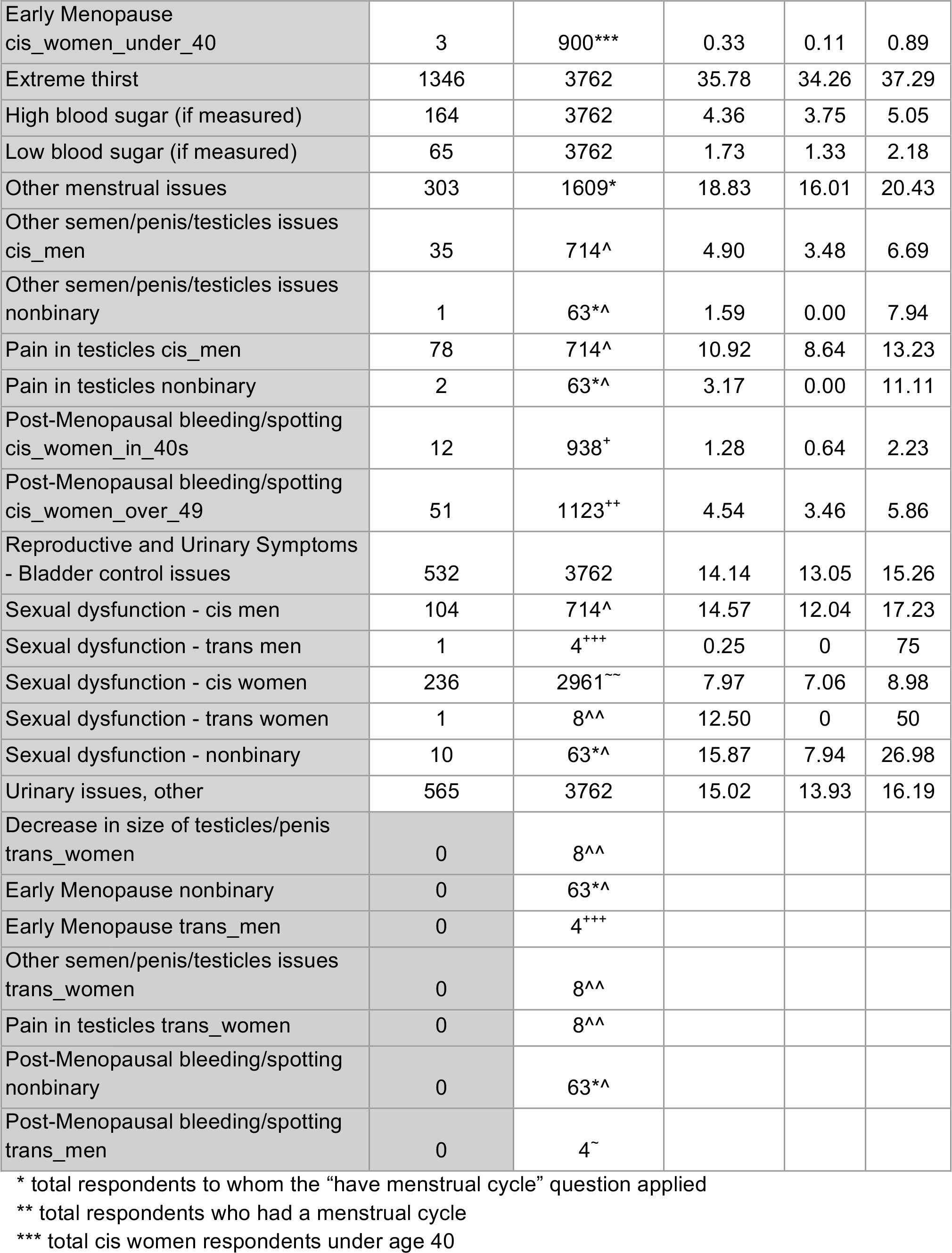
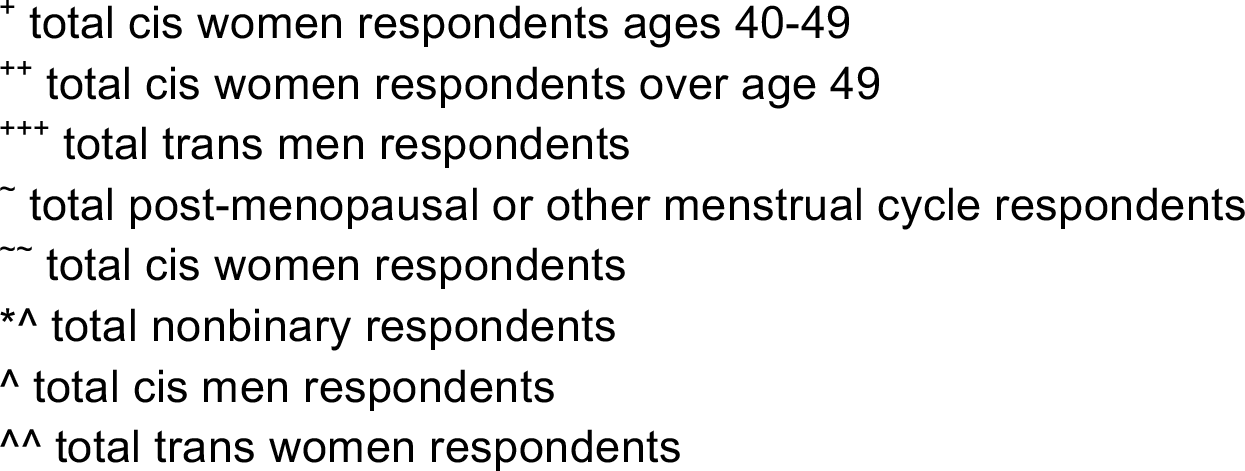
Reproductive/Genitourinary/Endocrine Symptom Prevalence Data.

| Symptom | Number of Respondents | Relevant Respondents | Mean Prevalence | Lower CI | Upper CI |
| --- | --- | --- | --- | --- | --- |
| Abnormally heavy periods/clotting | 355 | 1792** | 19.81 | 17.98 | 21.63 |
| Abnormally irregular periods | 474 | 1792** | 26.45 | 24.03 | 28.20 |
| All menstrual/period issues - with menstrual cycles | 632 | 1792** | 36.07 | 33.79 | 38.23 |
| All menstrual/period issues - post-menopausal and no/other menstrual cycles | 78 | 1970~ | 4.54 | 3.47 | 5.88 |
| All Reproductive | 1746 | 3762 | 46.41 | 44.82 | 47.98 |
| Decrease in size of testicles/penis cis_men | 23 | 714^ | 3.22 | 2.09 | 4.76 |
| Decrease in size of testicles/penis nonbinary | 1 | 63*^ | 1.59 | 0.00 | 8.04 |
| Early Menopause cis_women_in_40s | 28 | 938+ | 2.99 | 2.02 | 4.25 |
| Early Menopause<br>cis_women_under_40 | 3 | 900*** | 0.33 | 0.11 | 0.89 |
| Extreme thirst | 1346 | 3762 | 35.78 | 34.26 | 37.29 |
| High blood sugar (if measured) | 164 | 3762 | 4.36 | 3.75 | 5.05 |
| Low blood sugar (if measured) | 65 | 3762 | 1.73 | 1.33 | 2.18 |
| Other menstrual issues | 303 | 1609* | 18.83 | 16.01 | 20.43 |
| Other semen/penis/testicles issues<br>cis_men | 35 | 714^ | 4.90 | 3.48 | 6.69 |
| Other semen/penis/testicles issues<br>nonbinary | 1 | 63*^ | 1.59 | 0.00 | 7.94 |
| Pain in testicles cis_men | 78 | 714^ | 10.92 | 8.64 | 13.23 |
| Pain in testicles nonbinary | 2 | 63*^ | 3.17 | 0.00 | 11.11 |
| Post-Menopausal bleeding/spotting<br>cis_women_in_40s | 12 | 938<sup>+</sup> | 1.28 | 0.64 | 2.23 |
| Post-Menopausal bleeding/spotting<br>cis_women_over_49 | 51 | 1123<sup>++</sup> | 4.54 | 3.46 | 5.86 |
| Reproductive and Urinary Symptoms<br>- Bladder control issues | 532 | 3762 | 14.14 | 13.05 | 15.26 |
| Sexual dysfunction - cis men | 104 | 714^ | 14.57 | 12.04 | 17.23 |
| Sexual dysfunction - trans men | 1 | 4<sup>+++</sup> | 0.25 | 0 | 75 |
| Sexual dysfunction - cis women | 236 | 2961<sup>~~</sup> | 7.97 | 7.06 | 8.98 |
| Sexual dysfunction - trans women | 1 | 8^^ | 12.50 | 0 | 50 |
| Sexual dysfunction - nonbinary | 10 | 63*^ | 15.87 | 7.94 | 26.98 |
| Urinary issues, other | 565 | 3762 | 15.02 | 13.93 | 16.19 |
| Decrease in size of testicles/penis<br>trans_women | 0 | 8^^ |  |  |  |
| Early Menopause nonbinary | 0 | 63*^ |  |  |  |
| Early Menopause trans_men | 0 | 4<sup>+++</sup> |  |  |  |
| Other semen/penis/testicles issues<br>trans_women | 0 | 8^^ |  |  |  |
| Pain in testicles trans_women | 0 | 8^^ |  |  |  |
| Post-Menopausal bleeding/spotting<br>nonbinary | 0 | 63*^ |  |  |  |
| Post-Menopausal bleeding/spotting<br>trans_men | 0 | 4<sup>~</sup> |  |  |  |

\* total respondents to whom the “have menstrual cycle” question applied
\*\* total respondents who had a menstrual cycle
\*\*\* total cis women respondents under age 40
<sup>+</sup> total cis women respondents ages 40-49 <sup>++</sup> total cis women respondents over age 49 <sup>+++</sup> total trans men respondents <sup>~</sup> total post-menopausal or other menstrual cycle respondents <sup>~~</sup> total cis women respondents <sup>\*^</sup> total nonbinary respondents <sup>^</sup> total cis men respondents <sup>^^</sup> total trans women respondents

**Table S6.** Cardiovascular Symptom Prevalence Data.

| Symptom | Number of Respondents | Mean Prevalence | Lower CI | Upper CI |
| --- | --- | --- | --- | --- |
| Abnormally high blood pressure | 755 | 20.07 | 18.79 | 21.35 |
| Abnormally low blood pressure | 443 | 11.78 | 10.77 | 12.84 |
| Blood clots (Thrombosis) | 131 | 3.48 | 2.95 | 4.12 |
| Bradycardia (low heart rate, <60 beats per minute) | 658 | 17.49 | 16.27 | 18.69 |
| Fainting | 486 | 12.92 | 11.88 | 14.01 |
| Heart palpitations | 2534 | 67.36 | 65.87 | 68.82 |
| Pain/burning in chest | 1997 | 53.08 | 51.54 | 54.68 |
| Tachycardia | 2308 | 61.35 | 59.78 | 62.89 |

**Table S7.** Musculoskeletal Symptom Prevalence Data.

| Symptom | Number of Respondents | Mean Prevalence | Lower CI | Upper CI |
| --- | --- | --- | --- | --- |
| Joint pain | 1962 | 52.15 | 50.53 | 53.77 |
| Muscle aches | 2601 | 69.14 | 67.62 | 70.60 |
| Bone ache or burning | 910 | 24.19 | 22.83 | 25.57 |
| Muscle spasms | 1222 | 32.48 | 30.97 | 33.97 |
| Stiff neck | 1471 | 39.10 | 37.59 | 40.70 |
| Tightness of Chest | 2813 | 74.77 | 73.37 | 76.13 |

**Table S8.**
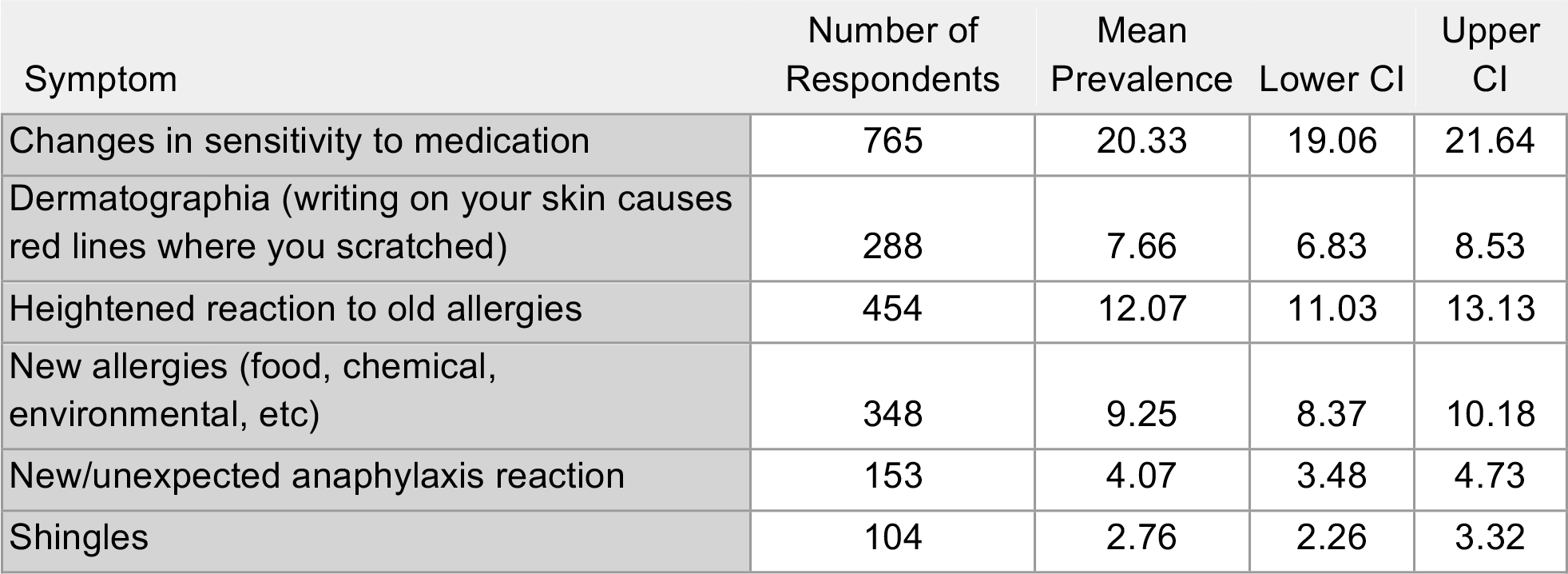
Immunologic/Autoimmune Symptom Prevalence Data.

| Symptom | Number of Respondents | Mean Prevalence | Lower CI | Upper CI |
| --- | --- | --- | --- | --- |
| Changes in sensitivity to medication | 765 | 20.33 | 19.06 | 21.64 |
| Dermatographia (writing on your skin causes red lines where you scratched) | 288 | 7.66 | 6.83 | 8.53 |
| Heightened reaction to old allergies | 454 | 12.07 | 11.03 | 13.13 |
| New allergies (food, chemical, environmental, etc) | 348 | 9.25 | 8.37 | 10.18 |
| New/unexpected anaphylaxis reaction | 153 | 4.07 | 3.48 | 4.73 |
| Shingles | 104 | 2.76 | 2.26 | 3.32 |

**Table S9.**
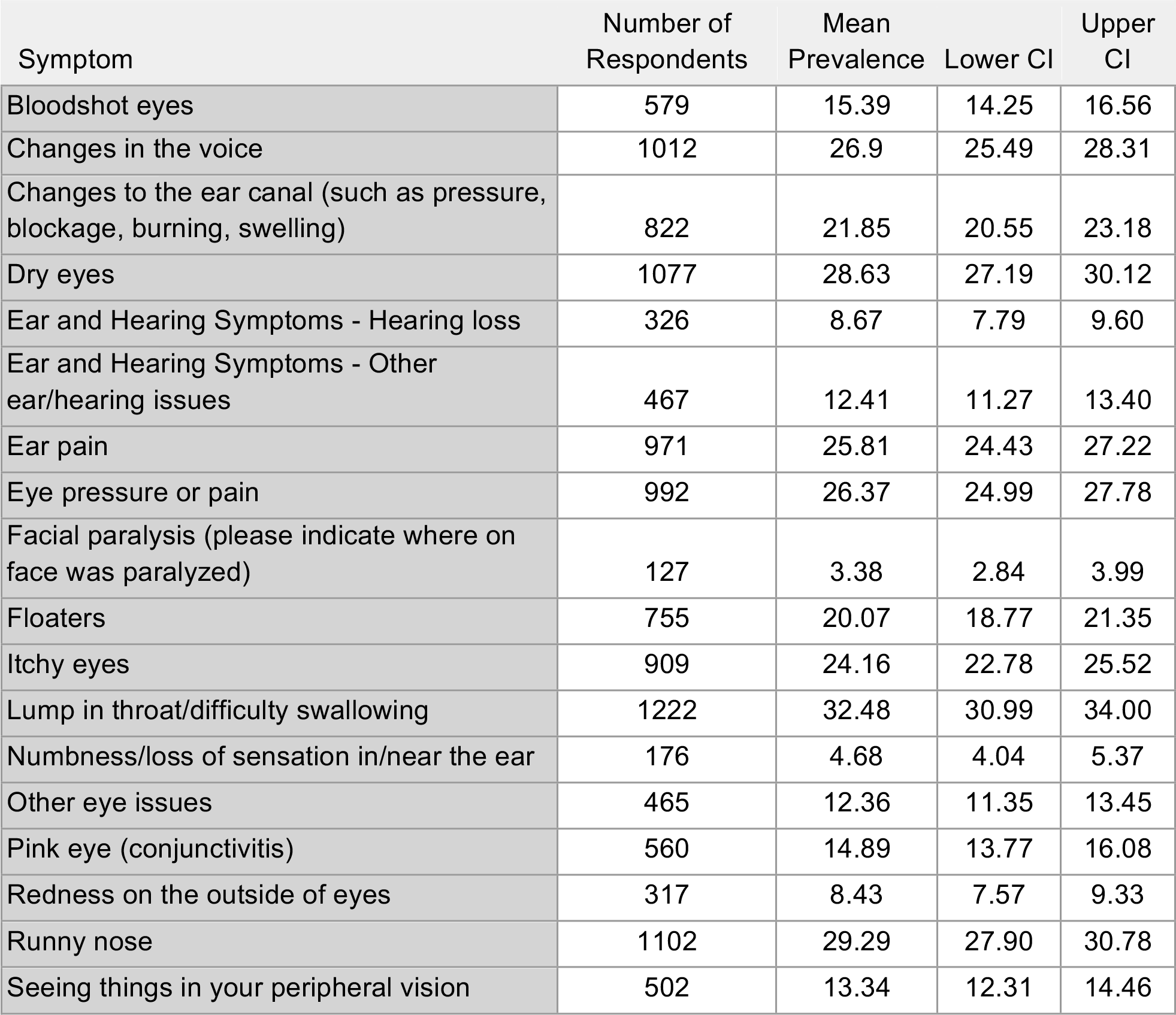
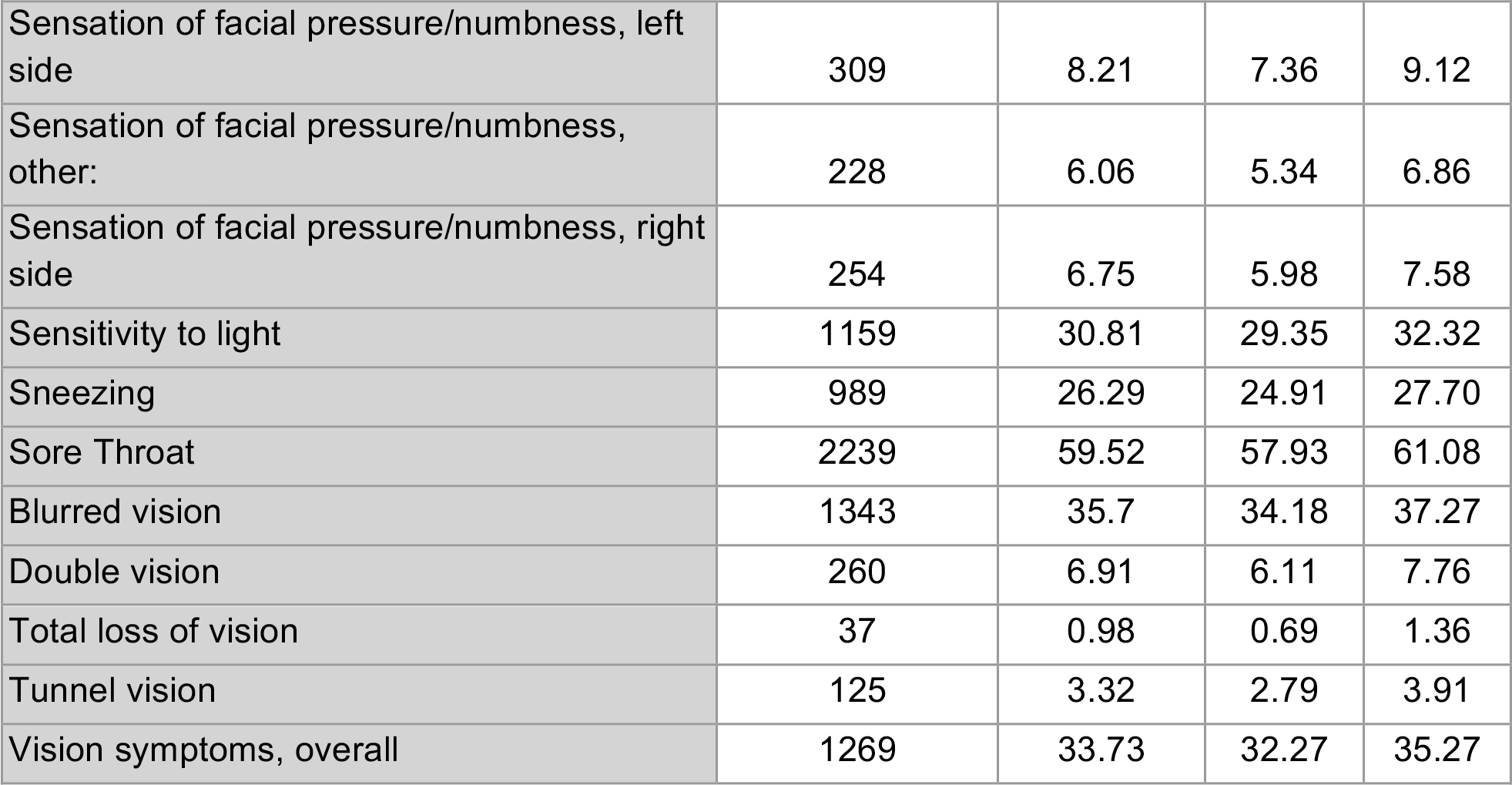
HEENT (Head, Ear, Eyes, Nose, Throat) Symptom Prevalence Data.

| Symptom | Number of Respondents | Mean Prevalence | Lower CI | Upper CI |
| --- | --- | --- | --- | --- |
| Bloodshot eyes | 579 | 15.39 | 14.25 | 16.56 |
| Changes in the voice | 1012 | 26.9 | 25.49 | 28.31 |
| Changes to the ear canal (such as pressure, blockage, burning, swelling) | 822 | 21.85 | 20.55 | 23.18 |
| Dry eyes | 1077 | 28.63 | 27.19 | 30.12 |
| Ear and Hearing Symptoms - Hearing loss | 326 | 8.67 | 7.79 | 9.60 |
| Ear and Hearing Symptoms - Other ear/hearing issues | 467 | 12.41 | 11.27 | 13.40 |
| Ear pain | 971 | 25.81 | 24.43 | 27.22 |
| Eye pressure or pain | 992 | 26.37 | 24.99 | 27.78 |
| Facial paralysis (please indicate where on face was paralyzed) | 127 | 3.38 | 2.84 | 3.99 |
| Floaters | 755 | 20.07 | 18.77 | 21.35 |
| Itchy eyes | 909 | 24.16 | 22.78 | 25.52 |
| Lump in throat/difficulty swallowing | 1222 | 32.48 | 30.99 | 34.00 |
| Numbness/loss of sensation in/near the ear | 176 | 4.68 | 4.04 | 5.37 |
| Other eye issues | 465 | 12.36 | 11.35 | 13.45 |
| Pink eye (conjunctivitis) | 560 | 14.89 | 13.77 | 16.08 |
| Redness on the outside of eyes | 317 | 8.43 | 7.57 | 9.33 |
| Runny nose | 1102 | 29.29 | 27.90 | 30.78 |
| Seeing things in your peripheral vision | 502 | 13.34 | 12.31 | 14.46 |
| Sensation of facial pressure/numbness, left side | 309 | 8.21 | 7.36 | 9.12 |
| Sensation of facial pressure/numbness, other: | 228 | 6.06 | 5.34 | 6.86 |
| Sensation of facial pressure/numbness, right side | 254 | 6.75 | 5.98 | 7.58 |
| Sensitivity to light | 1159 | 30.81 | 29.35 | 32.32 |
| Sneezing | 989 | 26.29 | 24.91 | 27.70 |
| Sore Throat | 2239 | 59.52 | 57.93 | 61.08 |
| Blurred vision | 1343 | 35.7 | 34.18 | 37.27 |
| Double vision | 260 | 6.91 | 6.11 | 7.76 |
| Total loss of vision | 37 | 0.98 | 0.69 | 1.36 |
| Tunnel vision | 125 | 3.32 | 2.79 | 3.91 |
| Vision symptoms, overall | 1269 | 33.73 | 32.27 | 35.27 |

**Table S10.** Pulmonary Symptom Prevalence Data.

| Symptom | Number of Respondents | Mean Prevalence | Lower CI | Upper CI |
| --- | --- | --- | --- | --- |
| Cough with mucus production | 1062 | 28.23 | 26.82 | 29.72 |
| Dry cough | 2491 | 66.21 | 64.67 | 67.70 |
| Episodes of breathing difficulty/gasping for air when your oxygen saturation is normal | 2271 | 60.37 | 58.80 | 61.94 |
| Low oxygen levels (<94%)* | 996 | 26.48 | 25.07 | 27.91 |
| Coughing up Blood | 194 | 5.16 | 4.49 | 5.90 |
| Other Respiratory and Sinus Symptoms | 359 | 9.54 | 8.64 | 10.50 |
| Rattling of breath | 641 | 17.04 | 15.84 | 18.26 |
| Shortness of Breath | 2913 | 77.43 | 76.08 | 78.76 |

\* Measurement required specialized equipment or tests that many participants may not have had access to.

**Table S11.** Gastrointestinal Symptom Prevalence Data.

| Symptom | Number of Respondents | Mean Prevalence | Lower CI | Upper CI |
| --- | --- | --- | --- | --- |
| Abdominal pain | 1492 | 39.66 | 38.12 | 41.23 |
| Constipation | 930 | 24.72 | 23.34 | 26.10 |
| Diarrhea | 2246 | 59.7 | 58.13 | 61.27 |
| Feeling full quickly when eating | 1158 | 30.78 | 29.32 | 32.22 |
| Hyperactive bowel sensations | 881 | 24.00 | 22.62 | 25.33 |
| Loss of Appetite | 1942 | 51.62 | 49.95 | 53.22 |
| Lower Esophagus Burning / gastroesophageal reflux / acid reflux | 1317 | 35.01 | 33.47 | 36.52 |
| Nausea | 1797 | 47.77 | 46.17 | 49.36 |
| Vomiting | 539 | 14.33 | 13.24 | 15.47 |

**Table S12.** Dermatologic Symptom Prevalence Data.

| Symptom | Number of Respondents | Mean Prevalence | Lower CI | Upper CI |
| --- | --- | --- | --- | --- |
| Brittle/discolored nail | 358 | 9.52 | 8.59 | 10.45 |
| Itchy skin | 1172 | 31.15 | 29.72 | 32.64 |
| Itchy, other | 192 | 5.1 | 4.44 | 5.85 |
| COVID toes (discoloration, swelling, painful, or blistering toes) | 490 | 13.02 | 11.99 | 14.14 |
| Other Skin and Allergy symptoms | 383 | 10.18 | 9.25 | 11.18 |
| Peeling skin | 488 | 12.97 | 11.91 | 14.06 |
| Petechiae (tiny purple, red, or brown spots on the skin, usually on arms, legs, stomach, buttocks, and occasionally inside mouth or on eyelids) | 671 | 17.84 | 16.64 | 19.09 |
| Skin rashes | 1045 | 27.78 | 26.34 | 29.24 |
| Visibly inflamed/bulging veins | 726 | 19.3 | 18.08 | 20.57 |

**Table S13.** Neuropsychiatric-Cognitive Functioning Symptom Prevalence Data.

| Symptom | Number of Respondents | Mean Prevalence | Lower CI | Upper CI |
| --- | --- | --- | --- | --- |
| Acute (sudden) confusion/disorientation | 691 | 18.37 | 17.15 | 19.64 |
| Agnosia (failure to recognize or identify objects despite intact sensory functioning) | 345 | 9.17 | 8.27 | 10.13 |
| Cognitive Dysfunction, overall (Brain fog) | 3203 | 85.14 | 83.94 | 86.26 |
| Difficulty problem-solving or decision-making | 2034 | 54.07 | 52.42 | 55.61 |
| Difficulty thinking | 2444 | 64.97 | 63.40 | 66.43 |
| Difficulty with executive functioning (planning, organizing, figuring out the sequence of actions, abstracting) | 2166 | 57.58 | 56.01 | 59.14 |
| Other Cognitive Functioning Symptoms | 323 | 8.59 | 7.71 | 9.52 |
| Poor attention or concentration | 2814 | 74.8 | 73.42 | 76.21 |
| Slowed thoughts | 1572 | 41.79 | 40.22 | 43.43 |
| Thoughts moving too quickly | 570 | 15.15 | 14.04 | 16.35 |

**Table S14.** Neuropsychiatric-Memory Symptom Prevalence Data.

| Symptom | Number of Respondents | Mean Prevalence | Lower CI | Upper CI |
| --- | --- | --- | --- | --- |
| Forgetting how to do routine tasks (tying your shoe laces, washing your hands) | 453 | 12.04 | 11.00 | 13.08 |
| Long-term memory loss (long-term memory can be anything from remembering yesterday, forgetting you've done a task, forgetting recently learned information, or forgetting your third-grade experience) | 1359 | 36.12 | 34.64 | 37.64 |
| Inability to make new memories | 275 | 7.31 | 6.49 | 8.19 |
| Other Memory Symptoms | 507 | 13.47 | 12.44 | 14.59 |
| Short-term memory loss (memory that lasts ~30 seconds, i.e. remembering a phone number before writing it down, or forgetting you're in the middle of a task) | 2438 | 64.81 | 63.34 | 66.37 |

**Table S15.**
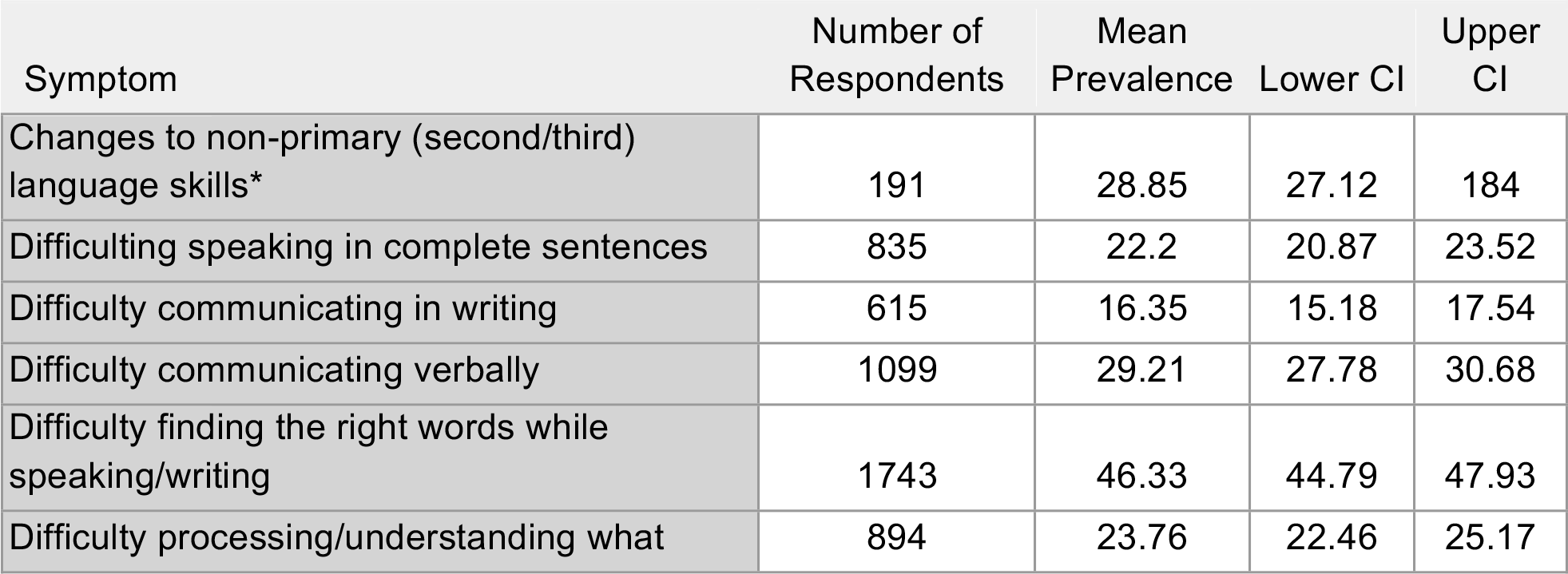
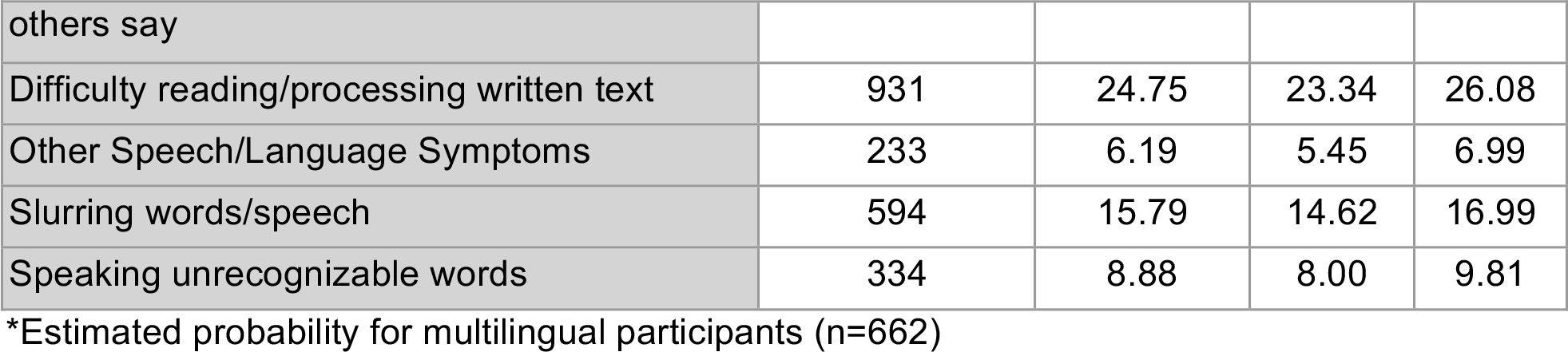
Neuropsychiatric-Speech and Language Symptom Prevalence Data.

**Table S16.** Neuropsychiatric-Sensorimotor Symptom Prevalence Data.

| Symptom | Number of Respondents | Mean Prevalence | Lower CI | Upper CI |
| --- | --- | --- | --- | --- |
| Dizziness / vertigo / unsteadiness or balance issues | 2531 | 67.28 | 65.76 | 68.77 |
| Electrical zaps/electrical shock sensation | 945 | 25.12 | 23.76 | 26.53 |
| Inability to cry | 184 | 4.89 | 4.25 | 5.61 |
| Inability to yawn | 250 | 6.65 | 5.87 | 7.47 |
| Neuralgia (nerve pain) | 1177 | 31.29 | 29.82 | 32.80 |
| Numbness/loss of sensation | 1332 | 35.41 | 33.86 | 36.90 |
| Numbness/weakness on one side of the body only | 472 | 12.55 | 11.48 | 13.61 |
| Seizures (confirmed) | 22 | 0.58 | 0.37 | 0.88 |
| Seizures (suspected) | 102 | 2.71 | 2.23 | 3.27 |
| Sensation of brain pressure | 1227 | 32.62 | 31.15 | 34.21 |
| Sensation of brain warmth/"on fire" | 461 | 12.25 | 11.22 | 13.32 |
| Sensitivity to noise | 1305 | 34.69 | 33.15 | 36.20 |
| Tingling/prickling/pins and needles sensation | 1852 | 49.23 | 47.66 | 50.82 |
| Tinnitus | 1280 | 34.02 | 32.56 | 35.57 |
| Tremors | 1511 | 40.16 | 38.62 | 41.76 |
| Vibrating sensations | 1610 | 42.80 | 40.34 | 44.13 |

**Table S17.**
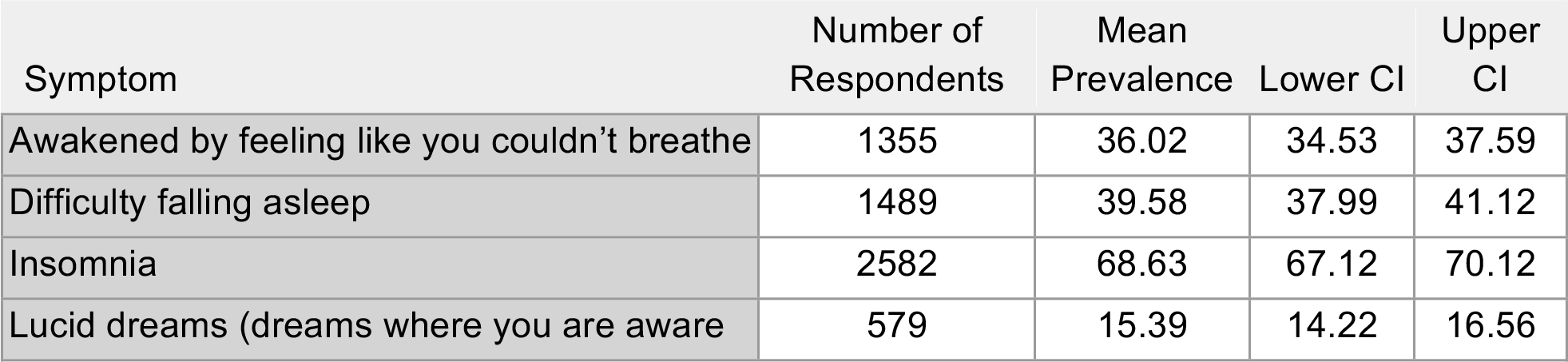
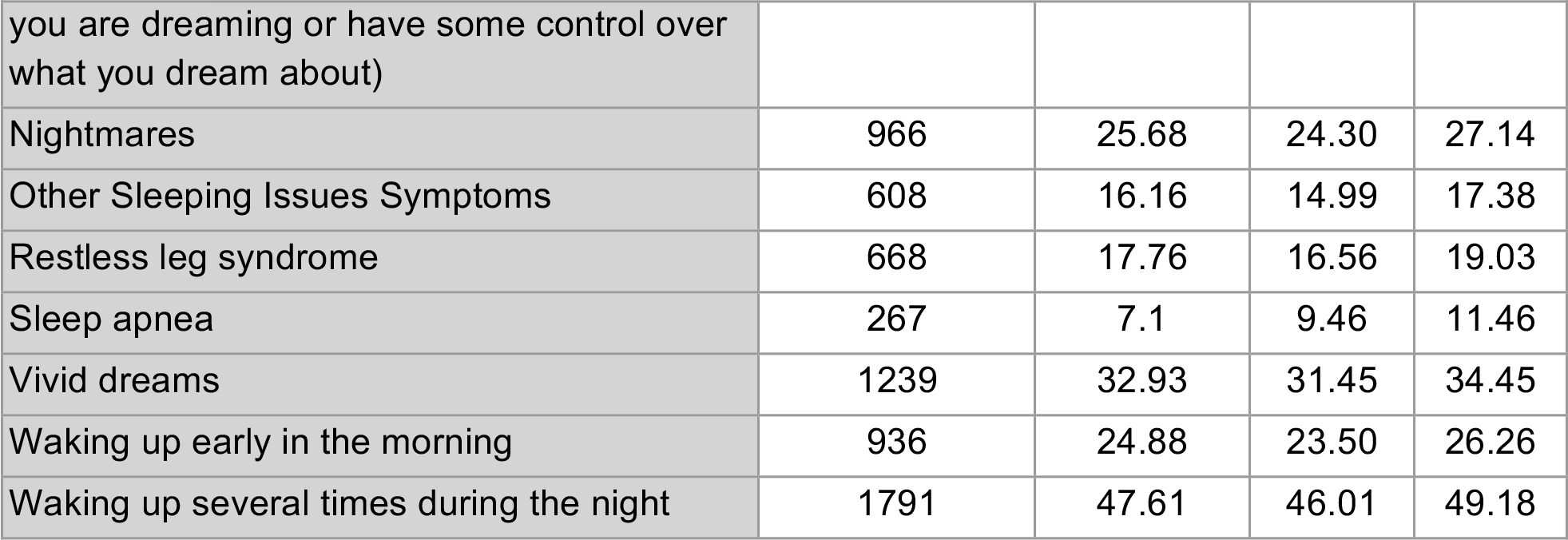
Neuropsychiatric-Sleep Symptom Prevalence Data.

**Table S18.**
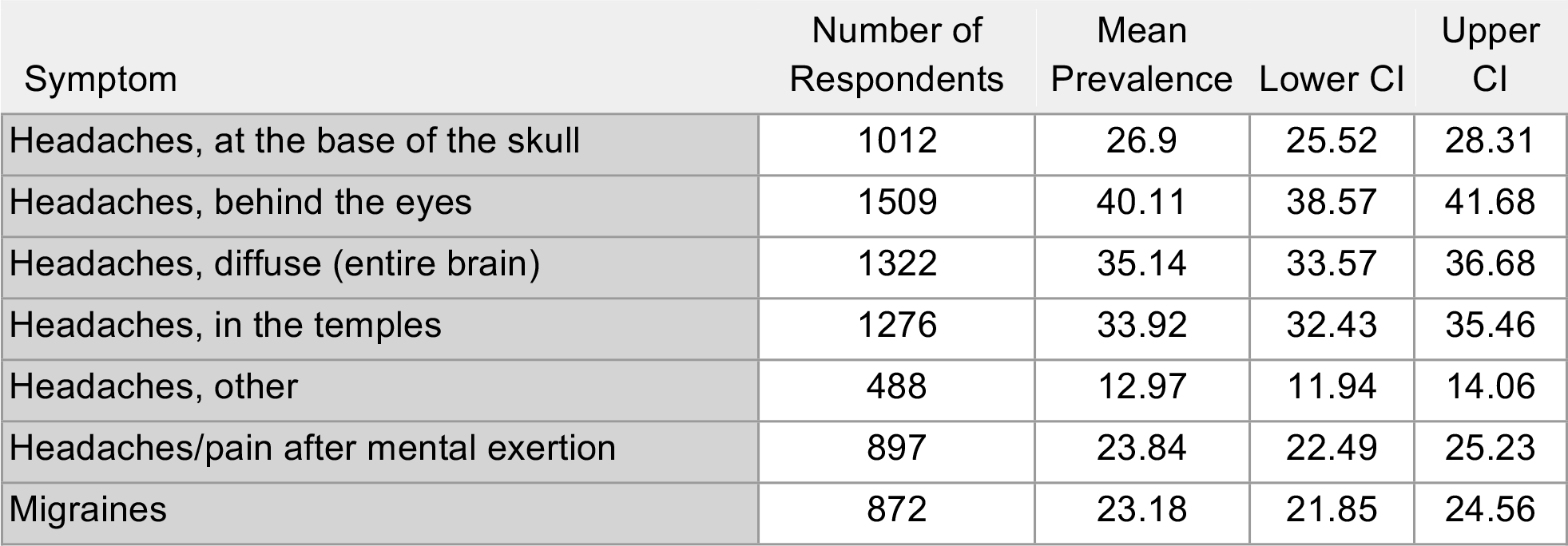
Neuropsychiatric-Headaches Symptom Prevalence Data.

| Symptom | Number of Respondents | Mean Prevalence | Lower CI | Upper CI |
| --- | --- | --- | --- | --- |
| Headaches, at the base of the skull | 1012 | 26.9 | 25.52 | 28.31 |
| Headaches, behind the eyes | 1509 | 40.11 | 38.57 | 41.68 |
| Headaches, diffuse (entire brain) | 1322 | 35.14 | 33.57 | 36.68 |
| Headaches, in the temples | 1276 | 33.92 | 32.43 | 35.46 |
| Headaches, other | 488 | 12.97 | 11.94 | 14.06 |
| Headaches/pain after mental exertion | 897 | 23.84 | 22.49 | 25.23 |
| Migraines | 872 | 23.18 | 21.85 | 24.56 |

**Table S19.**
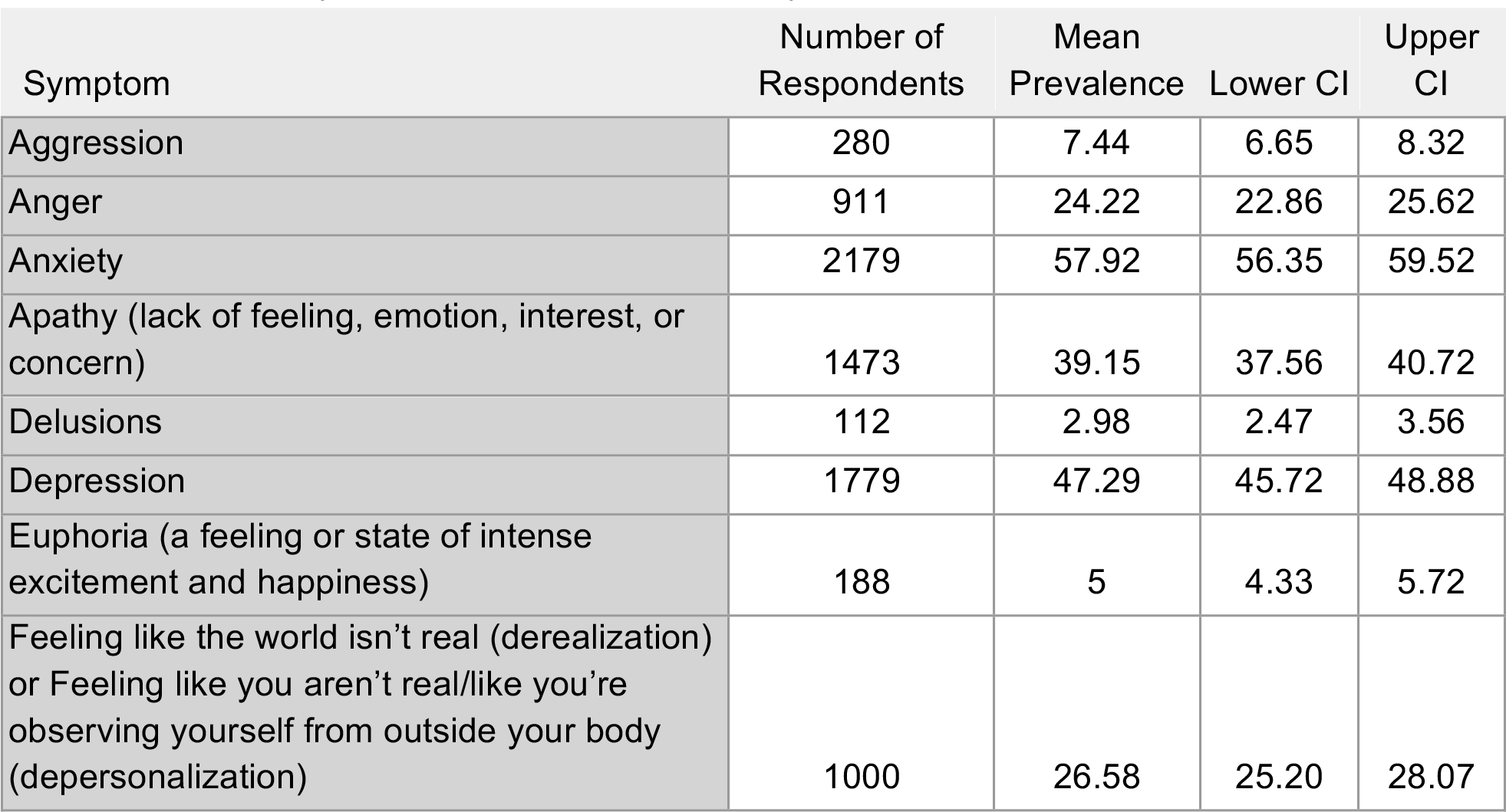
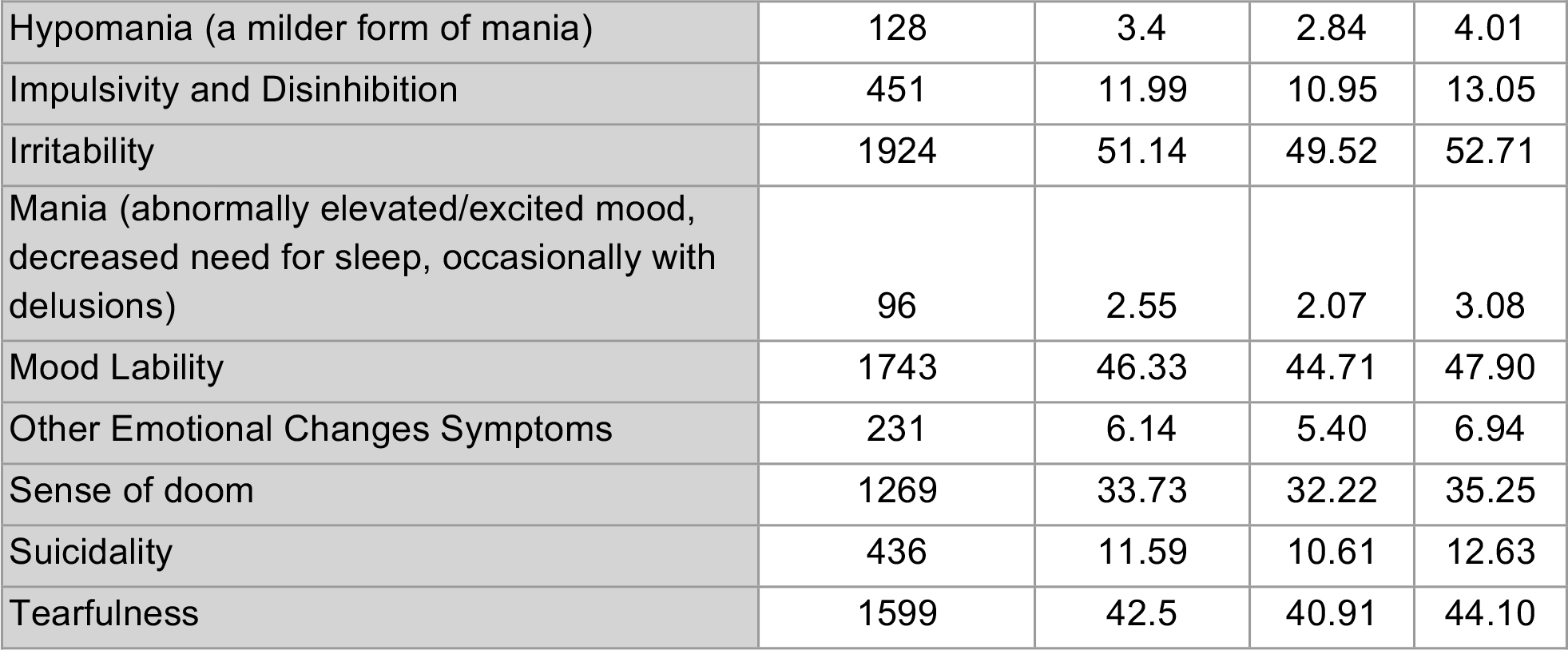
Neuropsychiatric-Emotion and Mood Symptom Prevalence Data.

| Symptom | Number of Respondents | Mean Prevalence | Lower CI | Upper CI |
| --- | --- | --- | --- | --- |
| Aggression | 280 | 7.44 | 6.65 | 8.32 |
| Anger | 911 | 24.22 | 22.86 | 25.62 |
| Anxiety | 2179 | 57.92 | 56.35 | 59.52 |
| Apathy (lack of feeling, emotion, interest, or concern) | 1473 | 39.15 | 37.56 | 40.72 |
| Delusions | 112 | 2.98 | 2.47 | 3.56 |
| Depression | 1779 | 47.29 | 45.72 | 48.88 |
| Euphoria (a feeling or state of intense excitement and happiness) | 188 | 5 | 4.33 | 5.72 |
| Feeling like the world isn't real (derealization) or Feeling like you aren't real/like you're observing yourself from outside your body (depersonalization) | 1000 | 26.58 | 25.20 | 28.07 |
| Hypomania (a milder form of mania) | 128 | 3.4 | 2.84 | 4.01 |
| Impulsivity and Disinhibition | 451 | 11.99 | 10.95 | 13.05 |
| Irritability | 1924 | 51.14 | 49.52 | 52.71 |
| Mania (abnormally elevated/excited mood, decreased need for sleep, occasionally with delusions) | 96 | 2.55 | 2.07 | 3.08 |
| Mood Lability | 1743 | 46.33 | 44.71 | 47.90 |
| Other Emotional Changes Symptoms | 231 | 6.14 | 5.40 | 6.94 |
| Sense of doom | 1269 | 33.73 | 32.22 | 35.25 |
| Suicidality | 436 | 11.59 | 10.61 | 12.63 |
| Tearfulness | 1599 | 42.5 | 40.91 | 44.10 |

**Table S20.** Neuropsychiatric-Taste and Smell Symptom Prevalence Data.

| Symptom | Number of Respondents | Mean Prevalence | Lower CI | Upper CI |
| --- | --- | --- | --- | --- |
| Altered sense of smell | 745 | 19.8 | 18.53 | 21.08 |
| Altered sense of taste | 943 | 25.07 | 23.68 | 26.45 |
| Heightened sense of smell | 323 | 8.59 | 7.71 | 9.49 |
| Heightened sense of taste | 101 | 2.68 | 2.18 | 3.22 |
| Loss of smell | 1352 | 35.94 | 34.42 | 37.51 |
| Loss of taste | 1267 | 33.68 | 32.16 | 35.22 |
| Phantom smells (imagining/hallucinating smells - smelling things that aren't there) | 872 | 23.18 | 21.88 | 24.56 |
| Phantom taste (imagining/hallucinating tastes - tasting things when there's nothing in your mouth) | 339 | 9.01 | 8.16 | 9.97 |

**Table S21.**
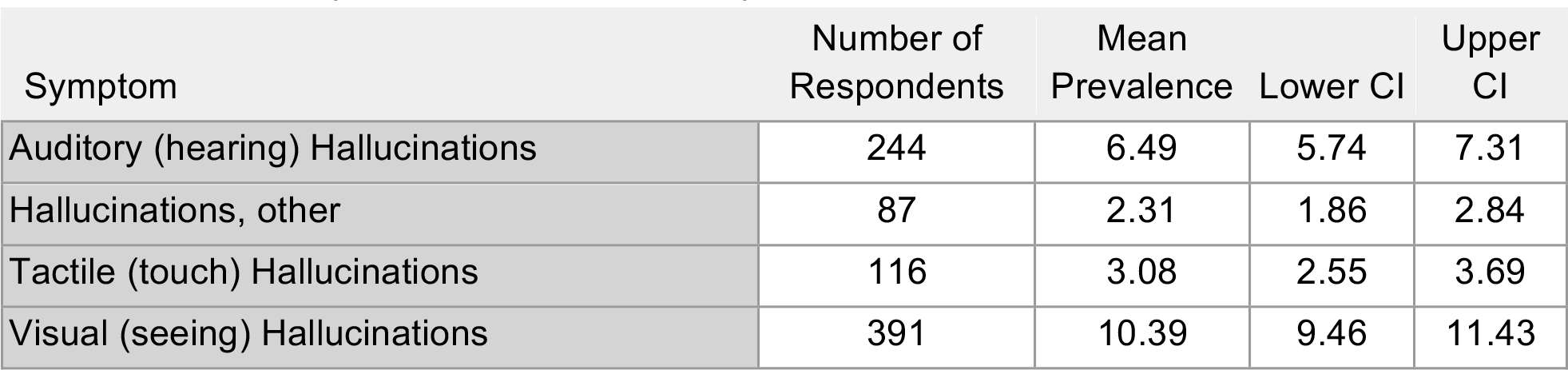
Neuropsychiatric-Hallucinations Symptom Prevalence Data.

**Table S22.**
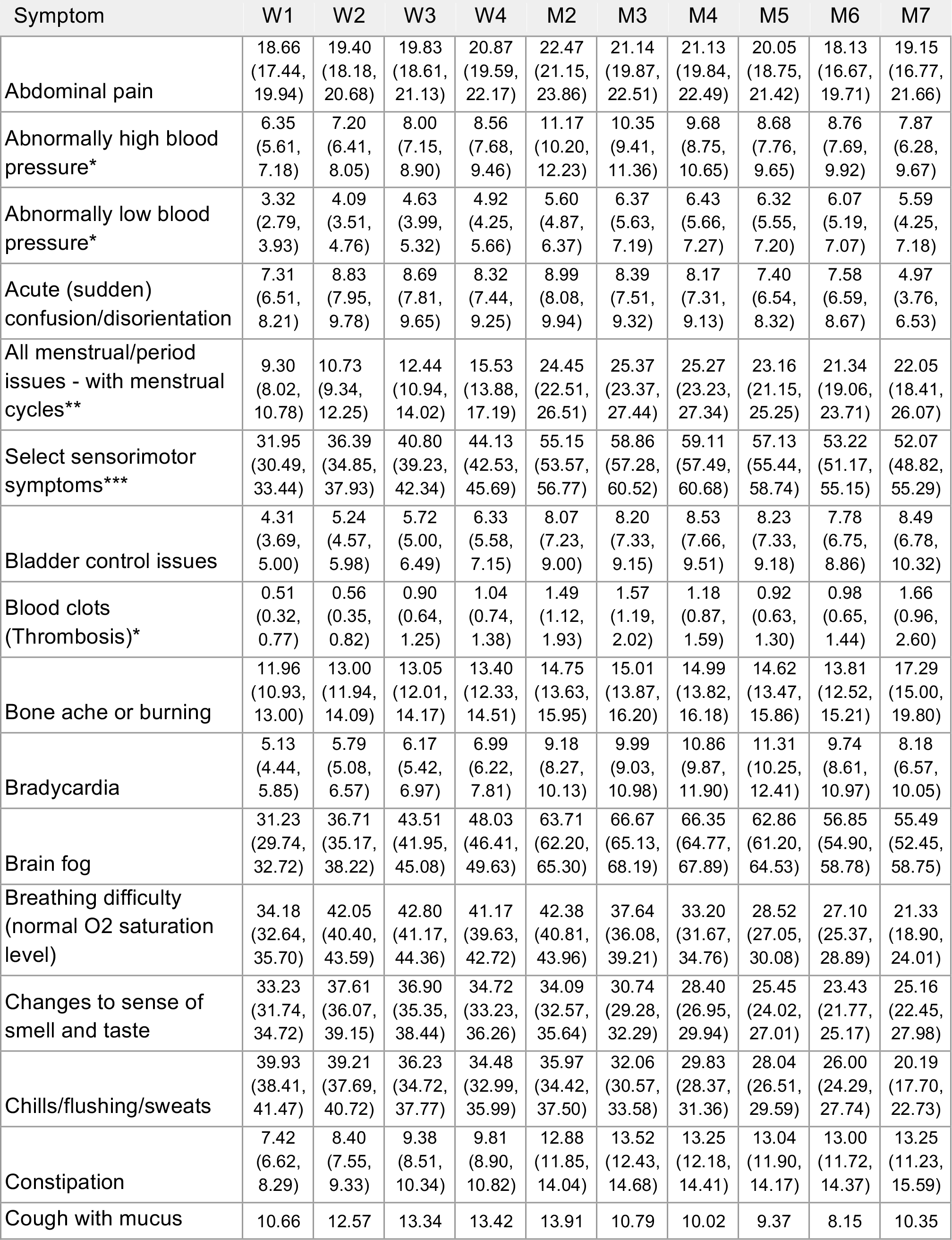
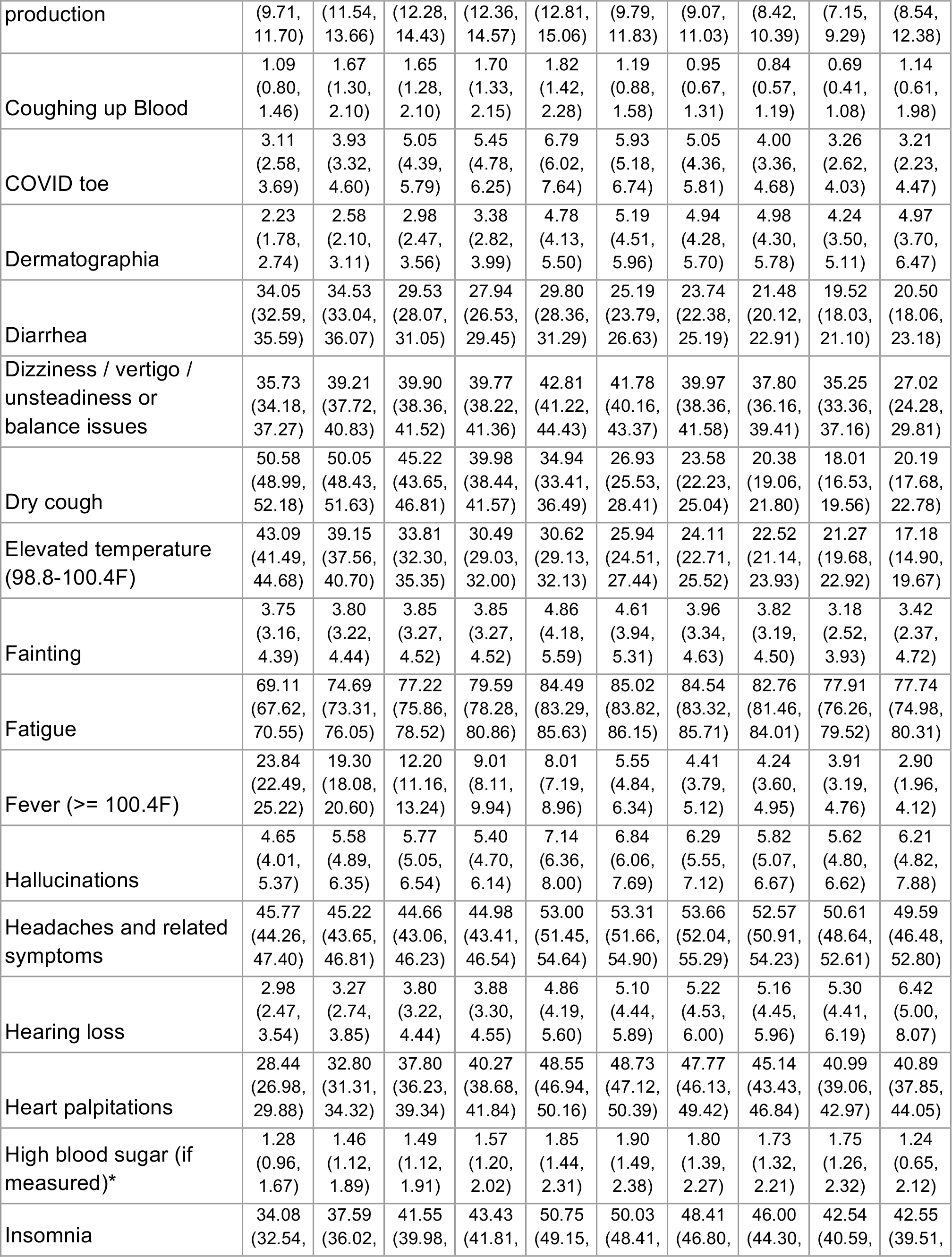
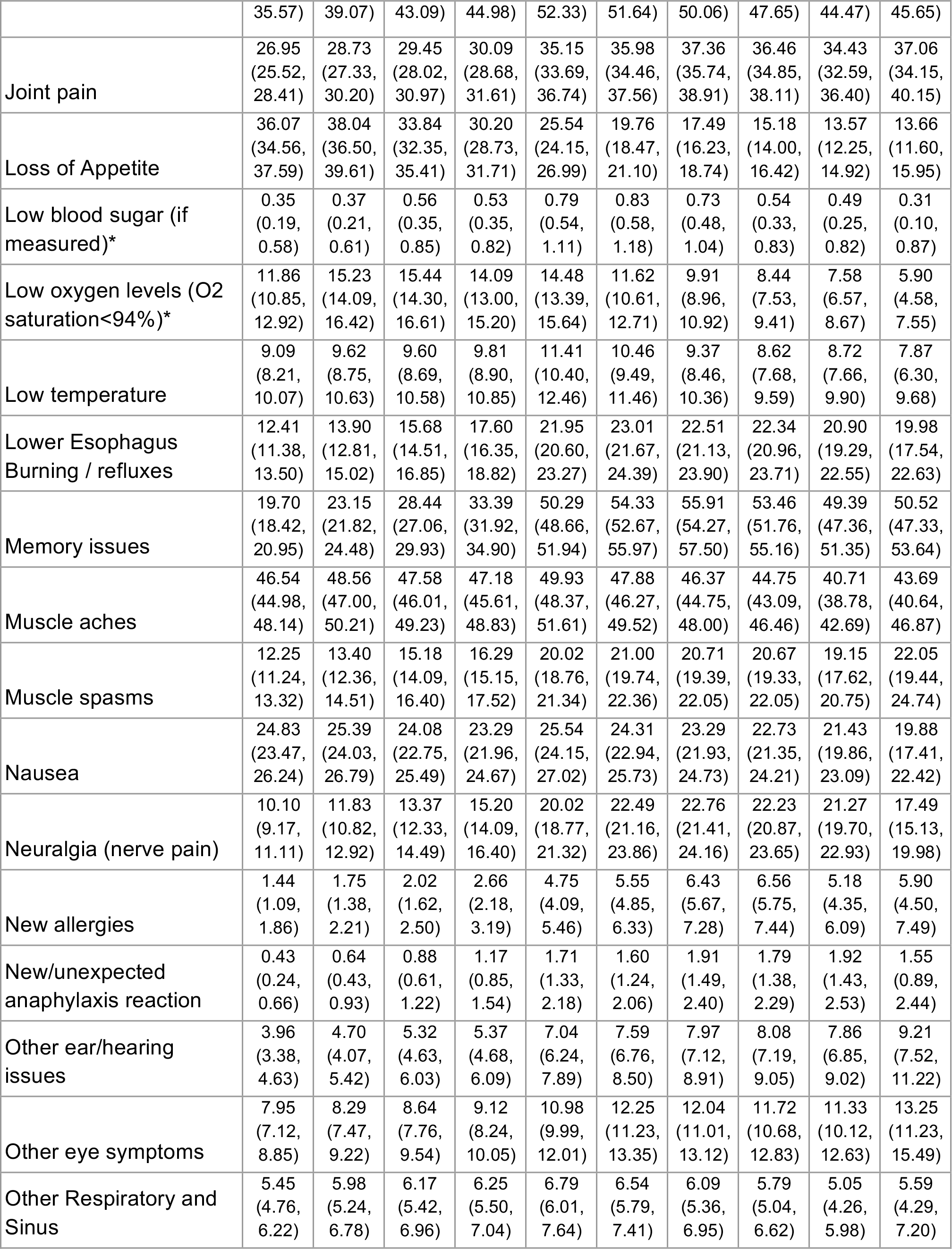
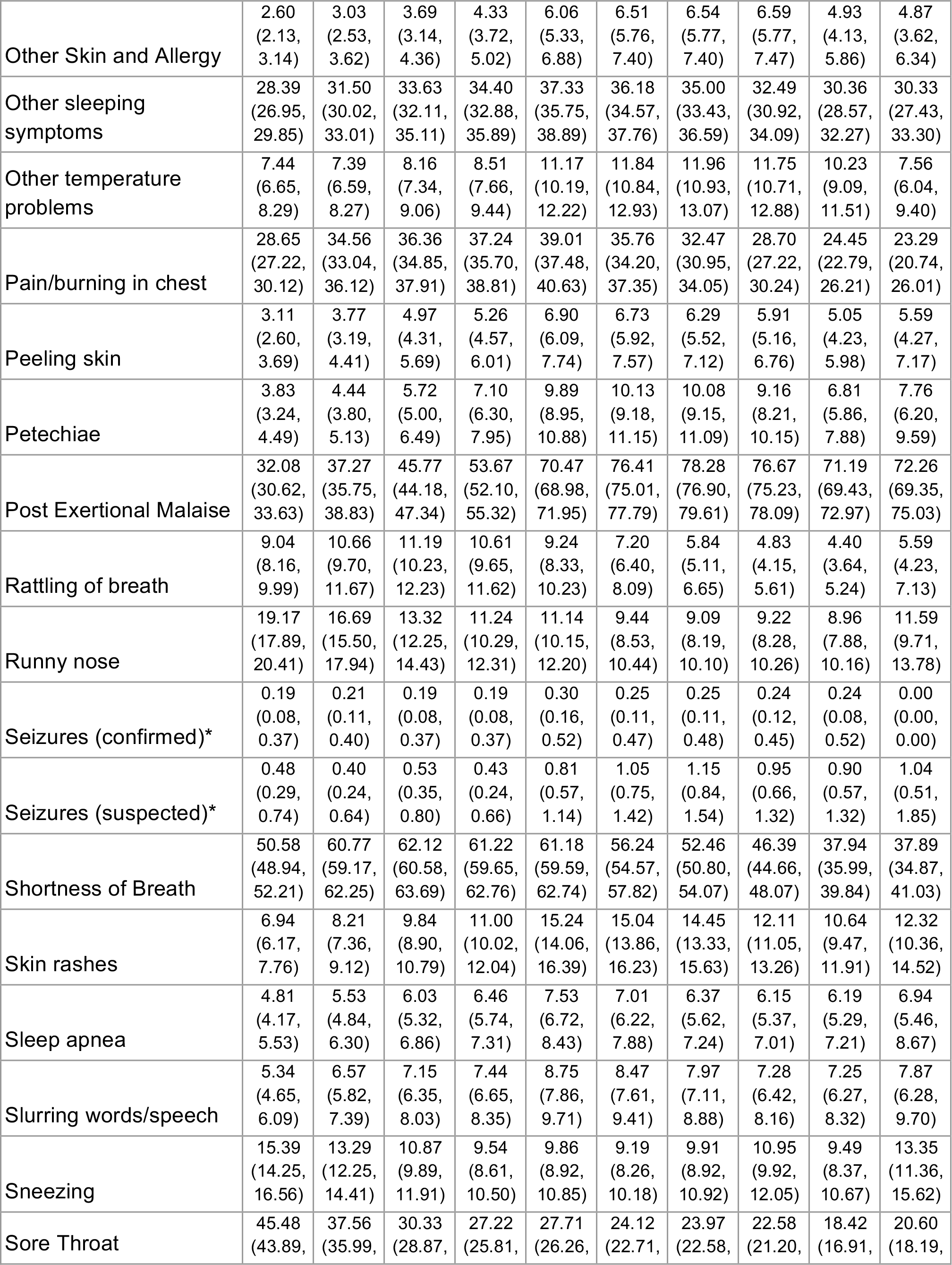
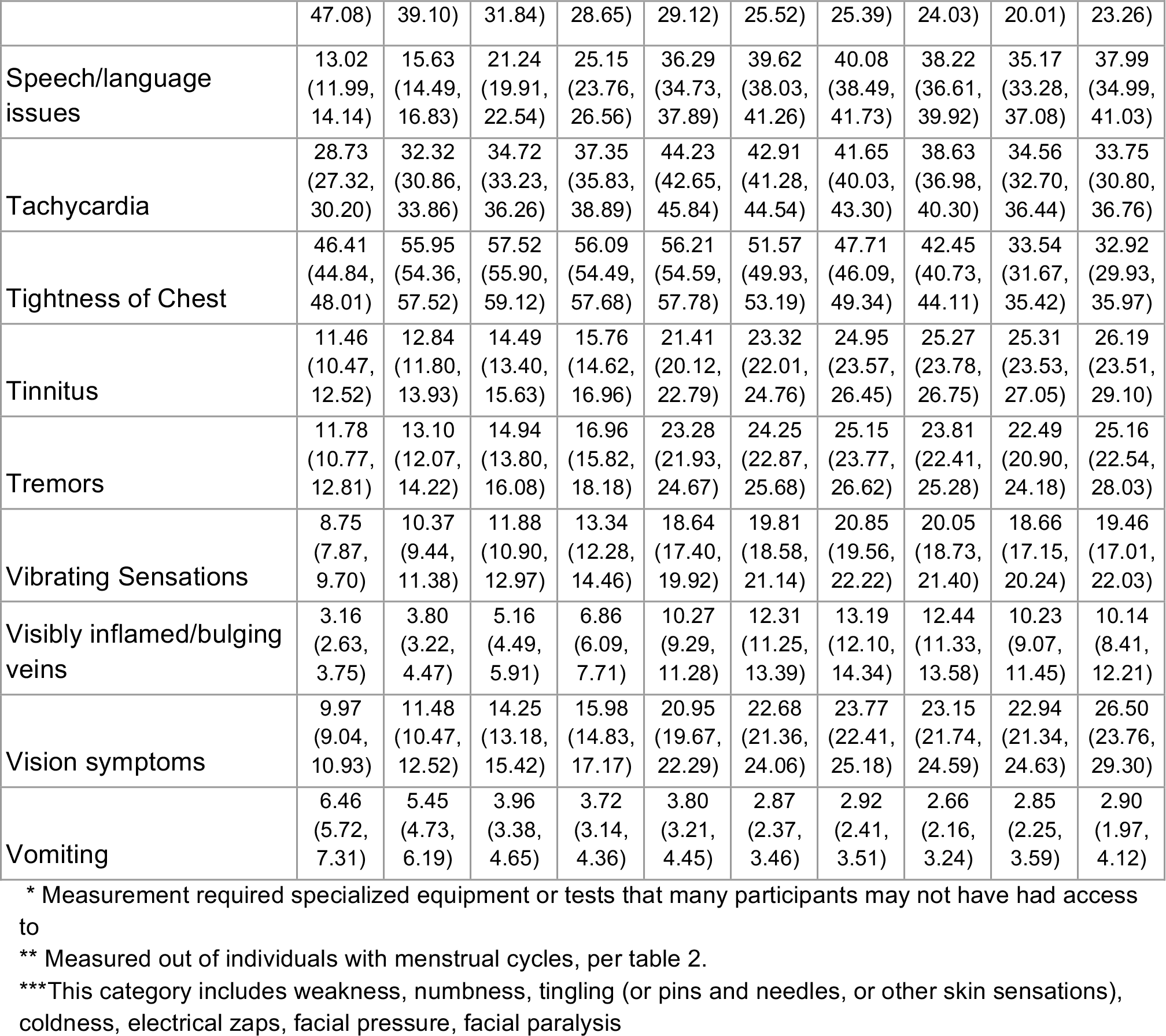
Symptom Prevalence Timecourse Data: Mean (Lower CI, Upper CI)

## Notes

### Competing Interest Statement

The authors have declared no competing interest.

### Funding Statement

Authors are group of volunteers. Survey expenses were covered by AA's research fund from Sainsbury Wellcome Centre (Wellcome Trust and Gatsby Charity). Some donations to Patient-Led Research for COVID-19 were directed towards graphic design work.

### Author Declarations

This study was approved by the UCL Research Ethics Committee [16159.002], and Oregon Health and Science University, Portland, Oregon, USA, with UCL serving as primary site. Weill Cornell Medical College Institutional Review Board (IRB) exemption was obtained. All participants gave informed consent.

### Summary of Updates

5 new supplemental figures focusing on the following analyses have been added to the manuscript, as well as all raw data. - Further comparisons between tested and untested participants (survival analysis, and symptom prevalence and time course) - survival analysis for Male vs Female participants - symptom severity and count for recovered vs unrecovered participants - All raw data (prevalences) are added to the supplementary

